# The Relationship Between Biological Aging with Interdisciplinary Health Indicators: A Scoping Review

**DOI:** 10.1101/2024.10.31.24316398

**Authors:** Jennifer T. H. Reeves, Emma Knock, Zoë M. Gilson, Leah Derry, Miki C. McGhee, Katherine Taylor-Hood, Alison Ziesel, Hosna Jabbari, Lorelei Newton, Jo Ann Miller, Jie Zhang, Ryan Rhodes, Anastasia Mallidou, Theone S. E. Paterson

## Abstract

**Objectives:** This review aims to provide a comprehensive examination of biomarkers and interdisciplinary variables related to aging.

**Methods:** This scoping review included studies which involved adult participants, and which reported on the relationship between any biomarker or biological age with chronological age.

**Results:** After screening, 447 articles met the selection criteria. Results were categorized into 10 distinct categories through an iterative process.

**Conclusions:** This review contributes information regarding the interdisciplinary influences on the rate of aging. Telomere length was the most commonly examined biomarker, and Horvath’s 353-CpG Pan-Tissue clock was the most common clock, with both demonstrating a strong and consistent relationship with chronological age. The interdisciplinary variables demonstrated relationships with biological aging with varying strengths and consistencies.

## Introduction

Biological aging, also referred to as epigenetic aging, is age-related declines to the human body and its functioning. The rate of biological aging compared to chronological aging varies between individuals, with individuals exhibiting either accelerated, decelerated, or equivalent rates of aging. Due to the implications that aging has for health outcomes, treatments, and mortality, it is important to distinguish and delineate the variables that may slow or quicken the rate of aging, especially those which may be modifiable.

Certain demographic variables are associated with variances in the rate of aging. For example, past research has identified that individuals who are female have longer telomeres, and thus a slower rate of aging.^1,2^ Socioeconomic status throughout the lifespan also plays a role in aging; parental income and education during childhood as well as current income during adulthood has been linked to accelerated biological aging for both children and adults.^3,4^ Other demographic variables also have an interaction; for example, racial differences in aging are stratified by socioeconomic status, with the greatest difference in telomere length (TL) between individuals who are white compared to black when of lower socioeconomic status.^5^

An individual’s lifestyle also has implications for aging. This includes the consumption of various substances, such as nicotine and alcohol. Generally, the relationship between cigarettes and accelerated aging is an established relationship, with smokers having shorter telomeres than non-smokers, even at low levels of smoking.^2,6^ In comparison, studies reporting the relationship between biological aging and alcohol consumption are more diverse in their findings. For example, while some researchers have reported differences in the rate of aging dependent on the level of consumption,^6^ other researchers have found an increased rate of aging at all levels of consumption.^7^ Drug use of illicit substances (e.g., methamphetamine, opioids) are associated with reduced TL.^8^ Another aspect of an individual’s lifestyle that can impact their health and aging includes their engagement in physical activity and sedentary behaviour. Physical activity may be an overall benefit to aging,^9^ as Tucker and colleagues^10^ found that individuals who engaged in exercise had a reduced rate of aging compared to those who are sedentary. However, there is currently no consensus regarding the effect of exercise on aging, with particular differences in the optimal level of exercise; while some researchers have found an overall benefit of exercise compared to sedentary behaviour,^9,10^ other researchers have not found this.^2^ Diet and the consumption of specific foods can also play a role in biological aging. For example, the consumption of legumes, nuts, seaweed, fruit, dairy products, coffee, and high-fiber foods is associated with longer TL, whereas red meat, processed meat, and high sugar products (e.g., sugar-sweetened soda) are connected to reduced TL.^9^ Certain diets, such as the Mediterranean diet (comprised of primarily vegetables, fruits, and fish), have been linked to a slower rate of aging.^9^

Various health indicators can also impact aging. The presence of certain chronic diseases may also be risk factors for advanced aging, such as digestive, endocrine, and cardiometabolic diseases.^11^ For example, past research indicates that frailty increases with both chronological age^12^ and biomarkers of biological age,^13^ and is associated with a higher mortality risk predictor of mortality.^13^ Anthropometric measures (e.g., BMI) may also play a role in biological aging, as one study indicated that an increase of 10 body mass index (BMI) units increases biological age of liver tissue by approximately 3.3 years.^14^ Mental health is also important; both the presence and severity of depressive symptoms is relayed to an increased biological age.^11,15^ Finally, early childhood experiences may alter the rate of aging, as Jensen and colleagues^11^ reported faster aging for those who reported experiencing childhood trauma.

Estimated biological age is also impacted by the method of estimation. Certain biomarkers are commonly used as a means to assess aging. TL is the most commonly examined indicator of aging.^16^ Other biomarkers used for this purpose include specific genes, and DNA methylation.^16^ Biomarkers are also used in algorithms to determine the rate of biological aging, such as the Klemera-Doubal method (KDM),^17^ which is a complex calculation with biomarkers as well as chronological age. Another common method of estimating biological aging involves CpGs,^27,2818^ which can involve individual CpGs, or models comprised of CpGs, such as the 99-CpG model and 3-CpG model.^18,19^ Most epigenetic clocks also utilize CpGs to calculate biological age. For example, the Horvath clock uses 51 tissues and cell types, and a total of 353 CpG sites,^20^ whereas the Hannum clock is an aging model that is comprised of 71 CpG methylation markers.^21^ In comparison, the DNAm GrimAge is a calculated age, with higher numbers representing an increased mortality/morbidity risk.^22^

It is essential to identify the impact of both modifiable and fixed factors that impact aging, as this has implications for health outcomes and treatment. The utility of biological aging in healthcare decisions is clear, as biological age a is significantly better predictor of mortality than chronological age.^12,23^

Some of the relevant past reviews have evaluated the benefits and challenges of available methods of biological age prediction,^16,24^ whereas others focused primarily on certain types of biomarkers, such as those related to vascular aging.^25^ Another review examined the negative impacts of aging, and possible strategies and policies that could improve slow aging and improve health outcomes.^26^ However, due to their limited scope, past reviews have not examined the impacts of indicators related to biological aging from an interdisciplinary approach, which would greatly enhance the depth of understanding on this topic. In addition, due to inconsistencies between studies in methodology (e.g., indicators of biological aging used, method of biological age prediction), and in study findings, it is essential to synthesize all current research findings into a comprehensive review. Thus, our review seeks to fill an important gap in knowledge and extend beyond past reviews by providing a more comprehensive and in-depth examination of variables that impact aging, methods of ascertaining biological age, and the overall relationship between chronological and biological aging. This review will inform clinicians, healthcare providers, and policy makers as to what impacts aging, and what modifiable factors to target.

To this effect, our review posed two research questions:

1. What is the relationship between biomarkers and calculations of biological aging with chronological aging?
2. What interdisciplinary health variables impact the rate of aging relationship?

## Methods

This review adhered to guidelines in the PRISMA Extension for Scoping Reviews (PRISMA-ScR),^27^ and was guided by Arksey and O’Malley’s methodological framework.^28^

### Search strategy

Four databases were searched from 2011 to August 2021, including PsycINFO, CINAHL, PubMed, and SPORTDiscus (see **Appendix A**). Our search terms included terms associated with biological aging and chronological aging, with all terms searched in the abstract and title. In addition, searches included relevant MeSH headings. Subsequently, limiters were applied to our search, including a publication data filter (2011 to August 2021), subject (humans), age (adults aged 18 or older), and language (English).

## Eligibility criteria

### Population

To be eligible for inclusion, studies needed to include only human participants, aged 18 or older. These studies were included irrespective of if they examined healthy adults, or adults with specific health conditions (e.g., cancer). Studies which included animals or individuals under 18 years of age were excluded.

### Outcomes

Studies needed to include chronological aging and compare either a calculated biological age or an indicator of biological aging (e.g., TL) with chronological age. Studies which examined only the biological aging of a specific organ (e.g., the heart, liver) were excluded.

### Design

Empirical studies were considered for inclusion if they were peer reviewed and published in a journal. Non-research (e.g., opinion pieces, commentaries, letters, editorials), grey literature (e.g., theses, dissertations), theoretical papers, seminar papers, case studies, book chapters, study protocols, review articles, and articles in-press were excluded.

### Other considerations

In order to examine the most recent scientific findings, only studies published in the last ten years from the initial literature search (2011 to present) were included. In addition, only studies published in English were considered, but studies were eligible for inclusion irrespective of the country they occurred in.

### Study selection and charting the data

Five reviewers (JR, VP, RS, LF, MM) used the software program Covidence^29^ to screen articles according to their title and abstracts. Two reviewers examined each title/abstract, and any conflicts during this step were resolved by the third reviewer. Subsequently, full text articles were screened according to our inclusion criteria (see **Table S1**). Disagreements were resolved by the three reviewers re-examining the articles and discussing the article in comparison to our inclusion criteria.

Data was extracted by four reviewers (JR, AZ, MM, EK), with data extracted initially by one reviewer including the aims, population, methodology, and findings of the studies. Extraction was then verified by one reviewer.

### Data synthesis

Results of included studies are examined in a narrative format. Following data extraction, a composite list of all outcomes which were compared to chronological and/or biological indicators of aging was composed. Subsequently, categories of outcomes were created through an iterative process, in which variables were sorted by their commonalities into themes of outcomes, with groups created and changed throughout this process. Working definitions were created for these categories. This resulted in 10 categories of outcomes; category definitions and a non-exhaustive list of example variables are described in **Table 1**.

**Table 1:**
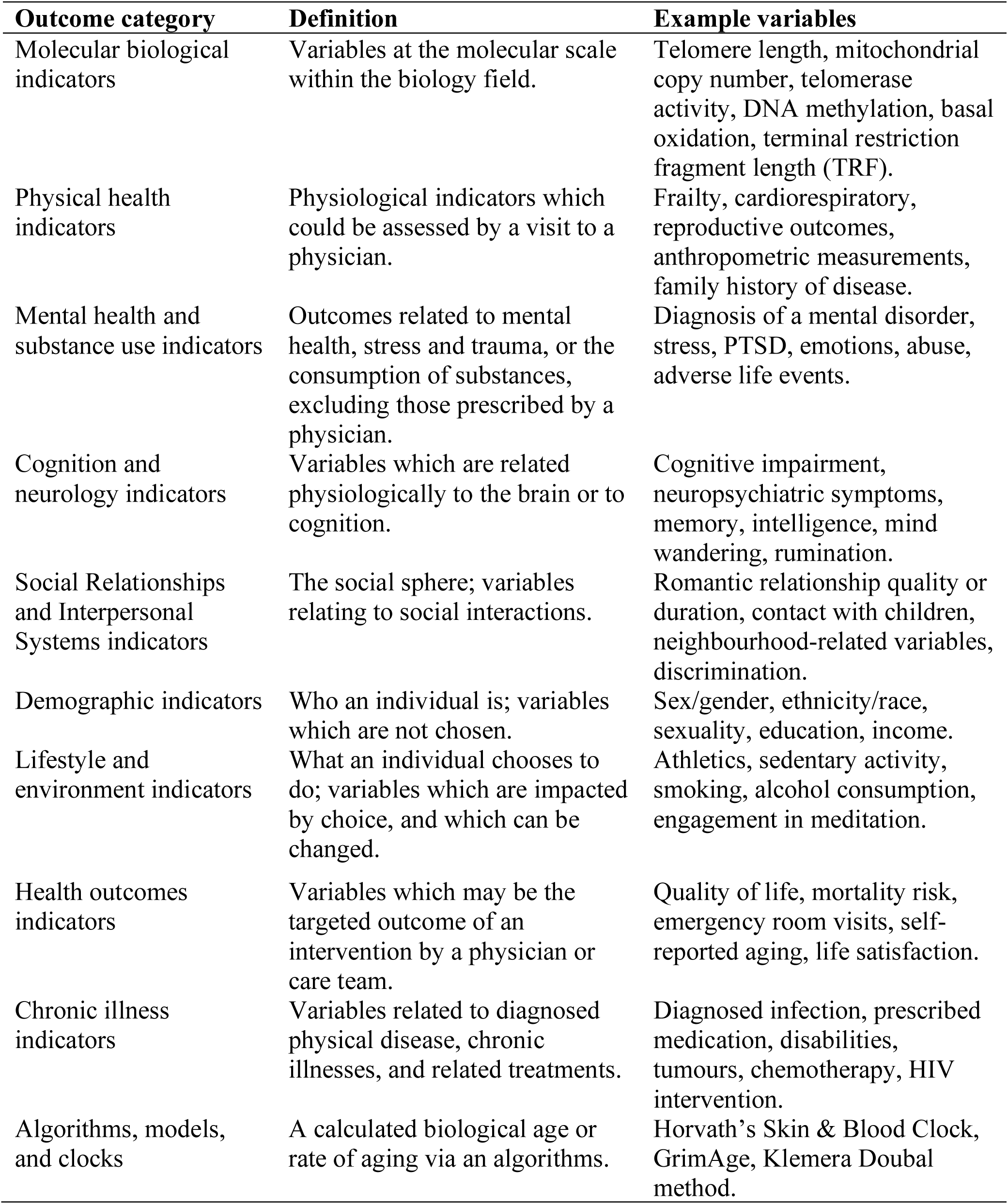
Outcome Categories of Included Studies.

## Results

A total of 1244 records were retrieved from which 317 duplicates were removed (**Figure 1**). Subsequently, 927 records were screened for title and abstract and 231 were removed. We sought 696 full-text articles, of which one could not be retrieved, and excluded another 248 based on our a priori inclusion and exclusion criteria. Afterwards, 447 articles were included in our scoping review. An overview of the results for each of the 10 categories of outcomes is demonstrated in **Figure 2**.

**Figure 1:**
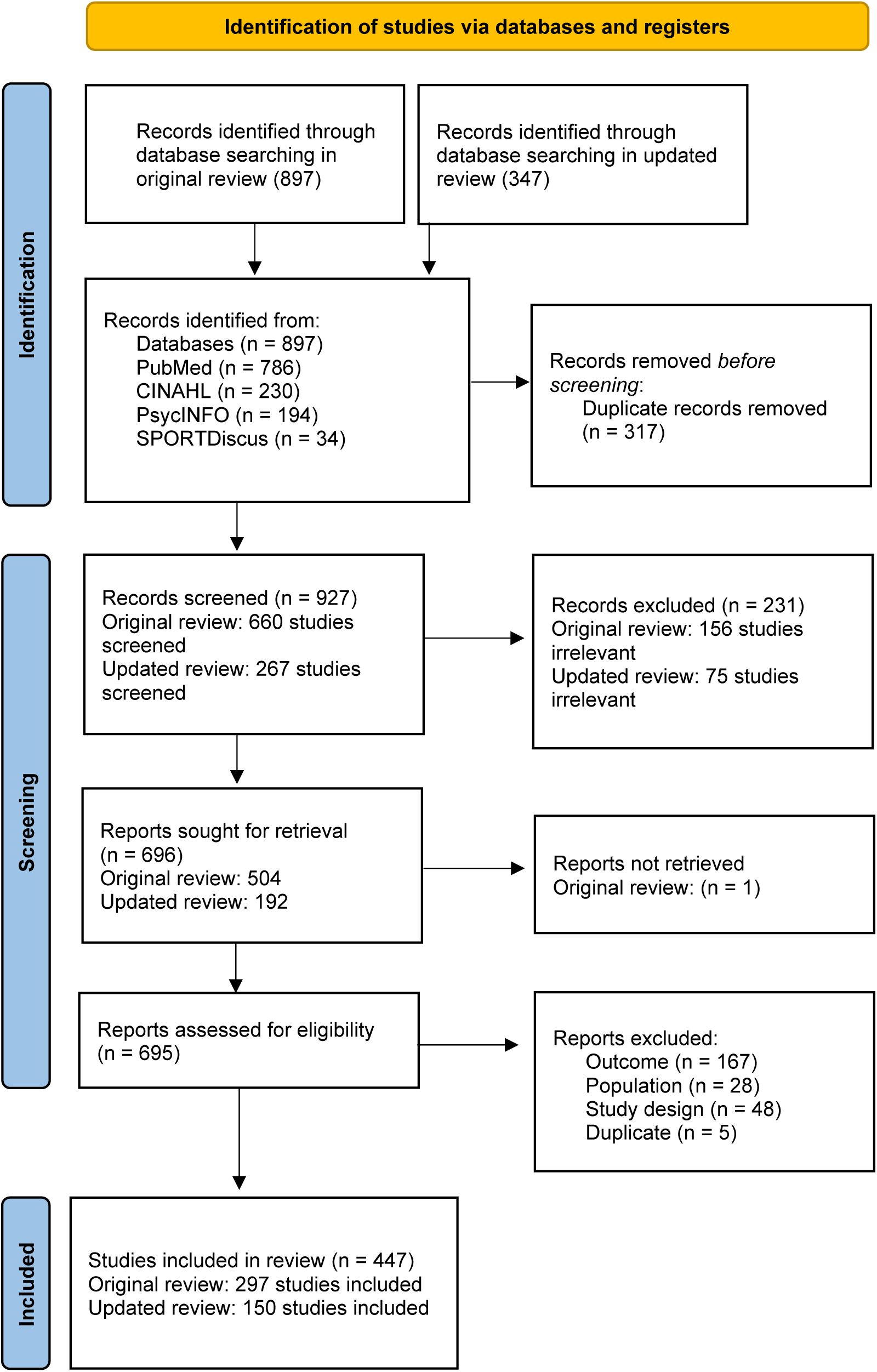
PRISMA Flow Diagram

**Figure 2:**
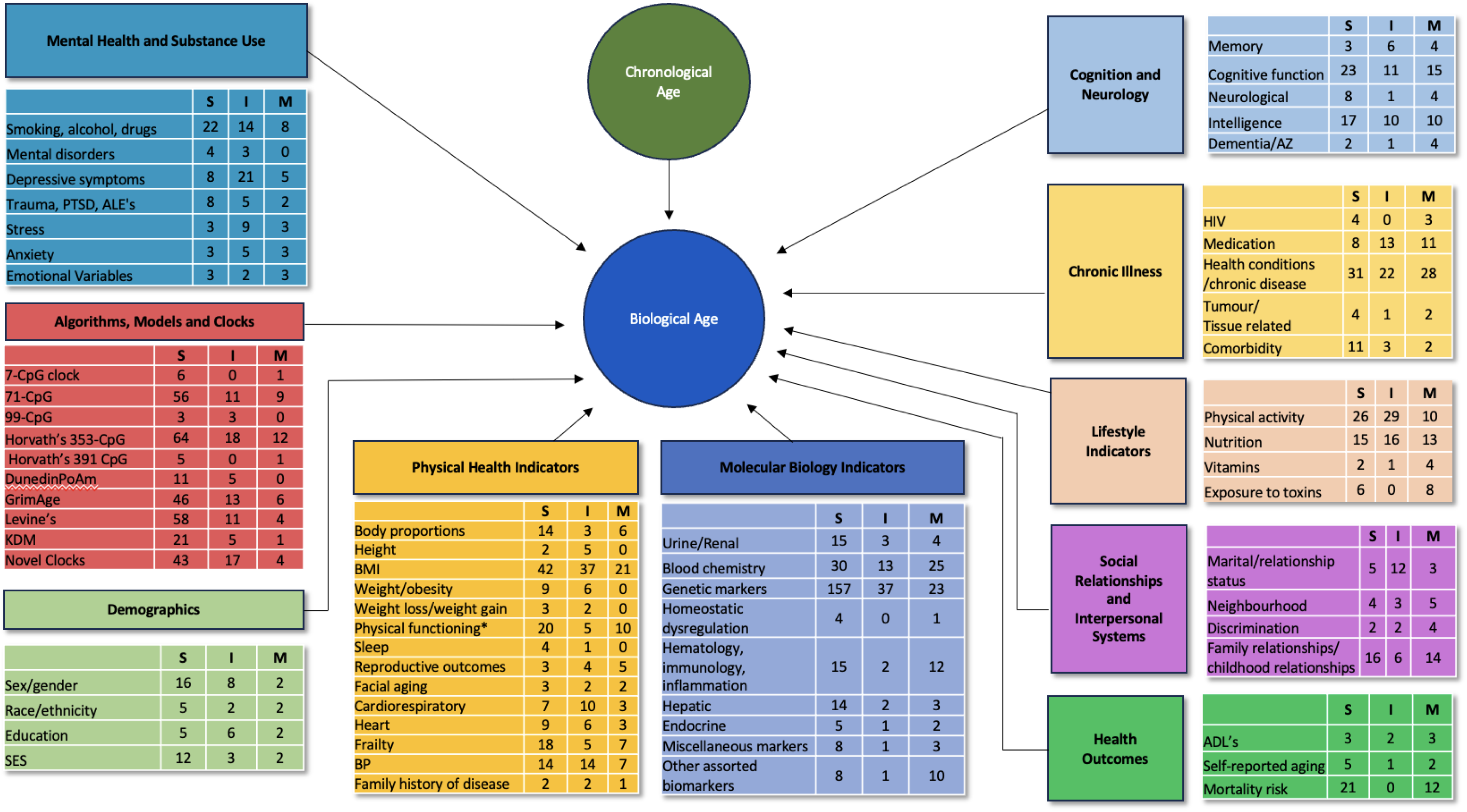
Overview of Aging-Related Variables

### Study characteristics

In total, the number of participants in the included manuscripts is 3,480,789. However, due to overlap in datasets between some studies, the number of unique participants is lower. A total of 184 studies used the same single dataset as another study; the most commonly used datasets include NHANES in 20 manuscripts, Health and Retirement Study (HRS) in 17, Family and Community Health Study (FACHS) population in ten, Dunedin cohort in nine, the Lothian Birth Cohorts 1921 and/or 1936 in seven, and the Normative Aging Study (NAS) population in six.. Forty-four studies used data from a combination of datasets. The remaining 219 studies used unique data.

A variety of populations were examined in our included studies. Over a third (125) of the included studies examined a general population. Some studies selected participants based on their age; two studies focused on young adults, nine studies included young and middle-aged adults, 17 studies focused solely on middle-aged adults, and 99 studies assessed middle-aged and older adults. In addition to this, five studies included long-lived individuals (e.g., centenarians) and/or their family members. Seventy-two studies included participants with specific physical health conditions, which includes 12 studies that included cancer patients and/or survivors, and nine studies with HIV+ individuals, and 51 studies with other health conditions (e.g., hypertension, diabetes) whereas 15 studies included participants who were healthy Another 24 studies included participants with specific mental health or cognitive conditions, and three studies recruited participants based on their substance use. Some studies examined populations specific to lifestyle facets, including nine which focused on athletes, two with smokers, and studies which examined other conditions such as participation in a wellness program, being married, being in a heterosexual relationship, and meditation. Career professions (e.g., helicopter pilots, police officers, veterans) were examined in 22 studies, culture in one study (i.e., Berne Indiana Amish), and 11 twin studies. Studies were selective in terms of race or ethnicity in 24 studies, including 18 with Black individuals, five with Caucasian individuals, and one study with Indigenous Americans. Other studies examined former Swiss indentured child labourers (’Verdingkinder’), fibroblasts from a male young adult, human remains from cemeteries, individuals exposed to certain compounds, and individuals deemed to be most and least deprived. Of these studies, some also specifically recruited by gender or sex, including 40 studies with females only, and 25 studies with only men.

Included studies were also categorized by their discipline. Just under half of the included manuscripts (207 manuscripts) fell within the health discipline, an additional 4 studies in medicine and pharmacology, 68 in the combined psychology, psychiatry, neurology, and substance use discipline, 78 lifestyle, 36 biological, 17 techniques and analyses, nine general aging, seven demographics, six genetics, 13 social sciences, and two in the mortality field.

Studies occurred in 43 countries, with the most common including the United States (167 manuscripts), followed by 25 in China, and 23 in the United Kingdom. Twenty-six studies occurred in multiple countries, and 4 studies did not report the country they occurred in.

## Demographic indicators

### Sex/Gender

A total of 132 included studies examined gender or sex in relation to aging. Overlap in datasets occurred in the included studies; six studies completed their analyses with NHANES data ^5,30–34^, three with the HRS population,^35–37^ three with Netherlands Study of Depression and Anxiety population,^38–40^ two with the FACHS population,^41,42^ two with the Korean Genome Epidemiology Study,^43,44^ two with the Multiethnic Study of Atherosclerosis (MESA) study population,^45,46^ two with the Sympathetic Activity and Ambulatory Blood Pressure in Africans study population,^47,48^ two with the same data involving heterosexual couples,^49,50^ and two used the same three datasets: Swedish Adoption/Twin Study of Aging (SATSA), Origins of Variance in the Oldest Old (OCTO-Twin), and Aging in Women and Men: A Longitudinal Study of Gender Differences in Health Behavior and Health among Elderly (GENDER).^51,52^ Of note, researchers of many of the included studies typically did not specify if they assessed sex or gender, and often used the terms interchangeably. It is thus unclear whether the researchers ascertained the participants biological sex or their gender identity. In this paper, these studies and variables have been grouped together due to this ambiguity.

Sex was most commonly compared with TL. Across the diverse study populations described in **Table S2**, a total of 65 papers reported on this relationship, with 31 finding that women had significantly longer TL,^32,36–40,45,47,48,53–74^ one found that men had significantly longer TL (all P < .05),^75^ 31 reported insignificant differences,^5,33,35,43,44,50,76–99^ and two found mixed results.^100,101^ These two studies found that age interacted with gender; Huang and colleagues^100^ reported that TL was significantly shorter among women at wave one of their study (P < .01), but this effect was no longer significant by wave three due to women having a decreased rate of shortening. In contrast, Robertson and colleagues^101^ reported significantly shorter telomeres among women in their 1950 and 1930 younger cohorts, equivalent to being five and 11 years younger biologically, but no significant difference in TL between men and women in the 1970s cohort.

A total of 47 studies compared gender or sex with a biological clock, including the Horvath Pan-Tissue clock, Hannum’s epigenetic clock, Levine’s PhenoAge clock, novel clocks based on various biomarkers, brain-PAD, and calculations with the Klemera Doubal Method.

The majority of these (29 studies) found significant differences, as nine studies reported that women were significantly biologically younger than men,^60,102–109^ 20 reported that women had decreased rates of aging compared to men,^42,61,110–128^ whereas no studies reported that that men were biologically younger or that they aged slower. 11 studies with clocks reported insignificant sex differences,^41,129–138^ and one study did not specify if their results were significant.^139^ Another six studies reported mixed findings. In one of these studies, Beydoun and colleagues^140^ reported significantly accelerated aging on two of the three examined biological clocks, including the Horvath and Hannum extrinsic epigenetic clocks, but not the Horvath intrinsic epigenetic clock. Functional aging index (fBioAge) was examined in one study; men and women did differ in fBioAge, but did not differ in their rate of change.^51^ A study by Levine and Crimmins^31^ demonstrated the impact of chronological age; biological age was significantly lower in women among those aged 20-39 (P = .003), but not those who were older. Tajuddin and researchers^141^ reported that men aged faster biologically according to their measures of universal age acceleration (P = .0051) and extrinsic epigenetic age acceleration (P < .001), but not intrinsic epigenetic age acceleration. Theodoropolou and colleagues^142^ found that intrinsic epigenetic age acceleration and extrinsic epigenetic age acceleration differed by sex (both P < .001), but PhenoAge acceleration residual did not. One study found an interaction effect.,^143^ While sex by itself did not significantly affect biological aging, sex did interact with birth weight; men born at a low birth weight had a significantly older biological age than normal weight men by 4.6 years (P = .01), whereas women did not differ in biological age by their birth weight.

Frailty was examined in three studies with mixed results; women were significantly more frail in one study (P = .001),^60^ and represented a significantly larger proportion of the frail group than men in a different study (P = 0.03)^91^, although were similar in non-frail and pre-frail categories, and no gender difference were reported in the third study.^90^ Two studies reported results with the fBioAge, with women having worse functioning than men in one of these studies (P < .001),^60^ and the other reporting that the average functioning was significantly better for women at age 75 (P < .001),^52^ but no gender difference in the rates of change in functioning.

Gender may also be related to mortality; men were more likely to have died by follow-up in one study (P = .002),^61^ and another study reported that gender was a significant predictor of mortality, but did not specify which gender.^144^

CDKN2A or p16INK4a expression was examined in five studies; one study found a significantly increased expression of CDKN2A among men (P = .04),^145^ whereas the remaining 4 studies did not find a gender difference.^49,50,90,91^ Mitochondrial DNA copy number (mtDNAcn) was assessed in two studies; Vyas and colleagues^146^ reported a significant positive association of increased mtDNAcn among women versus men, and Praveen and colleagues^92^ found that mtDNAcn was significantly higher among men than women for those aged under 60 (p < .05), but no gender effect above this point. MicroRNA (miRNA) expression was examined in one study, where Accardi and colleagues^147^ found that gender did not correlate with miRNA expression level.

Pathai and colleagues^90^ examined a variety of other variables in relation to gender and aging. Most of the assessed variables were not related to gender, including lens density scale, retinal vessel calibre, and endothelial cell parameters, but three of the six retinal nerve fibre layer (RNFL) parameters, including average, inferior, and temporal quadrants, were significantly thinner in men (p = 0.01, p = 0.0008 and p = 0.02, respectively). In contrast, Pathai and colleagues^91^ conducted a similar study, but found significant gender differences in one of the endothelial cell parameters (P = .03), but no difference in lens density, retinal vessel calibre, RNFL thickness (average, superior, inferior, nasal, temporal), and the remaining endothelial cell parameters and frailty status categories. Growth differentiation factor 15 (GDF-15) was examined in one study, in which being male was a predictor of GDF-15 (P = .017).^148^ No gender differences in waking cortisol were found in one study, however women did have a more rapid decline than men (P < .05).^149^ Twenty-two glycan peaks (GPs) were assessed in a study by Yu and colleagues,^150^ in which 10 GPs (FA1, A2, FA2, M5, FA2B, A2G1, FA2[6]BG1, FA2[3]BG1, FA2BG2S1, and FA2BG2S2) were significantly higher in males, 4 GPs (all P<0.001; A2G2, FA2G2, FA2BG2, and FA2G2S1) were higher among women, and eight were similar in men and women. Ultra-weak photon emission (UPE) in one study found no gender difference across the total study population, but when divided by age group they found that men had a larger difference in UPE between younger and older groups than the younger and older female groups.^151^ Various other variables were similarly only examined in one included study; levels of NT-proB-type natriuretic peptide (NT-proBNP) were significantly higher among women (P = .0001),^152^ indicating increased biological aging, soluble urokinase plasminogen activator receptor (suPAR) was increased among women in a different study,^153^ and an examined allele score did not differ by gender in another manuscript.^69^ One study examined the CPG methylation of a variety of genes, and found that the methylation of PDE4C was significantly lower in women than men (P = .024), but no gender differences in aspartoacylase (ASPA) and ITGA2B.^154^

### Race/Ethnicity

A total of 39 papers examined race or ethnicity and made a variety of comparisons. Common datasets of the included studies are: NHANES data in 4 studies,^5,32,33,155^ HRS population in 4,^35–37,156^ Sympathetic Activity and Ambulatory Blood Pressure in Africans study population in 2,^47,48^ and another two with the same data.^49,50^ Across all these manuscripts, 22 found significant race differences in biological indicators of aging, five reported mixing findings, and 12 had insignificant results.

Race was most commonly compared to TL, in a total of 19 studies. Eight of these studies reported that Black people had longer telomeres than other races, including Caucasian people,^5,35–37,65^ Hispanic people,^33^ both Caucasian and Hispanic,^32^ and in general.^62^ One study found significantly shorter telomeres in Caucasian people than non-Caucasian.^55^ In contrast, two studies found that Caucasian people had significantly longer telomeres than Black people (P < .001).^47,48^ None of the studies reported longer telomeres among Hispanic people. Instead, two additional studies found shorter telomeres among Hispanic people compared to non-Hispanics,^46^as well as Black and Caucasian .^56^ However, six studies found insignificant results related to race and TL.^50,54,63,64,83,157^

Fourteen studies compared race to either a calculated biological age or the difference in chronological and biological age. Results of five studies showed that non-White participants had higher calculated biological ages than White participants across all calculations.^105,134,136,137,158^ Tajuddin and colleagues^141^ found no difference between Black people and Caucasians in universal age acceleration and intrinsic epigenetic age acceleration measures, but did find a significant difference in extrinsic epigenetic age acceleration by race (P < .00001). After adjusting for covariates, Levine and Crimmins^155^ reported that Black people had older biological ages than Caucasian participants (P < .001), which equated to a three year difference in biological age. This difference did vary by chronological age, as the difference in biological age increased until ages 60-69 when Black people were found to be 4.82 years older biologically than Caucasians (P < .001), but then steadily declined until there was only a 1.17 year difference (P = .054) for those 80-89 years old. The specific clock used to determine biological age also matters; Hamlat and colleagues^159^ reported a negative correlation between the proportion of Caucasian participants related to DNAm GrimAge acceleration, whereas no race effects were found in relation to Horvath or Hannum DNAm age acceleration. Faul and researchers^120^ found Black individuals had significantly accelerated biological aging compared to white individuals in PhenoAge, GrimAge, and Dunedin, but significnatly decelrated compared to white individuals in Hannum, and no difference in Horvath; Hispanic individuals had significantly accelerated biological aging in Dunedin, significantly decelerated in PhenoAge, and no difference in Hannum, Horvath, and GrimAge. Kresovich and colleagues^160^ found Black participants had significantly accelerated biological aging compared to non-hispanic whites in GrimAge and DunedinPACE, but not PhenoAge. Four studies reported insignificant results related to biological age and race.^102,112,129,138^ CDKN2A/p16INK4a expression was assessed in three studies; one reported a significant association of CDKN2A expression with being Black compared to Caucasian (P=0.031),^145^ whereas two studies had insignificant results.^49,50^ Crimmins^156^ examined both disability and cognitive dysfunction compared to race; in their model involving biological, demographic, and social factors, being Black was related to significantly greater cognitive dysfunction (P = NR), and being either Black or Hispanic was related to a slightly increased rate of disability. Assessed in one study, mtDNAcn did not vary by race.^146^ In terms of cortisol in a study by Samuel and researchers,^149^ Black people had lower waking and higher bedtime cortisol than people of other races (P < .05), which equated to a 26% lower average waking cortisol level and a 1% slower diurnal decline for Black people.

### Socioeconomic Status

A total of 98 studies examined variables relating to socioeconomic status, including 64 which examined education, 51 income or poverty, 12 occupation, 4 home ownership, and five assessed health insurance. Some studies used the same datasets, which include NHANES data in five studies,^5,32,33,103,161^ the FACHS population in four,^3,41,42,162^ the HRS population in three,^35,37,156^ the Netherlands Study of Depression and Anxiety population in two,^38,39^ the Korean Genome Epidemiology Study in two,^43,44^ Dunedin cohort in two,^163,164^ Psychological, Social, and Biological Determinants of Ill Health (PSOBID) Cohort in two,^96,165^ the Korea National Health and Nutrition Examination Surveys data in two,^103,161^ and another two with the same data.^49,50^

### Income and Poverty

In three studies, income was significantly and positively related to TL,^69,74,99^ and these variables were inversely related in one study (P = .035).^44^ Higher income was significantly related to decelerated DNAm PhenoAge in three studies, ^106,125,159^ lower calculated biological age in three,^3,103,162^ and decelerated GrimAge in one (P ≤ .01).^42^ Similarly, decreased income was significantly related to accelerated DNA methylation phenotypic age in one study (P < .0001),^166^ quicker telomere attrition (p = 0.024),^96^ and accelerated biological aging in three studies (Arpawong, 2023; BeachSRH, 2022).^41,104,167^ Income level was not related to TL in 12 studies,^35,37,43,45,63,64,81,84,89,93,101,165^ Horvath DNAm Age in two studies,^118,159^ Hannum DNAm Age in 2,^159,166^ DNAm GrimAge in three,^119,138,159^ and allele score in one.^69^

Lower household wealth predicted significantly longer TL compared to those with high household wealth in one study (P < .05).^66^ Similarly, higher financial hardship was significantly related to accelerated aging in two studies.^3,168^ Poverty was not related to TL in one study,^78^ accelerated biological aging in three,^124,141,162^ and both intrinsic and extrinsic epigenetic aging in one study.^141^ Financial stress was not correlated with accelerated aging.^41^

Income to poverty ratio in relation to TL was assessed in two studies with conflicting results; one found these variables to be positively related (P < .01),^169^ whereas the other reported insignificant findings.^32^ Income below the poverty line was associated with higher Dunedin PACE in one study.^170^ Socioeconomic status (SES) was reported in seven studies; higher SES was significantly related to having a decreased biological age in two studies,^128,161^ but not related to telomere in two studies.^68,164^ Various other variables related to socioeconomic were also examined. First, socioeconomic deprivation was significantly correlated with both TL (P = .036) and global DNA methylation (P = .019).^165^ Increases in socioeconomic stress were associated with an increased speed of epigenetic aging using GrimAge, PhenoAge, and DunedinPoAm in one study.^171^ Socioeconomic vulnerability was not related to waking cortisol levels after controlling for covariates (P > .05), but was associated with a select differences in diurnal cortisol decline (p < .05).^149^ One study examined life course social class trajectory which they categorized into four groups based on childhood and adulthood social class; the participants who had a low childhood and adulthood social class were 2.42 years biologically older according to a GrimAge calculation than those with a both high childhood and adulthood social class.^172^ Finally, cumulative social disadvantage was significantly positively correlated with accelerated GrimAge and Pace of Aging.^42,136,173,174^

### Education

A total of 64 studies examined education, either as a categorical or continuous variable. TL was used as a proxy for biological aging in 35 studies with a moderate degree of consistency in results; 24 of these studies found that education was not related to TL.35,37–39,45,50,61,62,66,71,72,82,84,87,89,93,95,96,99,157,169,175–177 Seven studies reported that more education was predictive of longer telomeres (all P < .05),^32,33,57,64,69,81,178^ and one did not report the direction of their significant results.^5^ The remaining three studies reported mixed results.

Geronimus and colleagues^54^ analyzed eight statistical models, in which education was related to TL in one model, with individuals who reporting having less than a high school education having shorter telomeres than college graduates (p ≤ .05). Two studies found a conflicting gender interaction effect.^63,101^ In the first, while education did not effect TL in the whole sample, analyses divided by gender indicated that educational attainment (high school or less) was inversely associated with TL among men after adjustment for covariates (P=0.03), which was not found for women.^63^ The other study found that TL and education were significantly positively associated for women (Ptrend= 0.001), but not men (Ptrend= 0.698).^101^ One of these studies also indicated that the potential impact of analysis methods, as years of education as a continuous variable was positively associated with longer telomeres (Ptrend= 0.003), but was unrelated when education was dichotomized.^101^

Education was compared to a calculated pace of aging in 21 studies, with over half of these studies (11 studies) reporting significant benefit to higher education across a variety of clocks.^42,103,104,126,127,136,160,163,179–181^ For example, one of these studies reported that individuals with less education had increased Horvath, Hannum, and Levine EAA, with further analyses indicating a dose-response effect.^179^ This relationship equated to a difference in biological and chronological aging of approximately 1-2 years between those of higher education compared to those with less education in one study,^180^ and a difference of nine years in DNAm age between people with post-baccalaureate vs. lower levels of education in another study.^181^ In addition to pace of aging, increased educational attainment also predicted a decreased facial age (P < .01).^163^ George and researchers^123^ found that lower educational attainment was associated with greater PhenoAge and GrimAge but not Horvath or Hannum. Nine studies reported an insignificant relationship between education and rate of biological aging.^3,41,119,135,138,159,162,168,182^ Five of the studies examining both education and pace of aging reported mixed results.^61,111,114,116,161^ Two of these studies reported difference in the clocks used,^111,116^ with both finding that increased education was related to a decreased rate of age acceleration via the Hannum model (P = .04, P < .001; respectively), but insignificant results according to the Horvath age acceleration model. Faul^120^ found that greater education was associated with accelerated biological aging in GrimAge and Dunedin clocks, but not Horvath, Hannum, or PhenoAge. Han and colleagues^161^ reported that the relationship between education and biological aging varied by gender; education level was significantly associated with both biological age and the difference in biological and chronological age for women (both P < .0001), but the equivalent was insignificant for men. One study assessed this relationship in multiple datasets, with significant benefits of education found in one of four datasets (P = .01).^114^ McLachlan and researchers^61^ examined 4 different aging indicators, in which years of education was negatively correlated with accelerated DNA methylation GrimAge (P = .001), but was not associated with brain-PAD or mortality. Mortality risk was also not related to education in another study of older adult male Caucasian veterans.^183^ Two studies using the same data examined CDKN2A/p16INK4a expression and found that this was not related to educational status.^49,50^ In terms of cortisol, reporting some college or trade school education was associated with approximately 17% lower average waking cortisol (P < .05) than individuals who with a college education or greater.^149^ Education did not differ by allele score.^69^

### Employment and occupation

A comparison between being employed versus not resulted in conflicting findings. Being employed was related to being significantly biologically older in one study by 1.65 years (P = .049),^162^ whereas Han and colleagues^103^ reported that being employed was associated with being significantly younger than their peers (P < .0001). Kingma and researchers^81^ found that work situation was significantly correlated with TL, in that TL increased between being unwillingly unemployed, willingly unemployed, and employed (P = .002). Employment status was not related to TL in two studies,^101,169^ although subsample analyses of one of these studies revealed that male caregivers had significantly longer telomeres than those employed (P < .001), which was the opposite for women (P = .021).^101^ In addition, time spent working per week and being on a shift work schedule were unrelated to TL.^184^

Type of occupation was not related to biological indicators of aging in the majority of studies. Non-significant differences in TL between occupational grade (e.g., manual labour versus non-manual work) was reported in four studies.^53,96,168,178^ This relationship varied by biological aging indicator in a study by Marioni and colleagues;^114^ Hannum biological age significantly differed by occupational class in one if the two examined cohorts (P = .0029), but not Horvath biological age in either cohort. In another study, being classified as non-manual labour occupational class was correlated with both accelerated grimAge (P < .001) and death by follow-up (P = .0001), but not TL or brain-PAD.^61^

### Other socioeconomic variables

Home ownership was assessed in four studies. Shiels and colleagues^96^ reported that telomere attrition occurred at a faster rate among tenants than homeowners (P = .038). Similarly, Carroll and researchers^45^ found that homeowners had significantly longer telomeres than their peers after adjusting for covariates (P < .001); further analyses indicated that race plays a role, as this relationship present for Caucasians and Hispanics (P = .03; P < .001, respectively), but not for Black people. Home ownership and TL varied by chronological age in another study; renters were biologically older than homeowners by 9.6 years (P = .046) among their cohort of 35-year older, but this was insignificant for their cohort of 55-year old.^101^ Ng and colleagues^180^ categorized housing into public housing, private apartments or condominiums, and property with land; the difference in biological and chronological age significantly varied by housing (P < .001), with those with private housing having the lowest biological age compared to their chronological age. One study reported no differences in calculated biological age by housing status.^168^ Rural versus urban housing was not predictive of telomere.^66^

Having health insurance was examined in two studies with similar populations but conflicting findings; having health insurance was significantly negatively correlated with accelerated biological aging among Black women (P ≤ .05),^162^ but was unrelated in a different study comprised of Black people.^41^ Three additional studies tested the relationship between biological age and health insurance coverage, reporting insignificant results.^74,119,136^

### Region

Eight studies assessed the impact of region, each with a different specific variable. Living in a rural versus urban location was not predictive of TL in three studies.^66,74,107^, but was associated with shorter telomeres in one.^122^ An and colleagues^35^ examined Northeast, Midwest, South, and West United States, but did not find a difference in TL. Similarly, physiological dysregulation state did not differ between East Europe (i.e., Czech Republic, Poland) and West Europe (i.e., Austria, Germany). In terms of country of Birth (Australia/New Zealand, United Kingdom/Malta, Italy, or Greece), Greek nationality was associated with accelerated aging with the Horvath model (P = .01).^111^ Rapp and colleagues^185^ found that the United States, Italy and Israel exhibit elevated biological age relative to chronological age, while the Netherlands, Sweden, Greece and Switzerland exhibit delayed biological aging.

### Built Environment and Population

Built environment and biological age was examined in one study, which reported mixed results. Zhao and researchers^74^ (2021) found that higher land use mixture was associated with longer TL, but no other built environment or population density were significantly associated after controlling for covariates.

### Sexuality

Sexuality was examined in one study, which compared men who have sex with men vs men who do not have sex with men.^102^ The researchers did not find any significant difference in biological age between these groups.

### Mental Health and Substance Use Indicators

A total of 151 of the included studies examined a mental health or substance use variable in relation to biological aging. Dataset overlap within this subsection occurred in a total of 59 studies, including seven studies using data from the HRS,^35,36,105,120,156,186,187^ five using data from the Dunedin cohort,^153,163,164,188,189^ four with NHANES data,^10,31–33^ four with the FACHS,^41,42,167,171^ three with the Korean Genome Epidemiology Study population,^43,44,190^ and two studies for each of the following datasets: Netherlands Study of Depression and Anxiety,^38,39^ Multiethnic Study of Atherosclerosis (MESA),^45,46^ Korean KNHANES study,^103,161^ Lothian Birth Cohort,^61,191^ Prevention of Renal and Vascular End-stage Disease (PREVEND) study cohort,^81,192^ PSOBID Cohort,^96,165^ VITamin D and OmegA-3 TriaL-Depression Endpoint Prevention,^146,181^ Sympathetic Activity and Ambulatory Blood Pressure in Africans,^47,48^ Helsinki Businessman Study population,^7,193^ MIDUS National Survey,^194,195^ Moli-Sani Study,^168,196^ China Kadoorie Biobank,^107,197^ UK Biobank,^198,199^ Vietnam Era Twin Registry,^200,201^ National Health and Resilience in Veterans,^176,202^ data with geriatric caregivers,^98,184^ Finnish twins,^109,203^ and two with the same population of heterosexual adults.^49,50^

### Overall Mental Health

Four studies examined an overall measure of mental health flourishing or dysfunction.^99,156,175,176^ In a study of former Swiss indentured child labourers (’Verdingkinder’), mental functioning was not associated with TL or chronological age.^175^ Another study focused on older adults and found that adverse mental states (including negative psychological outlook, previously being depressed, and previous chronic stressors), were related to worse health outcomes, including difficulties with daily life activities, multimorbidity, cognitive dysfunction, and mortality.^156^ Two studies focused on psychological distress; this was not a significant predictor of TL in any of the examined statistical models in a study of older adults.^99^ Watkins and colleagues^176^ conceptualized psychological distress as including PTSD, depressive, and anxiety symptoms; psychological distress among military veterans did not reach significance in predicting TL (P = .09). One study examined having a mental illness as a covariate in their model predicting TL; this was an insignificant predictor (P = .445).^53^ Researchers from one study reported that resilience was not associated with GrimAge.^202^

### Depression

Depression or depressive symptoms were examined in 34 studies (Arts et al., 2018; Brouwers et al., 2016 Chae et al., 2016 Cruz-Almeida et al., 2019 D’Aquila et al., 2019 D’Aquila et al., 2017 Epel et al. 2013 Fang et al. 2021 Hassett et al., 2012 Hoen et al., 2013 Kuffer et al., 2016 Lee et al., 2017 McLachlan et al., 2020 Robinson et al., 2020 Ryan et al., 2020 Shalev et al., 2014 Verhoeven et al., 2014 Vincent et al., 2017 von Kanel et al., 2015 Vyas et al., 2019 Vyas et al., 2020 Watkins et al., 2016 Wium-Andersen et al., 2017 Yen et al., 2013 Zhao et al., 2016a Depressive symptoms]; Beydoun 2022 Brown 2022 Faul 2023 Forrester 2022 (#283)Hautekiet 2022 Katz 2022 Liu 2022 Loh 2023 Wagenmakers 2023).^39,40,48,61,69,72,78,95,99,129,146,164,166,169,175,176,181,186,192,204–211A^ total of eight articles reported significant results, (Arts et al., 2018; Fang et al. 2021; Hassett et al., 2012; Verhoeven et al., 2014; Wium-Andersen et al., 2017; Zhao et al., 2016a [Depressive symptoms are]; Forrester 2022 (#283); Liu 2022) including five articles reported a significant relationship between depression symptoms and TL,^39,69,72,78,209^ with shorter TL being related to increased depressive symptoms. Shorter TL was also significantly related to being admitted to a hospital for depressive symptoms (P = .01),^69^ but not the age of onset of depressive symptoms (P = .216).^39^ Depression and frailty were compared in one study with significant results; frail participants reported significantly more severe depressive symptoms and were more likely to report having a depressive disorder (P < .001) than non-frail participants.^204^ One study reported that being at risk for depression was associated with a 0.05 year increase in biological age acceleration long-term,^212^ and another study indicated that individuals reporting more depressive symptoms were associated with increased epigenetic age acceleration according to HannumAA (P = 0.019) and EEAA (P = 0.012) at baseline, and PhenoAA (P = 0.027) at follow-up.(Liu 2022) Five studies reported mixed findings. McLachlan and colleagues^61^ reported that depressive symptoms were significantly correlated with accelerated DNA methylation GrimAge (P = .021) and brain-PAD (P = .032), but not TL. In a study by Robinson and colleagues,^166^ having both minimal and severe depressive symptoms was associated with metabolic age acceleration (P = .01; P = .001, respectively), but depression was unrelated to age according to Horvath’s Pan-Tissue clock, Hannum’s epigenetic clock, and Levine’s phenotypic age clock.

Another study found that depression associated with telomere shortening in men (P = .003), but not in women.^164^ Beydoun and colleagues^105^ reported that the statistical significance between the trajectory of depressive symptoms biological clocks varied by clock, sex, and race, and comparison. Another study reported a significant relationship between previous depression and some epigenetic age calculators including Hannum and GrimAge (P < 0.01), DunedinPACE (P < 0.001), but not Horvath or PhenoAge (Faul et al., 2023). Mastrobattista^211^ compared depressive symptoms to GDF-15 levels; these variables were not significantly related, but results indicated that individuals with later onset depression had higher levels of GDF-15.

In contrast, 21 studies found an insignificant relationship between depression or depressive symptoms and the biological indicators.^43,48,73,95,99,129,146,169,175,176,181,192,205–208,210,213–216^ Results from these studies indicate that depression and depressive symptoms were not related to: specific CpG methylation sites,^206,207^ frailty,^213^ mtDNAcn,^146,181^ DNA methylation,^181^ biological clocks,^129,181,215,216^ and TL in nine studies.^48,99,169,175,176,181,192,208,210^ In addition to these findings, Ryan and colleagues^95^ reported that TL did not differ between individuals with severe depression and controls, was unrelated to the number of reported depressive episodes experiences and the duration of depression, and did not differ between individuals with unipolar versus bipolar depression, nor psychotic versus non-psychotic depression. Authors of one of the studies also reported that depressive symptoms significantly interacted with racial discrimination in predicting TL (P = .020);^169^ among individuals not reporting racial discrimination, TL was longest among individuals with less severe depressive symptoms, whereas for those reporting substantial racial discrimination, TL was longest among those with severe depressive symptoms.

### Anxiety

A total of 11 studies examined the relationship of anxiety with a biological indicator of aging, including six studies which examined anxiety symptoms,^61,78,166,169,181,209^ and five studies which assessed anxiety disorders.^39,164,192,214,217^

Five studies compared anxiety symptoms to TL, all of which found an insignificant correlation between these variables.^61,78,169,181,209^ However, McLachlan and colleagues^61^ also compared anxiety symptoms to other indicators of biological aging, and reported that anxiety symptoms were significantly correlated with accelerated DNA methylation GrimAge (P = .002) and brain-PAD (P = .011). One study assessed anxiety symptoms of policy officers in relation to metabolomic age acceleration,^166^ with results indicating this significantly increased with anxiety symptoms, with a 0.32 years of age acceleration among those with elevated anxiety symptoms compared to individuals without (P = .05). Another study comprised of middle-aged and elderly adults reported that anxiety symptoms were significantly positively correlated with epigenetic age acceleration, extrinsic epigenetic age acceleration, and Horvath’s intrinsic epigenetic age acceleration (all p < .05), but not DNA methylation age, mtDNAcn, or Hannum’s intrinsic epigenetic age acceleration.^181^

Four studies examined the relationship between anxiety disorders and TL (Heutekiet et al., 2022),^39,164,192,214^ with mixed results. Hoen and colleagues^192^ found that having an anxiety disorder predicted shorter TL (P = .022), after adjusting for age and gender. Further analyses indicated that of the examined anxiety-related disorders, panic disorder, agoraphobia and social phobia all significantly predicted telomere shortening (P = .006), but generalized anxiety disorder (GAD) did not. In a study divided by gender, men demonstrated TL shortening among those with generalized anxiety disorder (GAD; P =.015), whereas this effect was insignificant among women with GAD.^164^ Hautekiet and colleagues^214^ found that GAD was not significantly associated with TL or mitochondrial DNA content. Of the internalizing disorders examined by these researchers, individuals with GAD had the shortest TL. Verhoeven and researchers^39^ studied a sample consisting of individuals with remitted and current major depressive disorder (MDD) as well as controls. They found that comorbid anxiety disorder was significantly more frequent among those with current MDD. However, the relationship between comorbid anxiety disorder and TL among those with current MDD was not significant (P = .057). Martínez de Toda and colleagues^196^ examined biological aging in women with anxiety compared to healthy centenarians and found that women with anxiety were showed an increased biological age compared to their chronological age, whereas healthy centenarians showed the opposite.

### Stress

In total, nine studies examined stress.^49,50,103,161,178,202,208,209,218^ All 5 studies which examined TL found an insignificant relationship between TL and measures of stress, including four studies which included solely perceived stress,^178,208,209,218^ and one study which examined perceived stress, chronic stress, and accumulated daily stress.^50^

Three studies compared a calculated biological age with stress,^103,161,202^ of which two used the same dataset; all three articles reported no difference in biological age related to stress, but one of these studies did find that perceived stress was significantly associated with having a disproportionately high biological age compared to chronological age (P = .0044).^103^

Two studies reported the relationship between stress and the expression of the p16INK4a-encoding gene CDKN2A.^49,50^ Rentscher and colleagues^49^ reported that CDKN2A expression was significantly positively correlated with chronic stress exposure, perceived stress (both p < .01), and accumulated daily stress (p < .05). In a different study, Rentscher and researchers^50^ found contrasting results in terms of the relationship between measures of stress and biological age indicators; increased chronic stress exposure, perceived stress, and accumulated daily stress predicted elevated p16INK4a expression (gene CDKN2A), with each responsible for 8%, 14%, and 8% of the variance in p16INK4a, respectively. Further analyses indicated that particular facets of chronic stress were particularly important to p16INK4a expression, including marital conflict and work-related stress. However, all three stress outcomes in this study were not significant in random effects models predicting TL.

### Post-Traumatic Stress Disorder

Of the ten studies examining PTSD and aging, five reported significant findings as follows. In a study compared former indentured child laborers both with and without PTSD versus controls, former indentured child laborers with PTSD had significantly longer telomeres than the control group (P = .04).^175^ O’Donovan and colleagues^87^ assessed a model adjusted for chronological age, and found that individuals with PTSD had significantly shorter telomeres than those without (P = .03), but TL did not differ by symptom severity among those with PTSD. Among police officers, significant increases in the rate of aging were found for both individuals with PTSD who experienced trauma in the last six months according to their metabolomic age (P = .02), as well as individuals who experienced trauma without PTSD according to their phenotypic age (P = .029).^166^ In a twin study, researchers found that twins with PTSD had increased biological age acceleration compared to twins without PTSD.^201^ Yang and colleagues^219^ reported significantly increased biological age acceleration amongst individuals with PTSD compared to controls in all cohorts (p = 0.008) and a positive correlation between PTSD symptom severity and biological age acceleration (r = 0.39, p = 0.049). In contrast, TL was not significantly associated with PTSD in two studies,^164,176^ of which one examined military veterans.^176^

Two studies reported mixed findings, one of which found that PTSD predicted GrimAge epigenetic age acceleration in the younger (Mage = 32) but not the older (Mage = 52) cohort.^220^ The other found that individuals with PTSD had increased GrimAge acceleration compared to controls (p = 8.77e-9), and no change in GrimAge acceleration was indicated post-treatment of PTSD. Researchers from one study reported that lifetime PTSD or MDD was not significantly associated with GrimAge.^202^

### Other Mental Health Disorders

Schizophrenia was assessed in relation to aging in two studies.^70,221^ Dada and colleagues^221^ reported the association of various measures of biological aging as well as symptoms related to schizophrenia (paranoia, psychosis, reality distortion, disorganization, depression, negative symptoms, composite score). Significant correlations between these include scores on the composite measure with age acceleration on the Hannum clock (P = .019), psychosis with Horvath age acceleration (P = .029), EEAA (P = .014), and Hannum age acceleration (P = .017), reality distortion with Hannum intrinsic epigenetic age acceleration (P = .044), and disorganization with extrinsic epigenetic age acceleration (P = .039). Wolkowitz and colleagues^70^ studied both individuals with schizophrenia and controls; there was no effect of schizophrenia diagnosis on TL, however, there was a significant interaction effect of gender and diagnosis (P = .016), with women with schizophrenia having significantly shorter telomeres than healthy women.

Okazaki and colleagues^222^ examined biological aging in individuals with autism spectrum disorder (ASD) and found no significant difference in biological age acceleration between the group with ASD and the control group.

### Experiences of Abuse, Trauma, and Adaptation in Adulthood

Three studies reported the relationship between abuse and aging.^67,78,223^ Savolainen and colleagues^67^ examined four abuse related variables: emotional abuse, emotional neglect, physical abuse, and reporting experiences a threat to life, pain, or bizarre punishment. Overall, reports of lifetime abuse were not significantly related to TL. However, when the sample was divided into those who were or who were not separated from their family during World War II, individuals who experienced both childhood separation and abuse had shorter TL than non-separated and non-abused individuals (P = .012). In a study of abused and non-abused older adults with mild to moderate cognitive impairment,^78^ all forms of examined abuse were significantly correlated with shorter TL, including psychological abuse (P = .005), physical abuse (P = .028), and reporting multiple forms of abuse (P = .003). Mason and colleagues^223^ examined the impacts of various levels of physical and sexual abuse (none, mild, moderate, and severe) on young nurses. While TL did not significantly change between these categorizations, findings may still be of clinical importance; for example, individuals who reported moderate physical abuse had shorter telomeres than individuals who reported no abuse by an amount equivalent to 5.3 years of age.

Trauma and adverse life events were examined in six studies.^156,176,186,192,224,225^ Watkins and colleagues^176^ reported that the number of lifetime traumatic events experienced (e.g., assault during childhood, military-related trauma, death of loved one) among military veterans did not significantly differ between individuals with normal versus shorter telomeres. Two studies used trauma and adverse life events as a covariate in their respective models predicting TL,^186,192^ but this was not a significant covariate. One study examined multimorbidity; adulthood trauma (e.g., experienced natural disaster, victim of assault) was weakly associated with multimorbidity (P = NR).^156^ Barha colleagues^224^ assessed experiencing child mortality and reported that this was significantly associated with shorter TL after adjusting for a confounder (P = .015); further analyses indicated a mediation effect of HPAA activity on this relationship. One study found that individuals who experienced severe exposure to certain adverse life events (e.g. intense bombing, wartime exposure, witnessing death firsthand) exhibited higher frailty than those who did not face exposure as severe.^225^

One study examined adaption following a traumatic event by comparing the TL of individuals who reported adapting quickly or slowly; life event adaptation following a traumatic event was not related to TL.^178^

### Smoking

Smoking was examined in a total of 96 studies. Of the studies assessing smoking, 78 compared smoking and aging by categories, the most common was comparing any level of smoking to never smoking, which was examined in 31 studies. This was compared to TL in 18 studies, of which five studies reported significantly longer telomeres among non-smokers,^44,78,96,165,193^ whereas 12 reported insignificant results.^35,63,85,95,169,190,210,218,226–229^ One study had mixed findings, as the effect of smoking varied by health status: TL was significantly inversely associated with smoking for non-coronary artery disease patients (P = .039), but these variables were not associated in patients with premature coronary artery disease.^97^ Smoking was also compared to mortality risk in three studies; these studies consistently reported a significant relationship between smoking and an increased mortality risk,^152,183,193^ of which one reported that current smokers had over a 3-fold increase in mortality compared to non-smokers over a 7-year follow-up period.^193^ Six studies examined a calculation of biological age, but results were divided by their methodology; two of the three that compared smoking to a calculated biological age (i.e., GrimAge and a novel clock) reported smokers were significantly biologically older relative to their chronological age compared to non-smokers by 1.25, 2.3, and 7.1 years,^122,230,231^ and one reported that ever smoking was unrelated to several clocks (i.e., Hannum, Horvath, Levine, DNA Methylation-based BA),^232^ whereas studies that examined age acceleration (i.e., Horvath EAA, Hannum EAA) found only insignificant results related to smoking.^162,182,233^ One study using the 7-CpG clock reported no significant differences in DNA Methylation age acceleration between groups stratified by smoking.^234^ Two studies reported IL6 and found smoking was not related to this variable.^96,165^ Nine more variables were compared to smoking in one study respectively, with the following results: smoking was related to significantly greater levels of suPAR (P <.0001),^153^ serum Pi levels (P = NR),^165^ adiponectin (p=0.046),^165^ D-dimers (p=0.046),^165^ and decreased pulse wave velocity (P = .001),^58^ Augmentation index (Aix) (P = .002),^58^ eGFR (P = .046),^165^ and global DNA Methylation (P = .016).^165^ Smoking was unrelated to metabolic age score,^235^ physiological dysregulation,^236^ Vitamin D levels,^165^ and cholesterol.^165^

Thirty-one studies categorized smoking into three distinct groups: current smokers, ex-smokers, and never-smokers. Twelve studies compared these categories to TL with varied results by comparison; three studies reported that TL was longest among never smokers compared to both current smokers and ex-smokers,^38,39,69^ respectively, and another reported that longer telomeres were more common among never and current smokers compared to ever smokers (P = .048).^32^ Surtees and colleagues^178^ found that current smokers had significantly shorter telomeres than never smokers (P = .03), whereas ex-smokers and never smokers did not differ. In comparison, seven studies noted only insignificant relationships between smoking categories and TL.^54,61,71,76,186,237,238^ Of the 17 studies which compared current, never, and ex-smokers to a calculated biological age or age acceleration, 11 reported significant results,^31,61,103,147,161,172,179,199,239–241^ four insignificant results,41,102,138,242 and two mixed results.^116,166^ Divided by calculation methodology, Horvath EAA did not differ between the comparisons of the three smoking groups in two studies.^179,242^ In contrast, Horvath EAA varied by smoking status in two studies: current smokers had significantly younger Horvath DNA methylation ages in one study (P = .015),^166^ and the other noted that current smokers reported significantly increased Horvath and Hannum epigenetic age acceleration compared to never smokers (both P <0.001), but former smokers did not differ from current smokers.^116^ Three studies examined Hannum EAA: current smokers benefitted over other categories in two studies,^116,179^ but categories did not differ in another.^166^ GrimAge significantly differed in two studies (both P < .001),^61,172^ with one study indicating that current smokers were 7.91 years biologically older.^172^ Of the six studies that subtracted chronological age from biological age, no difference was detected between the smoking categories in two study,^102,138^ three found that non-smokers were biologically younger,^103,161,240^ and another study found that while current smokers were biologically older than never smokers in both populations examined (P = 6.9e-10; P = 1.0e-50),^239^ this biological age acceleration was reversed back to normal for those who quit smoking. One study used the ratio of predicted age to chronological age (AgingAccel) and found that compared to smoking never smokers had significantly slower AgingAccel (P <0.001) and aging rate (p =0.009).^241^ Another study found that smoking status significantly differed between Dietary Inflammatory Score (DIS) and Energy-adjusted Dietary Inflammatory Index quartiles, though smoking was not significantly associated with accelerated aging upon the analysis of DIS (Martinez et al., 2023). Another five studies took unique approaches to calculating biological age, with results as follows: former and current smokers had significantly increased Levine EAA (both p < 0.001),^179^ smoking was significantly related to accelerated biological aging^31^ and brain aging via brain-PAD (P = .014),^61^ but categories did not differ according to metabolomic age or the DNA methylation phenotypic age,^166^ and mRNA-based BA was not associated with smoking status.^41^ Other variables were compared less frequently to these three smoking categories. For example, three studies examined frailty with conflicting findings; two studies noted that current smokers were more commonly frail,^191,199^ whereas another found them unrelated.^71^ Mortality was significantly correlated with smoking (P < .001),^61^ ever smoking had a significant correlation with miR-126-3p and miR-181a-5p levels,^147^ smoking significantly affected disease status among males (P<0.001) and females (P = 0.048),^107^ allele score did not differ by smoking,^69^ and ferritin levels were more likely to be high among ever smokers and low among non-smokers.^32^

Ten studies examined daily smoking, of which seven studies treated this as a dichotomous variable, and three studies further categorized daily smoking by frequency or amount. Of the ten studies, three study reported that smoking and significantly related to a biological indicator of aging, as smoking was associated with a mortality in that smoking was related to 3.8 years reduction in lifespan (P = 6e−15).^144^ smoking daily during adolescence was associated with an increased biological age at midlife,^189^ and tobacco smoking was correlated with pace of aging (P < 0.001).^188^ Seven studies reported insignificant results, with smoking unrelated to TL,^48,50,53,72,226^ and CDKN2A expression.^49,50^ One study reported mixed findings: smoking status was significantly positively correlated with levels of fibrinogen (P = .01), but was not correlated with von Willebrand factor antigen (VWF:Ag) (P .518), D-dimer (P = .615), or plasminogen activator inhibitor-1 antigen (PAI-1:Ag) (P = .4).^47^

The final conceptualization of smoking categories compared non-smokers to smokers (current and/or past) via the number of cigarettes per day. Within these seven studies, increased age acceleration was detected in current and former smokers against never smokers, in which current smokers 20 or les cigarettes a day were 2.12 years older biologically (P < .001) and those who smoked more than 20 were on average 1.26 years older biologically (P = .01).^111^ One study reported accelerated aging (Δage>0) was significantly associated with current smoking (P < 0.001) and insignificantly associated with previous smoking.^168^ Ng and colleagues^180^ reported that smokers had significantly older biological ages relative to their chronological age, with current smokers the oldest (P < .001). Another study reported that moderate or heavy smoking was associated with worsening frailty status, wherein smoking 15-24 cigarettes daily was associated with increased risk of worsening frailty status in prefrail individuals.^197^ Three studies using this conceptualization found that smoking was unrelated to TL (Iannarelli et al., 2022).^43,81^

Twenty studies examined smoking without categorizing people into smoking categories. A calculation of pack-years was assessed in a total of 13 studies with assorted results. Increased pack years was significantly related to an increased pace of aging and facial age (both P < .01),^163^ and shorter telomeres,^10,45,46,98,243^ but one of these studies found that there was no difference between non-smokers and pack years under 10,^45^ one found this effect only in women (p < .001),^36^ and others found this relationship insignificant.^33,181^ In terms of biological clocks, pack years was significantly associated with the Levine clock (P = .002),^232^ GrimAge (P < 0.001), PhenoAge (P < 0.001), and DunedinPACE (P < 0.001), and Horvath (P < 0.01)^120^ in some studies. In other studies, the results were insignificant for the Hannum,^116,232^ Horvath,^116,181,232^ or epigenetic age acceleration.^116^ Using the principle components of five biological clocks, Faul and colleagues (2023) found that pack years was significantly related to the Horvath (P < 0.05), PhenoAge, GrimAge, DunedinPACE (P < 0.001), but not Hannum. One study looked at several clocks;^244^ pack-years was significantly associated with increased GrimAge (P < .0001), Zhang acceleration (P < .0001), Yang (P < .01), related only for portions of their examined populations for PhenoAge, MiAge, Lin deceleration, Weidner, and Garagnani clocks, and was not related to Horvath Horvath EAA, Horvath2 EAA, Hannum EAA, Vidal-Bralo, or Bocklandt clocks. This study also found that smoking was significantly associated with elevated GDF-15 (P < .0001) and elevated PAI-1 (p<0.01), whereas results varied by dataset in terms of Adrenomedullin (ADM), Cystatin C, TIMP-1, Beta-2 microglobin, NK cell levels, and granulocyte levels, and pack-years was unrelated to levels of leptin, CD8T cells, plasmablast, CD4T cells.^244^ Pack-years was not related to mtDNAcn in one study,^181^ whereas the other found that this relationship was significant for men but not women, and for Black participants but non-Hispanic White or Hispanic adults.^146^ Further, pack-tears was significantly related to lower DHEA levels (p=0.003),^243^ but was not associated with GH levels^243^ or DNA methylation.^232^

Other studies used a more unique approach of examining smoking. Four of these studies assessed smoking via methylation status at cg05575921; two reported that the impact of smoking on biological age increased linearly with increased exposure,^167,245^ another reported that this was significantly related to an increased mortality risk (P < .05).^183^ A study by Lei and colleagues^246^ reported that smokers were on average biologically older by 4.29 years and 2.15 years via the Hannum and Horvath clocks, respectively. Following smoking cessation as indicated by a reversal of the methylation level of cg05575921, they found a significant decrease in Hannum’s methylomic age (p < .001) and Horvath’s methylomic age (p ≤ .05). One study reported on the number of cigarettes consumed; one found this unrelated to frailty.^204^ Lu and colleagues^247^ examined second hand smoke exposure, and found that with severe second-hand smoke exposure, TL shortens more rapidly; individuals exposed to 20 or more hours of second-hand smoke exposure had significantly shorter telomeres than those without any exposure after controlling for covariates (P = .01) and those with moderate exposure (1-19 hours; P = .005).

### Alcohol consumption

A total of 59 studies examined alcohol in relation to biological aging, of which 22 had dataset overlap. Thirty-six studies conceptualized alcohol into categories of consumption, either by occurrence (e.g., current drinkers, never drinkers), by frequency (e.g., never, once per week, 2-5 drinks per week, or 5+ drinks per week), or by amount (e.g., mL consumed). Increased drinking, or being a drinker compared to a non-drinker, or a heavy drinker compared to a non-drinker, was significantly associated with an increased biological age in four studies,^42,161,180,181^ decreased biological age in one study,^103^ shorter telomeres in five,^38,43,44,58,198^ an increased age acceleration in four,^103,161,166,179^ and frailty in one.^199^ Two of these studies did report a benefit to low-level consumption of alcohol, as those in this category had a decreased proportion of short telomeres (p=0.004)^58^ and biological age acceleration.^168^ Additionally, alcohol consumption was predictive of cardiovascular disease status in men (p < 0.001) and women (p = 0.048),^107^ aortic systolic blood pressure (P = .007), aortic diastolic BP (P = .001), aortic pulse pressure (P = .049), and pulse wave velocity (P = .025),^58^ increase ferritin,^32^ PAI-1,^244^ and ADM. In contrast, 19 studies reported insignificant results, in which alcohol consumption was unrelated to TL,^32,33,50,72,181,190^ age acceleration,^102,111,116,220,234^ biological age,^138,174,230,241^ suPAR levels,^153^ CDKN2A/p16(INK4a) expression,^49,50^ mtDNAcn,^146,181^ stochastic epigenetic mutations,^179^ and mortality risk.^183^ Another 5 studies reported mixed results with alcohol categories, with results differing by specific measurement, cohort, and gender. Two of these studies noted that alcohol consumption was only associated with some of the clocks they examined.^232,244^ Strandberg and colleagues^7^ reported a difference by cohort, in which one cohort had no significant relationship between TL and proportion of short telomeres with alcohol consumption, whereas for the other cohort, TL was significantly related to alcohol consumption. Kankaanpää and researchers^109^ found that greater alcohol use was only associated with accelerated AAPheno in older twins, but all age groups were associated with accelerated AAGrim. One study reported a gender effect; a general aging acceleration was detected for those who drank three or more drinks almost every day (P = .003), but when divided by sex this relationship was only significant for men.^180^

Six studies assessed average consumption of alcohol. Findings from these studies indicated that average consumption of alcohol was positively associated with frailty^191^ and unrelated to mortality.^61^ McLachlan and colleagues^61^ reported that the number of days alcohol was consumed per week was correlated with accelerated DNA methylation GrimAge (P = .004) and accelerated brain-PAD (P = .001). Boström and colleagues^248^ reported a positive association between weekly average consumption and PhenoAge and GrimAge (both P < 0.001) acceleration, but found all other biological age measures insignificant. One study reported an insignificant association between weekly consumption and biological age, but a positive association between lifetime average consumption and accelerated biological aging (P = 0.02).^249^ In terms of TL, two studies reported insignificant findings;^61,238^ in contrast, one study assessed several analyses, and found that alcohol consumption was related to TL only when controlling for all other assessed covariates.^69^

Thirteen studies reported on excessive consumption, inclusive of excessive consumption, binge drinking, hazardous drinking, or disordered consumption, of which, the majority of studies reported insignificant results. The three relevant included studies with significant results found that excessive consumption was significantly more common among frail participants (P < .001),^204^ was at baseline associated with a higher increase in frailty index value per five years,^197^ and was predictive of shorter TLs (P = .045).^78^ One study analyzing post-mortem brain tissue reported mixed findings, wherein alcohol use disorder (AUD) was strongly correlated with biological age acceleration in DNAmAGE (r=0.93), ARR (r=0.99), PhenoAge (r=0.79), and SkinBlood (r=0.82) but was weakly correlated with other clocks and TL.^250^ Similarly, Faul and colleagues^120^ found that heavy drinking was significantly associated with increased biological age measured by PhenoAge (p < 0.01), GrimAge (p < 0.001), and adjusted (principle components) GrimAge (p < 0.001) clocks, while all other measures were insignificant. Eight studies found insignificant results, with excessive consumption unrelated to TL,^36,39,96^ accelerated biological aging,^41,162,172,220^ serum Pi levels.^165^

Two studies assessed alcohol consumption via methylation status at cg23193759; one noted that this was unrelated to Horvath and Hannum biological age.^246^ The other study compared cg23193759 to each loci of the Hannum clock; with methylation status of 55 of the 71 loci significantly related to cg23193759.^245^ Another two articles studied biomarker γ-glutamyl transferase as a biomarker of alcohol consumption with differing results; while authors of one study reported that alcohol abuse was related to significantly shorter telomeres (P < .001),^48^ Martens and colleagues^85^ found no difference in telomere quartile by drinkers vs non-drinkers. Finally, one study used methyl DetectR values for alcohol consumption per week and found that alcohol use was insignificantly associated with DunedinPace epigenetic age acceleration.^167^

### Other Substance Use

A total of thirteen studies examined the consumption of either specific drugs such as methamphetamine, nicotine, or marijuana, or general drug use or disorders in relation to aging. Of these, five reported significant findings,^138,164,202,227,251^ four reported insignificant findings,^102,184,210,246^ and three reported a mixture.^176,220,250^ Five of the included studies examined overall drug use.^102,164,176,202,210^ Two of these studies with significant findings examined the presence of a substance use disorder (SUD); the researchers found that reporting a substance dependence disorder was significantly correlated with TL at age 38(P = .021),^164^ and reporting a SUD was significantly associated with accelerated GrimAge (P = .01).^202^ Recreational drug use (excluding marijuana) was not significantly related to the rate of aging among HIV-positive individuals and controls.^102^ In a study examining Caucasian adults with a depressive disorder and controls, neither drug use nor drug dependency had a significant effect on TL. Watkins and colleagues^176^ reported that lifetime nicotine dependence did predict significantly shorter TL (P = .02), and was significantly more common among those participants with shorter telomeres compared to those with normal length telomeres (P = .005). Cabrera-Mendoza and colleagues^250^ found higher blood DNAmPhenoAge in individuals with SUD, including AUD, stimulant use disorder, and opioid use disorder compared to control groups (p = 0.0104). No significant differences were found in TL between individuals with SUD and controls. Hawn and researchers^220^ found that lifetime AUD did not predict GrimAge biological age acceleration (P = 0.107) but that SUD did (P < 0.001). No significant differences were found in lifetime alcohol or drug use disorders.

Watkins and colleagues^176^ examined a combined measure of drug and alcohol disorders, but no significant differences in TL was found. They also reported that lifetime nicotine dependence significantly predicted shorter TL (P = .02), and were significantly more common among those participants with shorter telomeres compared to those with normal length telomeres (P = .005). In contrast, an inverse relationship between nicotine consumption and TL was insignificant in another study.^184^ Lei and colleagues^246^ were the only study to solely examine marijuana use, as they used this primarily as a covariate in their model predicting methylomic age. They found that marijuana use was not a significant predictor of Hannum’s or Horvath’s age. Peixoto and colleagues^138^ examined cannabis use in people living with HIV and found that cannabis was more likely to be consumed in the group with higher biological age than chronological age as opposed to the group with a biological age less than or equal to their chronological age, indicating significant usage differences between the groups (p = 0.045). Mehta and colleagues^251^ assessed the role of methamphetamine, and divided their sample based on HIV status and methamphetamine dependence; in their model adjusting for covariates, being HIV+ (P = .0027) and dependent on methamphetamine (P = .0058) were associated with significantly shorter telomeres length. This equated to an increased biological age of 22.3 years in the HIV+ and methamphetamine dependent group. Minami and researchers^227^ reported that past and present substance use (cannabis, amphetamines, legal intoxicants, and 5-MeO-DIPT) was associated with significantly shorter TL (P = .0058). In their multivariate regression modeling examining only individuals with HIV, substance use continued to predict shorter telomeres (P = .0001).

### Emotions and Personality

The role of emotions and personality in aging was examined in nine studies.^54,83,84,129,167,176,187,188,202^ This includes three studies which examined overall emotions and affect, of which two used the Positive and Negative Affect Scale (PANAS),^84,129^ and one study assessed 10 negative and 10 positive emotions (e.g., gratitude, joy).^83^ Cruz-Almeida and colleagues^129^ reported conflicting results; while greater emotional stability was significantly associated with a younger epigenetic age (P = .027), epigenetic age was not correlated with positive or negative affect. In a study focusing on African Americans, telomere was not correlated with positive or negative affectivity.^84^ Similarly in a study of middle-aged adults, TL was not significantly correlated with either positive or negative emotions.^83^

Geronimus and colleagues^54^ examined anger and hopelessness. Hopelessness was associated significantly with a reduction in TL (P < .05), whereas anger was unrelated to TL. In a study comprised of military veterans, Watkins and fellow researchers^176^ found that individuals with shorter telomeres reported significantly increased hostility and difficulties controlling anger (both P = .001) compared to individuals with normal TL, but no difference in aggressive urges and impulses. Similarly, researchers found that study participants with a history of antisocial behaviour presented with accelerated biological aging at midlife, particularly those with life course-persistent antisocial behaviour (P ≤ 0.001).^188^ Tamman^202^ assessed a multitude of personality variables (i.e., hostility, openness, extraversion, emotional stability, conscientiousness, agreeableness, gratitude, optimism, and curiousity); veterans with higher scores of openness, gratitude and curiosity were at significantly lower odds of having accelerated GrimAge, whereas hostility, extraversion, emotional stability, conscientiousness, agreeableness, optimism were not related to GrimAge.

### Cognition and neurology indicators

#### Cognitive Functioning

Cognitive functioning and aging were assayed in 47 studies (Zhang 2014a).^46,60,71,77,78,81,95,99,104,120,126,127,129,138,140,148,153,156,172,181,205–207,211,220,222,251–271^ Three of these studies assessed intelligence independently to aging,^77,81,253^ two used intelligence testing as a means of determining cognitive function,^153,258^ and two studies considered independent measures to report on both cognitive functioning and intelligence.^254,255^ It should be noted that memory was grouped with cognitive functioning in nine of the articles mentioned.^60,104,140,181,211,254,260–262^ Dataset overlap was present in studies which used data from the Dunedin Study^153,252,255^ or the HRS.^104,120,126,127,156^ TL was the most utilized biological indicator, assessed in 16 of the studies mentioned above.^46,71,77,78,81,95,99,205,251,252,258,259,264,265,269,270^ Lower intelligence was associated with shorter TL at T3 (P = .001),^81^ and worse performance on the digit span task predicted shorter TL (P = .005).^265^ Cognitive impaired adults who had been exposed to abuse had shorter TLs compared to those who had not been abused (P = .002).^78^ Ryan and colleagues^95^ found that within the whole sample, there was a positive correlation between MMSE scores and TL (P = .01), however after age adjustment this finding was not significant (P = .14). Processing speed and motor performance was positively correlated with TL (range of P = .013 - .097), however global performance, executive functions, memory, learning, and verbal functioning were not significant.^251^ Insignificant findings between TL and cognitive functioning were reported in 11 of the articles.46,71,77,99,205,252,258,259,264,269,270

Ten of the studies used Horvath and Hannum’s clocks to predict DNAm age.^60,120,129,140,172,181,260,263,268,269^ Increased DNAm age was associated with lower cognitive function,^60,181^ and decreased selective attention abilities (P = .041).^263^ Epigenetic age was significantly correlated to lower memory scores (P <.01), however not Dimensional Change Card Sort, Pattern Comparison, or Flanker Inhibitory control and attention.^129^ Beydoun and colleagues^140^ reported Hannum’s algorithm predicted worse cognition with increased aging only among men in visual memory, visuoconstructive ability, attention, and processing speed (all P < .01), but not while using Horvath’s algorithm. Similarly, another report found cognitive dysfunction could be predicted using Hannum’s method, but not Horvath.^120^ Four studies did not find significant findings between DNAm age and cognition after age adjustment.^172,260,268,269^

The Klemera-Doubal method measured aging in six studies.^252,255,257,258,262,266^ Four articles showed that participants with increased biological ages had lower cognitive functioning (three P < .001,^252,255,258^ one P = NR).^257^ Similarly, Belsky and colleagues^255^ showed worse cognition associated with lower IQ scores (P = .01). Frailty and mortality were increased in individuals with lower MMSE scores, indicating cognition as a salient biomarker of aging (P < .001).^266^ Alternatively, cognition was weakly associated to biological age estimates.^262^ Levine’s method was used to predict biological age in two of the studies; ^253,258^ one reported a significant negative relationship between biological age and cognitive function (P < .001),^258^ and the other found that full scale intelligence quotients were not correlated with biological age.^253^

Multiple biological clocks were used in 12 studies that assessed cognitive function.^60,104,120,127,138,172,220,222,252,267,269,271^ Significant findings of increased PhenoAge and GrimAge were found with worse cognition (P = NR).^60^ GrimAge was also significantly associated with cognitive disinhibition.^220^ Similar results were found with increased MOCA and MMSE errors positively correlating with PhenoAge and GrimAge acceleration (all P<.01), however these effects did not survive adjusted models.^172^ GrimAge was significant in explaining differences between men and women’s cognitive function while Horvath, Hannum, and PhenoAge were insignificant.^127^ PhenoAge and DunedinPACE was found to predict cognitive dysfunction, though GrimAge was insignificant.^120^ Scores of MMSE and MoCA assessments worsened with older PhenoAge and DunedinPACE (P=NR).^267^ Similarly, MMSE scores were found to be a viable indicator of biological aging, when researchers determined 7 variables to compute age.^271^ Cognitive scores were not associated with GrimAge in another report.^104^ Cognitive function was significantly associated with the 71-CpG clock (P<.0001) but not the 353-CpG clock or 99-CpG clock.^252^ When compared to controls, subjects with autism showed a positive correlation with one component of GrimAge (DNAmPAI-1) while Horvath’s clock, Hannum’s clock, and PhenoAge were insignificant.^222^ Biological age calculated using Aging AI3.0 was not significant to MMSE scores (Peixoto et al., 2023).^138^ PhenoAge, GrimAge, and the 7-CpG clock were insignificant to MMSE scores.^269^

Homeostatic dysregulation was utilized in three studies to analyze aging; increased homeostatic dysregulation was correlated with decreased cognition in each of these articles (two P < .001,^252,258^ one P = NR).^257^

Two studies considered CpG methylation as a biological indicator.^206,207^ Impaired cognition was found to be significantly associated with increased methylation at CpG_5 unit (P=.039), however not associated with the other three CpG units measured.^207^ Similarly, cognitive function was insignificant to methylation levels of CpG units RAB32 and RHOT2.^206^

Thirteen articles each analyzed differing biological indicators._126,148,153,156,205,211,213,252,254,256,261,270,272_ GDF-15 levels positively correlated with cognitive impairment (P=0.01).^148^ Similarly, GDF-15 showed an inverse association with executive function, but was insignificant to other cognitive domains.^211^ Cognition positively correlated with fBioAge (P < .0005).^261^ Faster DunedinPoAm in individuals predicted worse cognitive abilities and IQ scores (P < .002).^254^ Worse fluid cognition was associated with a higher brain-predicted age difference score, indicating increased aging (P = .007).^256^ Carriers of Apolipoprotein-ε4 (APOE e4) were shown to have worse learning performance than non-carriers (P < .001).^126^ At 45 years of age, higher suPAR levels were found to be associated with lower intelligence scores (P < .0001).^153^ A decline in cognitive function was significantly associated with increased pace of aging (P < .001).^252^ Brain-age estimates utilizing resting-state functional connectivity and structural connectivity measures were significantly younger than participants cognitive age (P<.01).^270^ Monocyte Chemotactic Protein 1 (MCP-1) and Regulated on Activation, Normal T cell Expressed and Secreted (RANTES) were both found to be uncorrelated with cognition.^205^ Crimmins^156^ utilized 10 biomarkers including: albumin, alkaline phosphatase (ALP), CMV, PUFF, CRP, creatinine, total cholesterol, BUN, fasting glucose, and systolic blood pressure; results showed that cognitive dysfunction increased with increasing biological age (P = NR). Telomere activity was positively correlated with attention, motor speed, and executive functions (all P < .05), but related to visuospatial, learning abilities, or subjective cognitive decline.^272^ Additionally, Carroll and colleagues^272^ found that DNA damage was increased with worse executive functioning (P = .027), but not significant with the other cognitive domains tested. Higher scores on the frailty index was found to correlate significantly with worse attention and processing speed, though executive functions were unrelated.^213^ Finally, soluble tumor necrosis factor receptor was unrelated to cognitive function.^272^

#### Cognitive Impairment

Ten studies looked at cognitive impairment in relation to aging using various analysis.^56,59,78,181,213,240,267,268,273,274^ Populations varied; participants were from VITAL-DEP,^181^ the Washington Heights–Inwood Community Aging Project,^56^ Chinese abused and non-abused seniors (aged 55+),^78^ along with comparison between healthy individuals and those with mild-cognitive impairments or Alzheimer’s disease (AD).^59,273^ Definitions of cognitive impairment and scales varied.

Three studies assessed cognitive impairment in relation to TL,^56,59,78^ of which two found cognitive impairment to increase the rate of biological aging (both P < .01).^56,78^ Lee and associates^59^ found mixed results, with no significant correlation between either AD or MCI with TL in healthy individuals. However, when comparing TL between AD and MCI groups, there was significantly shorter telomeres found in those with AD (P = .02). Of the 76 saliva components analyzed, 36 had increased levels in participants with AD when compared to healthy controls (P = NR).^213^ There was a significantly higher risk of mild cognitive impairment in individuals with coronary heart disease for every 5-year increase in Horvath’s biological age; though Hannum, PhenoAge, and GrimAge were insignificant to mild cognitive impairment and dementia status.^274^ When analyzing the sample as a whole, Horvath’s biological age was not predictive of mild cognitive impairment or dementia.^274^ Increased aging utilizing DunedinPACE and Horvath measures showed a higher risk of dementia (P=NR).^267^ One study compared cognitive impairment to DNAm age using Horvath’s clock method;^181^ researchers found no significant correlation between DNAm age and cognitive impairment. Waziry and colleagues^240^ found that neither Dementia or AD were not predictive of aging, as assessed using the Klemera-Doubal method. Horvath’s age acceleration was insignificant to Parkinson’s severity.^268^

#### Memory

Twelve studies looked at memory in terms of aging in different populations,^95,126,127,181,207,213,220,253,267,272,275,276^ such as women with early-stage breast cancer,^272^ severely depressed and non-depressed individuals, and different age groups.^207^ GrimAge had a significant positive association with verbal memory recall.^220^ Carriers of APOE e4 had significantly worse memory scores than non-carriers.^126^ Slower GrimAge acceleration in women explained in part higher verbal memory scores in women, while PhenoAge, Hannum’s clock and Horvath’s clock were insignificant.^127^ Five studies reported insignificant results,^95,181,253,272,276^ and one had mixed findings.^207^ Two of the studies that were insignificant used TL as a biological indicator of aging,^95,272^ while Vyas and colleagues^181^ considered Horvath and Hannum’s biological clocks. Although Beam and fellow researchers^253^ found no statistical significance between episodic memory and aging biomarkers, they found a strong correlation between lower episodic memory and increased biological aging. Another study resulted in a significant correlation between immediate recall and CpG_5 methylation (P = .036), however no significance was found between the methylation levels with delayed recall testing in their study (P=.071).^207^ PhenoAge was inversely related to memory performance in only Caucasian women, whereas older GrimAge and DundedinPACE of aging associated to lower memory scores in Caucasian men and women.^275^ Both PhenoAge and DunedinPACE of aging had a negative association to episodic memory and the Logical Memory Test (P=NR).^267^ Episodic memory was not significant when using frailty index as a biological aging measure.^213^ Working memory had a positive correlation to DNAmTL in controls when compared to master rowers, though it was not significant.^276^

### Neurological indicators

There were 13 studies that used neurological measurements to assess aging.38,130,132,143,209,213,220,224,227,263,264,277,278 The most common biomarker selected was TL. Two of the studies that measured TL found significant findings overall.^224,227^ Minami and colleagues^227^ reported that shorter TL was associated with white matter hyperintensity (P = .0006). Barha and fellow researchers^224^ assessed hypothalamic-pituitary-adrenal axis activity, finding a negative relationship to TL (P = .007). Three studies that compared TL to neurological measures found mixed findings overall.^38,209,264^ Results differed between patients and healthy controls in a study conducted by Hassett and colleagues,^209^ which reported a significant positive correlation between TL and gray matter volume of the right primary somatosensory cortex, left middle frontal gyrus, and the left precuneus in patients with fibromyalgia (P < .05), but no significant correlation for healthy participants. Wikgren and colleagues^264^ found no significant correlation between TL and white matter intensity across the entire population study, however there was significant shorter TL above 69.9 years of age (P = .009). Révesz and colleagues’^38^ results suggest a non-linear association between only one of the four HPA-axis measures used (AUCi) to compare TL (P = .01), after sociodemographic adjustment.

Of the four studies that used Horvath’s biological clock to predict aging, two of these studies resulted in significant findings,^132,143^ and two had mixed results.^130,263^ Global cortical thickness was negatively associated with biological and chronological age (P < .001),^132^ and neurosensory impairment was found to be significantly associated with accelerated aging (P=.02).^143^ Hodgson and colleagues^130^ detected significant negative relationships between both chronological and epigenetic age when compared to global white matter track intensity (P < .05) in all regions except for the hippocampus. Greater DNAm age was associated with increased anterior cingulate cluster gamma activity (P=.02), but not in the left superior parietal and left primary visual cortices.^263^

In terms of brain matter, total brain matter, white matter, grey matter, along with fibular nerve conduction speed all declined with age (all P < .001) when looking at various biological indicators (i.e., body composition, energetics, and homeostatic mechanisms).^277^ Individuals with older GrimAge had significant astrocyte damage and reduced cortical thickness in the right lateral orbitofrontal cortex and left fusiform gyrus (P < .05).^220^ Angioni and colleagues^278^ found that persistent fatigue significantly associated with increased white matter hyperintensity volumes but not cortical thickness or sub-cortical volumes. Frailty index scores were not associated with white matter hyperintensity volume.^213^

### Other Cognitive Outcomes

Other domains of cognition were analyzed in terms of aging in a single study, and included mind wandering, psychological inflexibility and experiential avoidance, mental fixations, awareness, and rumination.^208^ Increased mind wandering was significantly associated with shorter TLs (P < .01) as assessed through self-reporting. Psychological inflexibility and experiential avoidance, mental fixations, awareness, and rumination were not significant in comparison to TL.

### Social Relationships and Interpersonal Systems Indicators

Sixty-four of the included studies reported results on variables related to social relationships and dynamics. Dataset overlap was common among these studies; ten studies utilized data from the FACHS,^41,42,119,125,162,167,171,174,279,280^ ten used data from HRS,36,37,105,120,136,156,186,187,281,282 five used data from the Dunedin cohort,163,164,188,189,283 and another two studies used the same data.^98,184^

### Romantic Relationships and Intimate Partner Violence

A total of 21 studies reported the impact of marital or relationship status on aging; these studies compared a variety of categories, such as being currently married, divorced, widowed, or never married. Results were mixed; five studies reported that marital status significantly affected aging,3,103,106,180,202 12 found insignificant results,^35,37,119,138,162,167,174,177,182,186,280,284^ two studies found that this effect depended on the other variables that were controlled for,^42,99^ and two studies reported mixed results.^122,171^ Of the studies reporting significant effects, Ng and colleagues^180^ found that being single, divorced, or widowed was associated with having a significantly older biological age, Simons and fellow researchers^279^ reported that being married predicted a decreased GrimAge (P < .01), and Tamman^202^ reported that those who were married or cohabitating with a partner were less likely to have an accelerated GrimAge. Han and colleagues^103^ found significant differences in both biological aging and the difference between biological and age between the three examined categories of marital status (P < .001; P = .0159; respectively), with individuals reporting marital problems having the highest biological age and difference score, whereas single people had the lowest values of these variables. In contrast, analyses conducted by Simons and colleagues^42^ first indicated that marriage did not significantly predict GrimAge, but once gender was added to the model (i.e., being male), marriage was then associated with decelerated aging. The effect of marital status also differed in one other study,^99^ as being married was significantly predictive of longer TL (P = .20) in the first examined linear regression model, but this effect was insignificant once other variables were controlled for. Of the remaining five studies with insignificant findings, marital status was not significantly related to TL in three studies,^35,37,186^ as well as CpG methylation and DNA methylation age acceleration in one study each (Lei et al., 2019; Li et al., 2019, respectively).^162,182^ The two studies reporting a mixed effect had opposite results: Galkin and colleagues^122^ reported that being married reduced the rate of aging whereas being widowed did not have an effect, and Simons and colleagues^171^ reported that widowhood was significantly related to accelerated GrimAge, but getting married did not impact any of the clocks examined.

Bourassa and colleagues^163^ compared relationship variables to pace of aging; being in a relationship at more time points was protective, as pace of aging decreased with the number of romantic relationships. Individuals who did not report a romantic relationship at any of the study timepoints reported a significantly increased rate of aging (P < .001), which was equivalent to a 2.9 year increase over the 19 study years. Of those who were in a relationship, higher relationship quality was associated with a slower rate of aging (P < .001). However, relationship length was not related to pace of aging. Rentscher and fellow researchers^49^ compared CDKN2A expression with both relationship closeness and satisfaction. Relationship closeness was not correlated with CDKN2A expression; however, this did significantly interact with perceived stress to affect CDKN2A expression and together accounted for 17.8% of the variance in CDKN2A expression. Specifically, CDKN2A expression was elevated among individuals with either low or moderate relationship closeness and greater perceived stress. This stress-buffering effect of relationship closeness on CDKN2A expression was independent of relationship satisfaction. In addition, relationship closeness was significantly negatively correlated with chronic stress exposure and accumulated daily stress, but not perceived stress. Relationship closeness was also significantly greater among men than women (P = .03). Forrester and colleagues^212^ examined both social ties and social strains; mean biological age decreased over time inversely with the number of social ties and social support. Similarly, another study examined social support and contact with others, with the results indicating that additional contact with children and friends at baseline was associated with a lower GrimAge (both P < .001), contact with friends was associated with a slower pace of aging (P = .009), and an increase in family contact over time was associated with a lower Hannum age (P = .015). Tamman^202^ examined social network size, perceived social support, and attachment style, but none of these variables were significantly related to GrimAge.

One study examined intimate partner violence.^163^ Pace of aging and both partner violence victimization and perpetration were positively associated (both P < .001), however perpetration was unrelated to pace of aging when examined in the same model as victimization. In terms of victimization, both psychological and physical victimization were related to an increased rate of aging (both P < .001).

### Childhood and Family Relationships

A total of 36 studies examined variables related to family relationships and childhood. Fourteen studies assessed childhood socioeconomic status in relation to aging,^41,45,63,101,106,120,123,136,156,163,188,189,283,285^ of which four utilized data from the Dunedin cohort,^163,163,188,283^ and three from the HRS.^120,136,156^ Six studies reported that decreased childhood socioeconomic status was predictive of an increased pace of aging.^101,136,163,188,189,283^ Another six studies reported mixed effects, with most reporting that the significance of this relationship depended on which clock was examined.^106,120,123,285^ Of the other two studies, one reported that only the father’s education (P = .03) was a significant predictor of TL, with these 2 variables inversely related,^45^ and another study examining paternal education found that decreased father’s education and experiences of childhood food deprivation were significantly related to shorter TL for women (both P = .04), but not men.^63^ Two studies reported insignificant results related to childhood socioeconomic status, with this unrelated to accelerated aging in one study^41^ and mortality in another.^63^

Three studies examined childhood health with different results; increased pace of aging was associated with reporting worse childhood health (P < .001),^163^ whereas a composite measure of childhood hardship, childhood health, and adult trauma was responsible for 1.3% of variance in mortality, 1.2% of variance in cognitive dysfunction, 4.7% of variance in the number of reported diseases, and 9.9% of variance in difficulties in activities of daily living (ADL) in one study.^156^ A single study examined childhood health, family longevity, and other childhood-related variables; pace of aging was significantly negatively associated with the age of longest-lived grandparent (P = .004), childhood social class (P < .001), childhood health (P = .004), childhood IQ (P < .001), and childhood self-control (P < .001).^283^

Trauma and abuse experienced during childhood were examined in nine studies. The majority of these studies assessed a composite variable of abuse and neglect, which included some combination of physical abuse, emotional abuse, sexual abuse, and/or neglect. Of the six studies examining a composite abuse and neglect, studies found that this was significantly associated with the number of chronic health conditions later in life (P < 0.001),^194^ positively associated with accelerated PhenoAge (P = 0.013),^286^ and were split regarding both TL with two studies indicating an insignificant relationship^175,210^ and one study finding significantly shorter telomeres with more types of abuse experienced,^87^ and these studies were also split regarding GrimAge, with one reporting no relationship^286^ and another reporting that early life abuse and early life trauma were significantly correlated with GrimAge acceleration (both P < .01).^159^ Neglect was assessed in three studies; results of one study indicated that this was not associated with either PhenoAge or GrimAge,^286^ which was confirmed in another in which neglect was not related to biological age according to the Horvath, Hannum, PhenoAge, and GrimAge clocks.^159^ The third study indicated that the relationship between aging and neglect depended on the type of neglect, as childhood physical neglect was related to shorter TL (P = .007) and explained 4.2% of the variance in TL, whereas emotional neglect was insignificant.^210^ Sexual abuse and physical abuse were compared to indicators of biological aging in two and three studies respectively, physical and sexual abuse were separately unrelated to TL in two of these studies,^210,223^ and emotional abuse (95% CI: 0.00, 0.12) was associated with higher DNAm GrimAge acceleration in the third study.^124^ Emotional abuse was not associated with TL in one study.^210^ Another study utilized different conceptualizations of abuse; Surtees and colleagues^178^ reported that experiencing childhood difficulties and trauma was related to having shorter telomeres (P = .015).

A total of 13 studies examined adverse childhood experiences (ACE) in relation to biological aging. Of the six studies with significant results, more ACEs were significantly related to an increased midlife aging factor score,^189^ pace of aging,^163,189,283^ BrainAge,^189^ facial age,^189^ TL,^177^ GrimAge,^174^ DunedinPoAm,^174^ and mRNA-based biological clock.^41^ In comparison, results were insignificant in five studies, with ACEs were unrelated to GrimAge,^119,280,286^ TL,^218^ the Hannum 71-CpG clock,^3^ PhenoAge,^286^ or mtDNAcn.^218^ Another two studies reported mixed results regarding ACEs and biological aging, including one study finding significant and insignificant relationships depending on the biological clock examined,^124^ and another study reporting that childhood exposures were significantly related to shorter telomeres among men (P = .045), but not women.^36^ Another four studies examined specific experiences during childhood; those reporting a childhood police encounter had significantly elevated biological ages in two of three examined clocks in one study,^282^ another study grouped participants based on their adolescent lifestyles, with differences detected in some clocks but not others^203^ and researchers of one study reported that having an internalizing disorder persistent from age 11 was associated with having significantly shorter TL at age 38 among men (P = .005) but not women, and did not find an association between childhood maltreatment and TL at age 38 was found.^164^ The fourth study conducted by Langevin^188^ reported that pace of aging was significantly correlated positively with self-control difficulties during early childhood and childhood maltreatment (both P ≤ 0.001).

Three studies examined variables related to family relationships and history. Lincoln and colleagues^37^ examined both positive social support and negative interactions with family members. In their multivariable linear regression model, higher levels of social support was predictive of shorter TLs (P < .05). Negative interactions with family had a significant interaction effect with social support (P = .046), but not quite race (P = .084). One study examined the potential mediating role of weekly versus no contact with children in a model predicting TL; contact with children was not a significant mediator.^66^ Savolain and colleagues^67^ examined the effect of family separation during World War II on TL but did not find a significant effect. However, those who did experience family separation more often reported having coronary heart disease (P = .012), a previous stroke (P = .013), less education (P = .027), having a father who worked manual labour, a younger mother at age of delivery (P = .003), and more experiences of emotional or physical trauma (P = .044).

### Neighbourhood Disadvantage and Experiences

Eleven studies compared aging with neighbourhood-related variables. Results indicated that higher neighborhood disadvantage was significantly related to accelerated aging,^42,125,285^ and that community disadvantage, which was characterised as community disorder (i.e., criminal acts) was significantly related to accelerate aging for juvenile participants (P = .026), but not adult participants. Lei and fellow researchers^162^ also assessed neighborhood disadvantage, which they conceptualized as including median household income, the percent of unemployed males, the percent below poverty threshold, the percent of single mother families, and the percent of individuals receiving public assistance; neighborhood disadvantage was significantly associated with accelerated biological aging (P ≤ .01). In contrast, Berg^119^ conceptualized community disorder as the frequency of crime in the neighbourhood, and reported that this was not predictive of GrimAge. A later study by Berg^280^ identified that this variable was not predictive of GrimAge, but they were positively correlated (P = .011). Martin and colleagues^287^ examined three facets of neighborhood environment: neighborhood observations, neighborhood poverty, and social cohesion score. Of the 19 examined neighborhood observations, only the indicator regarding abandoned cards and homeless people was significantly associated with Horvath age acceleration, Hannum age acceleration, and PhenoAge acceleration (P = NR). In comparison, neighborhood poverty and social cohesion were not associated with any measure of biological aging, although results indicated a minimal sex effect. Among Black Americans, Geronimus and colleagues^54^ identified that being highly satisfied with one’s neighborhood as well as perceiving their neighborhood to have more positive and less negative physical characteristics was associated with longer TLs (P < .03, P < .001; respectively). Counterintuitively, stress related to feeling unsafe in one’s neighborhood was associated with longer telomer length (P < .001). In comparison, Yen and fellow researchers^99^ assessed perceived neighbourhood experiences, as well subscales to examine neighbourhood-related social support, perceived security, and available services and facilities; these variables did not predict TL in any of the examined models. Neighbourhood resources was unrelated to biological age acceleration in one study.^212^ Recent relocation within the past year did not impact biological aging according to one study.^125^

### Work Environment and Experiences

Various work-related variables were reported in 4 studies. Chmelar and fellow researchers^184^ examined psychosocial work conditions and occupation burnout among geriatric care home employees. After adjustment for variates, reporting burnout involving depersonalization was significantly related to having shorter telomeres (P = .02). However, TL was not related to job autonomy, work-related time pressure, occupational social support, work ability, and emotional exhaustion. Among professional caregivers, neither TL or chronological age were correlated work learning opportunities or selection, optimization and compensation strategies employed during work.^98^ Ahola and colleagues^53^ sorted individuals by work-related exhaustion into three categories of exhaustion: no, mild, or severe. After adjusting for age and sex, work-related exhaustion was significantly and negatively associated with TL (P = .031).

Differences in exhaustion was also related to several demographic variables; individuals with high exhaustion were more often female, unmarried, working manual labour, and ill with a somatic or mental illness. One study included young and middle-aged South African teachers; they reported that work stress was not significantly related to TL, but was significantly higher among Black South Africans compared to White South Africans.^48^

### Experiences of Discrimination and Unfair Treatment

Eight studies examined discrimination, of which 5 examined a Black population, one study included low to middle-income mothers with a primary identity of Hispanic/Latina, Black, or non-Hispanic White, and one study included a general population. The three studies which assessed experiences of reported racial discrimination reported conflicting findings; one reported that racial discrimination was significantly correlated with increased accelerated GrimAge (P ≤ .05),^42^ one found that racial discrimination was not correlated with TL,^169^ whereas one study reported decelerated biological aging with increased racial discrimination.^212^ Rej and colleagues^93^ examined unfair treatment to both the individual and to someone close to the participant, including both general unfair treatment and treatment they attributed to race. Race-related unfair treatment experienced by the self was significantly correlated with TL (P = .046), and significantly predicted TL in their multiple linear regression models (P = .03). In contrast, race-related unfair treatment experienced by others, and general unfair treatment for both themselves and someone close to them were not related to TL. In a study conducted by Lee and fellow researchers,^186^ increased experiences of general discrimination were related to shorter TL (P = .018) after adjusting for chronological age. This relationship remained after controlling for further covariates. Further analyses divided their sample into categories of discrimination severity: low, moderate, and high. Their results suggest a threshold effect, as individuals with moderate discrimination experiences did not have shorter TL, but individuals with high discrimination experiences did have significantly shorter telomeres (P = .009). In the study with a general population, perceived unfair treatment did not influence TL.^54^ Everyday discrimination was not correlated with TL^157^ or biological age,^105^ but lifetime discrimination was related to TL in one study ((p < .001).^157^

### Other Social Outcomes

Three studies assessed loneliness,^122,167,187^ two of which used items from the same scale.^167,187^ Both reported significant findings; Beach and colleagues^167^ found that loneliness was significantly associated with DunedinPace linear growth, particularly within-person experiences of loneliness (p < 0.05), and Crowe and researchers^187^ reported that participants who experienced persistent feelings of loneliness compared to participants who experienced intermittent loneliness had a higher phenotypic age, were at an increased risk of mortality, and a higher risk of disease. In contrast, one study identified that individuals who reported rarely feeling lonely did not have a significantly different pace of aging.^122^

One study conducted by Vyas and researchers^181^ examined frequency and satisfaction of received social support in relation to DNAm age and accelerated aging; they found that as social support increased, both DNAmage and accelerated aging significantly decreased (P = .048; P = .01, respectively). Ng and colleagues^180^ assessed the potential predictive role of engagement in social activities (e.g., attending religious worship, sport events, card games) in regard to biological and chronological age (P < .001); researchers found that social activities were a protective factor regarding aging. While one study found that negative social interactions with family and friends was not associated with shorter TL (P = .06),^54^ these interactions may still have meaningful implications. Increased cumulative social disadvantage, characterized by Simons and colleagues^42^ as including education, income, living in a disadvantaged neighbourhood, and discrimination, was found to be significantly related to accelerated aging via GrimAge (P ≤ .01) in two studies.^42,280^

Social desirability bias was examined in one study,^169^ which found that the degree of social desirability bias was significantly inversely related to TL (P < .05). Lucas and colleagues^84^ examined trait justice views; they found that TL was not significantly correlated with any examined facet of trait justice beliefs. However, when participants were divided by their belief in justice for self, a group difference was found, as age and TL were significantly negatively correlated for those with a low belief in justice for self (P < .001), but not for those with a high belief. This relationship was moderate by beliefs about justice for others (P = .038).

### Lifestyle indicators

A total of 115 studies examined lifestyle variables, such as physical activity or nutrition. Across the review, dataset overlap in included studies occurred in 73 studies. Of the studies examining lifestyle indicators, dataset overlap occurred with data from: NHANES in ten studies,^5,10,32,33,144,288–291^ FACHS in eight studies,^3,41,42,162^ Moli-Sani in four, UK biobank in three, Korean Genome Epidemiology Study population in three, CALERIE in two, National Health and Resilience in Veterans Study population in two, European Prospective Investigation into Cancer-Heidelberg cohort in two, Netherlands Study of Depression and Anxiety population yes,^38–40^ Health in Men Study population in two,^238,292^ Berlin Aging Study in two, HRS population in two,^36,186^ Korean KNHANES study in two,^103,161^ Multiethnic Study of Atherosclerosis (MESA) study population in two,^45,46^ PREVEND cohort in two,^81,192^ PSOBID Cohort in two,^96,165^ VITamin D and OmegA-3 TriaL-Depression Endpoint Prevention study in two,^146,181^ Dunedin in two, China Kadoorie Biobank in two, Lothian Birth Cohort in two, Sister Study in two, and another two studies with the same data comprised of caregivers.^98,184^

### Physical Activity and Sedentary Behaviour

Sixty-nine studies assessed physical activity variables in comparison to biological aging and biomarkers. Physical activity was examined as either interval categories of intensity or duration, or a dichotomous variable or being active or inactive. This was most commonly compared to TL; in 28 studies, eight reported that decreased physical activity was significantly related to shorter TL, ^32,46,69,73,81,192,199,293^ whereas two reported exercise was inversely related to TL,^43,44^ albeit these used the same dataset, and another 17 articles found these were unrelated.^33,36,38,39,45,72,74,96,176,178,181,186,190,198,237,238,292^ One study had mixed results by gender, as while their dichotomous physical activity variable was not associated with TL in their overall sample, women reported significantly longer telomeres among those who did not exercise (P = .04), and men reported the opposite, in which men who exercised had longer telomeres (P = .03).^63^

Another 27 studies reported on biological aging; increased physical activity was associated with significantly younger biological ages or decreased age acceleration in 12 studies,^103,128,161,172,180,181,202,239,241,294–296^ with one study finding that those who engaged with less physical activity were six to eight years older biologically.^181^ Six studies had inconsistent results, including a mixture of accelerated aging and insignificant findings varying by clock or dataset,^116,121,179,235,297,298^ and 11 reported this relationship to be insignificant.^41,42,111,119,162,166,168,171,234,279,280,299^ Two studies implemented an intervention featuring exercise with mixed results; Ho and colleagues^300^ reported their the exercise intervention group reported improvements in KDM biological age and homeostatic dysregulation but not according to the Healthy Aging index, and improvements were larger for the exercise and diet intervention group. Loh and researchers^215^ reported no consistent relationship between exercise and changes in aging, but did find that participants who increased their steps more than the average participants had decreased DNAm age compared to those who were below average. None of the studies found accelerated aging with increased exercise.

Physically inactive people were more likely to be classified as a high mortality risk,^144,183^ have a worse frailty status,^197^ have higher ferritin levels (P < .001),^32^ increased carotid-femoral pulse wave velocity (cfPWV),^218^ and higher suPAR (P < .0001).^153^ One study reported that increased average physical activity was significantly related with lower: C-reactive protein, glycated hemoglobin, total cholesterol, serum urea nitrogen, serum alkaline phosphatase, systolic blood pressure, pulse, high density lipoprotein, white blood cell count, platelet count, and chronological age, and significantly higher serum albumin, diastolic blood pressure, hemoglobin, lymphocyte percent, hematocrit, and red blood cell count (all P < .001).^301^ However, physical activity was not related to stochastic epigenetic mutations (SEMs),^179^ mtDNAcn,^146^ or allele score.^69^

Two studies examined athletes. Aguiar and colleagues^302^ compared master athletes to untrained controls; master athletes had significantly longer telomeres than untrained controls (p < 0.05). Osthus and fellow researchers^303^ noted that chronological age interacted with athleticism, in that older athletes had significantly longer TL than older people who were moderately active (P = 0.04), whereas younger athletes did not differ from younger non-athletes.

Another four studies assessed sedentary behaviour. Results indicated that time spent sitting per week was significantly negatively correlated with TL (P = .002),^293^ significantly increased GrimAge,^295,296^ unrelated to aging acceleration in one study.^304^

### Nutrition

Various aspects of nutrition were examined in 41 studies. Five studies assessed nutritional quality or risk. Of these, nutritional status or risk was included in four studies, with poor nutritional status and a high nutritional risk associated with being significantly older (P < .001) in one study,^180^ whereas this was not predictive of biological aging outcomes in the other three, including mortality,^107^ TL, Normal T cell Expressed and Secreted (RANTES), or monocyte chemotactic protein 1 (MCP-1),^205^ and did not differ between aging accelerators versus aging decelerators.^138^ Additionally, results regarding diet quality differed; diet quality was unrelated to both Horvath and Hannum aging in one study,^111^ but improved diet quality significantly reduced GrimAge, DunedinPoAm, and PhenoAge in the second study.^171^ Vetter and colleagues^269^ reported that scores on the MNA was significantly associated with frailty score, physical functioning, and mobility (all P < .001). Two studies^42,305^ conceptualized diet quality by the consumption of fruits and vegetables, fatty foods, and fast food, but this was not related to accelerated aging in one study, and was related to one of four biological clocks in the other. Five studies reported calories consumed or calorie restriction. Overall calorie intake did not predict TL,^43,44^ but results differed for calorie restriction interventions. One study found that participants had significantly slower biological aging compared to the controls (p = .003) after a calorie restriction intervention,^306^ and another similarly reported that a calorie restriction intervention significantly improved KDM biological aging, homeostatic dysregulation and scores on the healthy aging index.^300^ A third study which implemented this type of intervention found reductions in DunedinPACE, but not in PhenoAge or GrimAge.^307^ Senior and colleagues^308^ examined macronutrient consumption and reported that increased consumption was associated with significantly lower biological age. One study compared the use of nutritional data to biological age, with those who used this information biologically younger (P < .0001).^161^

Different dietary patterns were compared related to biomarkers in six studies. Researchers from one study^44^ examined both a prudent dietary pattern, comprised of primarily fish, grains, legumes, vegetables and seaweed, versus a western diet, which was characterized as the consumption of primarily red or processed meat, refined grain, and sweetened carbonated beverages; results indicated that the prudent diet was significant related to longer telomeres (P = .027), whereas the western diet was not significantly associated with telomeres. Two studies examined the Mediterranean diet and reported that this diet pattern was associated with significantly decreased aging acceleration (P < .05).^168,309^ Esposito and researchers^310^ compared four diets (Mediterranean diet, Dash diet, Palaeolithic diet, Nordic diet); both the Mediterranean diet and Dash diet were associated with significantly decelerated aging, whereas the other two did not impact the rate of aging. Another study examined both the alternative mediterranean and Dash diet,^305^ and reported that both diets were associated with significantly reduced GrimAge, but were unrelated to Hannum, Horvath, and PhenoAge; this relationship was altered by physical activity. Another study implemented a dietary intervention, with the intervention group consuming a polyphenol-probiotic-enhanced diet (consisting of high dietary fiber and low energy dietary profile, and probiotic powder).^311^ Compared to the control group, the intervention group had significantly reduced CRP, IL-6, and IL-10 levels after two weeks.

Other studies focused on specific food items. Vegetables and fruits combined did not predict biological aging in six studies.^3,41,119,125,162,280^ In contrast, other studies indicated that the consumption of vegetables and fruits was significantly related to reduced frailty,^197^ longer TL (P < .0001), and both vegetables and fruit separately were also related to longer telomeres (all P < .001),^180,290^ but in one study this relationship disappeared for men when separated by gender.^290^ Another study also found that fruit consumption was predictive of significantly longer telomeres (P = .0081), but not vegetables.^44^

The majority of studies that examined protein focused on a specific protein source. The exception to this was a study by Yu and colleagues,^71^ in which overall protein intake was not related to TL for overall sample, or when divided by gender. Three studies assessed the impact of consuming fish; results were mixed, as this was significantly associated with a decreased rate of biological aging (p< 1.2× 10e−2),^231^ and was unrelated to TL,^44^ whereas one study found this differed by the type of fish, in that deep-sea fish was related to a younger BA (P = .002), but not other fish.^180^ Higher consumption of red meat was associated with significantly increased levels of Pi in men (P = .008),^165^ and when combined with processed meat in another study, was related to significantly shorter telomeres (P = .0073).^44^ Similarly, a study assessing solely processed meat found this was inversely related to TL (P = .01).^45^ Poultry and eggs were assessed separately in another study, but neither were related to TL.^44^

Dairy products were examined in three studies. Results indicated that increased consumption of dairy products predicted significantly longer telomeres (P = .0054),^44^ but cheese was not associated with serum PI.^165^ Researchers of one study examined milk based on consumption frequency and milk fat content; milk consumption did not impact TL, but telomeres did differ by fat content, with fat content and TL significantly positively related after controlling for covariates (P < .0066).^288^

Beverages were compared to markers of biological aging in four studies. Three examined soda; higher consumption of soda was related to significantly accelerated biological aging ( P < .001),^231^ serum Pi for women only (P = .008),^165^ and shorter telomeres (P = .04) in one study each,^44^ but another reported that TL and soda consumption was unrelated.^165^ Drinking coffee predicted significantly shorter telomeres (P = .05) in one study,^44^ did not impact aging acceleration in another,^180^ and a third study found that participants who drank 3-6 cups of coffee per day aged slower (P < .001), as they were biologically younger than their chronological age by 5.6 years.^231^ Results related to tea were inconsistent, with tea predicting younger biological age (P < .001)^180^ but not TL.^44^

Another 11 food products were reported in less frequently. Legumes were positively associated with longer telomeres in one study,^44^ and unrelated in another.^290^ Lee and colleagues^44^ reported that significantly longer telomeres were predicted by the consumption of nuts (P = .0080), seaweed (P = .0013), whereas telomeres were unrelated to rice, flour products, cereal, mushrooms, and kimchi. Other researchers have identified that potatoes were not related with TL (P = .030), whereas soy products were associated with having a younger biological age (P < .001).^290^ Tucker and researchers^289^ reported on the relationship between fibre and telomeres; after adjusting for covariates, these variables were significantly and positively related (P = 0.0101). Finally, biological aging was inversely related to polyphenol antioxidant content of an individuals diet (P < .0001), but biological aging did not differ by the total antioxidant capacity of their diet.^312^

Five studies reported the use of vitamin supplements. One study examined numerous vitamins,^43^ with results indicating that TL increased with higher consumptions of vitamin C (P = 0.04), folate (P = 0.05) and potassium (P = 0.04), but did not differ by consumptions levels of vitamin A, retinol, carotene, vitamin B1, vitamin B2, niacin, vitamin B6, vitamin E, calcium, phosphorus, iron, or zinc. Several studies implemented a nutritional supplement intervention, with one reporting significant reductions in biological age from baseline to post-intervention (p < .001).^217^ Chen and researchers^313^ assigned participants into four groups with differing levels of vitamin D3 supplements or placebo; both the groups with vitamin D3 supplementation of 4,000 IU/d and 2,000 IU/d had slower rates of biological aging (p = .046; p = .044, respectively).

Another study reported on the use of vitamin supplements in relation to ASPA, ITGA2B, and PDE4C over a one-year between groups; ASPA methylation was reduced in the group receiving vitamins D and calcium compared to the groups taking vitamin D, calcium and vitamin B group (p = 0.046), and groups did not vary by ITGA2B or PDE4C.^154^ After a Vitamin D intervention, participants of one study demonstrated significantly reduced Horvath and 7-CpG DNAmAA aging, but did not have changed rates of acceleration by the Hannum, PhenoAge, or GrimAge clocks.^314^ Tucker and colleagues^10^ found that dietary vitamin E and supplemental vitamins separately were inversely associated with gamma-tocopherol blood levels (p = 0.0059; p < 0.0001, respectively) and positively correlated with alpha-tocopherol blood levels (p = 0.0016; p < 0.0001, respectively), but not TL. Dietary copper consumption was assessed in one study; increased dietary copper was associated with longer telomeres.^315^ The overall number of dietary supplements and their frequency was also examined, but did not impact the rate of biological aging.^299^

Two studies assessed inflammation risk due to diet with mixed results; sex and BMI influenced the significant relationship between increasing values in inflammation measures and aging acceleration,^196^ and dietary inflammation was significantly positively correlated with biological age and phenotypic age, and negatively correlated with TL and klotho.^316^

### Engagement in Health-Related Activities

Meditation was assessed in two studies. One compared the rate of aging for long-term meditators compared to controls,^317^ with results differing by chronological age; aging acceleration was significantly higher in older controls compared to younger controls (P = .04), but meditators did not differ in aging acceleration by their age group. Similarly, aging acceleration and years of meditation practice variables were significantly negatively correlated for older participants (p-value = 0.03), but not younger participants. In contrast, the second study did not find a relationship between TL and time spent meditating.^83^ In their comparison of three groups, including two different meditation groups and one control group, the TL of the control group significantly differed over time. Compared to the control group, one of the meditation groups had a significantly smaller change in TL (P = .024), whereas the other meditation group did not differ from the control group. Participants in a study conducted by Earls and colleagues^112^ engaged in a wellness program involving coaching related to exercise, sleep, nutrition, and managing stress, with participants reporting significantly slower biological aging.

One study examined engagement in health activities, recreational activities, and cognitive-functioning-related activities, with all three groups of activities significantly negatively related to aging acceleration (P < .001).^180^

### Other Lifestyle Variables

Four studies included a variable related to work. Those reporting severe work-related exhaustion also had significantly shorter telomeres than those without exhaustion (P = .008),^53^ whereas perceived work stress wasn’t related to TL.^48^ Chmelar and colleagues^184^ reported on psychosocial work conditions, including time pressure, job autonomy, social support, emotion exhaustion, and burnout; results indicated that none of these predicted TL. Similarly, Weber and researchers^98^ assessed self-reported learning opportunities at work and the use of selection, optimization, and compensation strategies, but telomere was not correlated with either of these. When further divided, TL was inversely related to the use of compensation behaviours (P=.043).

Ten studies assessed exposure to toxins, of which five reported significant results and found significantly older biological aging with increased exposure of PM2.5,^318–320^ sulphur dioxide (SO2),^319^ nitrogen dioxide (NO2),^319,320^ sulfate, ammonium, and Polybrominated biphenyl (PBB),^321^ and shorter telomeres with exposure to total polychlorinated biphenyls (PCBs; P = 0.015).^322^ Three studies reported mixed results regarding PM2.5, one of which found that higher PM2.5 was significantly associated with increased EEAA, but not other measures of biological aging, and that BC and NOx, which are both indicators of air pollution from traffic, were related to accelerated biological aging for women only.^117^ Second, Gao and colleagues^323^ reported that mixed results, with increased PM2.5 exposure related to faster rates of biological aging according to the Horvath, Hannum, DunedinPoAm, and GrimAge clocks, as well as the methylation mortality risk score, but not the PhenoAge clock. Third, Pan and colleagues^324^ found that PM2.5 was significantly related to accelerated biological aging for only men, and that the relationship between aging and the components of PM2.5 varied. Three pesticides were compared to accelerated biological aging in one study,^325^ with higher levels of two of these associated with accelerated aging. Another study assessed the levels of seven organophosphate insecticide metabolite; TL significantly decreased with one, increased with another, and were not related to five metabolites.^33^ Ozone (O3) was not associated with Horvath DNAm age in one study.^319^

Overall lifestyle was assessed in two studies. In one composite lifestyle variable, Hautekiet and researchers^214^ included BMI, smoking status, physical activity, alcohol consumption and diet; results indicated that healthier lifestyles were significantly predictive of improved TL and mtDNAc. Another study sorted participants into groups based on their adolescent lifestyle (which included BMI, leisure-time physical activity, smoking, and alcohol use), with significant differences in three of the five of the biological clocks examined when comparing the groups.^203^

Three studies examined the impact of experiencing incarceration in relation to aging, with all three studies indicating that experiencing incarceration was related to significantly faster pace of aging.^119,188,280^ History of antisocial behaviour was examined in one study;^188^ this was related to a significantly faster pace of aging.

### Health outcome indicators

There were 49 studies that assayed biological age with various health outcomes. It should be noted that there was overlap between some datasets within this subsection; data from the Dunedin Study was analyzed in three studies,^153,252,255^ the HRS population in three,^120,156,158^ Lothian Birth Cohort 1936 data in three,^18,61,256^ the National Health and Nutrition Examination Survey in 2 articles,^155,258^ and the Atherosclerosis Risk in Communities (ARIC) cohort in two.116,326

### Mortality Risk

Mortality or mortality risk was analyzed against biological aging markers in 33 articles, and was most commonly compared to biological clocks in a total of 26 studies. Dependent on the clocks examined, the majority of the studies indicated a significant predictive relationship between biological age and mortality. Biological age via a novel clock was predictive of all-cause mortality in nine studies,^34,60,135,168,326–330^ including one study that also linked Reti-Age to cancer-related and CVD-related mortality.^327^ Horvath biological aging acceleration was significantly associated with mortality risk in four studies,^60,120,156,331^ but unrelated in two studies.^172,183^ Increased biological age as calculated with KDM was significantly related to increased mortality in five studies,^137,155,158,257,332^ and increased risk of cancer-specific mortality in one study.^155^ Results from one study did not support this relationship,^195^ and another study utilized KDM and reported a relationship between biological aging and mortality (P = NR).^240^

Results related to PhenoAge were significant in four studies for all-cause mortality or mortality risk,^60,158,168,183^ significant in one study for cancer-related mortality,^284^ and not significant in one study.^172^ Brain-PAD score had a significant positive correlation with mortality in two studies.^61,256^ Another three studies examined GrimAge, all of which found a significant relationship between the clock and mortality risk.^60,61,172^ Authors of two studies examined the Hannum clock and were split, with one reporting significant results^60^ and the other not reporting significant results related to mortality.^172^ The 99-CpG and 3-CpG clock were each examined in one study; mortality was significantly associated to the 99-CpG clock, but not the 3-CpG clock.^18^ Increasing difference between biological and chronological age (Δage) increased mortality in two studies, one was significant,^333^ while the other did not report significance (P=NR).^156^

Results examined non-clock variables compared to mortality produced a greater variety of results. Heightened mortality risk was significantly associated with shorter telomeres in three studies,^56,156,183^ whereas two independent researchers found that TL was not associated with mortality risk.^60,61^ Increasing scores on the frailty index significantly accelerated mortality risk,^60,331^ with similar results for the fBioAge.^52,60^ Some variables were compared to mortality in only a single study, with results including: all-cause mortality and cardiovascular mortality increased significantly with rising GDF-15,^148^ allostatic load (AL) as significantly associated with all-cause mortality,^195^ mixed results for cell type variables,^120^ and not significant findings for mitochondrial number.^156^ Gaydosh and colleagues^257^ found a positive relationship between mortality risk and both physiological dysregulation and homeostatic dysregulation, and Graf and researchers^158^ also found that greater homeostatic dysregulation was related to an increased hazard of mortality. Drewelies and colleagues^135^ examined blood parameters; 12 parameters with significantly predictive of mortality hazards, whereas 21 were not. In comparison, Dugue and researchers^108^ compared 35 blood markers to mortality; from baseline to follow-up, changes in 8 markers were significantly associated with mortality. There was a significant positive correlation between mortality and circulating histidine-rich glycoprotein profiles determined by HPA045005, but not those obtained from BSI0137.^334^ There were 14 of 58 CpG sites analyzed that were associated with mortality, two loci (cg08362785 and cg23842572) had a negative relationship with mortality risk.^183^ Srour and colleagues^296^ examined a multitude of variables, with results indicating that their association with mortality was impacted by gender; GDF-15, NT-proBNP, HbA1C and CRP were significantly related to mortality risk among men above the age of 60 but not cystatin-C, whereas only NT-proBNP, GDF-15 and HbA1C were related to mortality risk for women of the same age. Researchers of one study assessed physiological indices, with low calcium, levels of inflammatory markers, urea, liver enzymes, and glucose were associated with a greater risk of death, whereas integrated albunemia was not associated to the risk of death.^335^

### Self-Reported Health and Aging

Self-reported health and aging were assessed in eight studies. Three reports found a significant inverse relationship between self-reported health and increasing age calculated using KDM,^252,255,258^ while one article found this relationship insignificant.^257^ Individuals with a faster pace of aging had poorer self-reported health (both P<.01).^252,255^ Increasing homeostatic dysregulation had a significant inverse association with self-reported aging.^252,258^ Levine’s clock significantly increased with worse self-reported health.^258^ Levels of suPAR had a negative correlation with self-reported health (P<.01).^153^ Calculated biological age as determined by metabolic syndrome status, along with the difference between biological and chronological age were both significantly higher in those who reported worse health status.^161^ Dunedin’s Pace of Aging had a negative correlation with self-reported health at age 45.^254^ Shorter telomeres were significantly correlated with worse subjective health status in one study^258^ while another report found this association insignificant.^252^ Hannum’s clock had a significant positive relationship for “fair” health status rating, but insignificant for “poor” and “good” ratings.^116^ Belsky and colleagues^252^ found no relationship between self-reported health and the 353-CpG clock or Weidner’s 99-CpG clock, but was significantly related to the 71-CpG clock.

### Activities of Daily Living

Eight of the articles looked at the effects of aging biomarkers on ADL performance. Levine’s biological age, KDM, and homeostatic dysregulation increased significantly with more ADL restrictions.^258^ Similarly, researchers of one study found that those with incident ADL limitations had more advanced PhenoAge, KDM biological age, and greater homeostatic dysregulation (P = NR).^158^ Levels of GDF-15 had a significant inverse relationship with ADL abilities.^148^ Horvath’s biological aging clock, the difference between Horvath’s age and chronological age, and mtDNAcn heightened with ADL limitations (P=NR).^156^ In comparison, results of one study indicated that the association between ADL and biological clocks varied by both the specific clocks examined and by gender.^269^ The CpG marker in the RAB32 gene increased with ADL impairments, while the RHOT2 CpG was insignificant.^206^ TL was not associated with ADL in four reports,^78,156,258,269^ whereas frailty was significantly negatively correlated with ADL in one study.^269^ Likewise, D’Aquila and colleagues^207^ found no significance in the several CpG units for predicting ADL performance.

### Other Health Outcomes

A variety of other health outcomes were compared to biomarkers in ten reports. Life satisfaction had a significant inverse relationship with biological age as measured using the KDM,^180^ whereas TL in middle-aged women was not significant in terms of life satisfaction.^208^ The calculated difference between KDM and chronological age significantly predicted self-sufficiency in women, but not in men.^180^ The fBioAge was significant in anticipating entry into long-term care.^52^ Emergency visits and hospitalizations were examined in three study and compared to biological clocks in two; emergency visits and hospitalizations increased significantly with advanced KDM age in most of the 4 different panels of biomarkers, except for when only considering a panel nine blood biomarkers in women,^262^ and the risk of first hospitalizations increased with each year increase in biological age (P < .001) in another study.^168^ In the third, ER admissions were significantly correlated with frailty (P = 0.025), but this relationship was no longer significant after adjusting for age and sex.^336^ GrimAge and brain-PAD scores had a significant inverse relationship to attitudes about physical change, while GrimAge increased significantly with subjective psychosocial growth, brain-PAD score was not associated.^61^ Attitudes towards psychosocial loss were not significant with either GrimAge or brain-PAD score, and TL did not relate to any aging attitude measured.^61^ Quality of life was assessed in one study; a better quality of life was significantly associated with a lower biological age acceleration according to the Levine (P = .001) and Horvath 2013 clocks, but was not associated with either of the Hannum and Horvath 2018 clocks.^284^

### Chronic illness and treatment indicators

There was a total of 118 articles that assessed aging biomarkers to various chronic health indicators. It should be noted that there were overlap observed between some datasets including the following studies: HRS,^35,37,156,158,186,281^ Health In Men Study,^238,292^ Korean KNHANES,^103,161,337^ Korean Genome Epidemiology,^43,190^ Lothian Birth Cohort 1936,^61,256^ Netherlands Study of Depression and Anxiety,^38,39^ Sympathetic Activity and Ambulatory Blood Pressure in Africans (SABPA),^47,48^ ARIC,^116,326^ the CARDIA study,^137,338^ Dunedin study,^153,164,188^ Health in Men Study,^238,292^ NHANES,^31,33^ and two studies which used the same data involving heterosexual adults.^49,50^

### Chronic Conditions

Shorter TL were found in those with a diagnosis of multiple sclerosis (MS), colorectal cancer,^86^ HIV,^47,48,89,227,251^ hypertension,^73^ peripheral arterial disease (PAD)^339^, and cancer history.^73^ AD correlated with TL (P<.05).^59^ Two studies found an inverse relationship between comorbidity and TL,^39,78^ while other reports found number of chronic illnesses was not significant to TL.^37,71,156,176(p201),186,210^ One study found a negative association between LTL and multimorbidity reported by individuals,^38^ another group of researchers found this relationship significant though the directionality was not reported.^69^ McLauchlan and others^61^ found no correlation between comorbidity and LTL. In a study of Chinese patients with premature coronary artery disease (PCAD) and controls, dyslipidemia and type 2 diabetes were found to have an inverse relationship with TL only in patients, while hypertension had a similar trend in controls. Type 2 diabetes (T2D) and dyslipidemia were insignificant compared to TL in controls, hypertension was insignificant in patients, and family history of PCAD did not relate to TL in both groups.^97^ There were a number of health conditions that did not correlate to TL including: hypertension,^33,76,340^ diabetes,^33,59,73,76,340^ hypercholesterolemia,^76^ cardiovascular disease (CVD),^33,183^ cancer,^33,183^ heart disease,^59^ stroke,^59^ arthritis,^73^ and hyperlipidemia.^59^ Cardiac risks defined as hypercholesterinemia, hypertension, diabetes, nicotine consumption, coronary heart disease (CHD), and myocardial infarction in first line family members did not relate to blood sample TL.^184^

In a Chinese population, patients with CVD had decreased leukocyte TL (LTL) compared to controls.^341^ Risk scores for both stroke and CVD inversely related to LTL (P<.001).^251^ Independent studies found significantly shorter telomeres were found in individuals with hypertension,^85^ hypercholesterolemia,^228^ dyslipidemia,^190^ and diabetes.^43^ Women had longer LTL compared to men, especially those with a history of chronic health conditions (P=.03).^63^ Type-2 diabetes had an inverse relationship to LTL, this was insignificant after model adjustments.^45^ An and colleagues^35^ found a significant negative association with both stroke and cancer compared to LTL, while arthritis, hypertension, lung disease, diabetes, and heart disease were insignificant. When comparing patients with acute myocardial infarction to controls, LTL was shorter in those with hypertension, individuals with a family history of CAD were found to have longer LTL, while subjects with CAD were not significant in predicting LTL.^228^ Multiple reports supported the findings that hypertension^43,99,190^ and diabetes ^85,99,190,228(p20),238,292^ were unrelated to LTL. Sister studies reported insignificance between LTL and CVD, another report found no association between LTL and risk factors for CVD, including insulin sensitivity.^68^ LTL was not associated with hypercholesterolaemia,^43^ Parkinson’s,^99^ cerebrovascular disease,^99^ or upper respiratory infection.^50^ In a sample of African American men, common health conditions did not correlate with LTL.^169^

PhenoAge correlated significantly with diabetes,^342^ hypertension, ^160,166^ MS,^142^ incident atrial fibrillation,^342^ and scores on both the comorbidity index^281^ and Charlson Comorbidity Index.^284^ Increasing PhenoAge related to increased type 1 diabetes (T1D) duration while CVD, retinopathy, diabetic peripheral neuropathy, and cardiovascular autonomic neuropathy were insignificant; all of these health conditions were not related to Horvath’s DNA methylation age.^230^ Increased KDM age was associated with an increased risk for chronic illness.^195,326^

Physician-diagnosed chronic conditions were correlated with advanced biological aging using PhenoAge, KDM, and homeostatic dysregulation measures (P=NR).^158^ In a population of male subjects, CVD was significant when compared to PhenoAge, but not Horvath’s clock, while cancer was insignificant in terms of either biological clock.^183^ Younger amyotrophic lateral sclerosis (ALS) age of onset was correlated to decreased Horvath’s biological age.^343^ Individuals with hypertension,^128^ CHD, heart failure, or PAD showed increased Horvath’s aging.^99^ For every 5-year increase in Horvath’s age acceleration, Parkinson’s Disease onset was 6-years earlier (P<.01).^268^ Horvath’s clock was associated with diabetes in two reports,^111,342^ while another found this insignificant.^181^ Two studies found a positive correlation between CVD and Horvath’s calculated age.^113,116^ Human papilloma virus was significantly associated with Horvath’s clock.^284^ Independent studies found no relation between Horvath’s clock and MS,^142^ ischemic stroke,^344^ incident atrial fibrillation,^342^ diabetes,^128^ hyperlidemia,^128^ or myocardial infarction.^116^ Dugué and colleagues^111^ found no association between Horvath or Hannum’s clock and CVD, arthritis, hypertension, or a history of gallstones, kidney stones, and asthma. Independent reports also found that hypertension was not related to Horvath’s clock;^181^ nor was CVD associated to Hannum’s clock.^113^ Horvath’s age acceleration had a positive relationship with comorbidity in two studies (Hillmann 2023, Crimmins, 2020), while mitochondria DNA copy number was insignificant in one.^156^ Resting respiratory sinus arrythmia had a significant negative association with Horvath’s clock in extremely low birth weight survivors, though was insignificant in the sample of normal birth weight adults.^345^ Roetker and fellow researchers^116^ found significant positive correlation between Hannum’s biological aging clock and CHD, heart failure, PAD, CVD, and myocardial infarction. Diabetes diagnosis significantly affected biological aging using the Hannum clock.^342^ Comorbidity index was significantly and positively associated with the Hannum clock.^281^ Both Hannum’s clock and GrimAge were significantly associated with incident atrial fibrillation.^342^ DiffAge calculated as the difference between Hannum’s clock and chronological age, significantly associated with ischemic stroke.^344^ Diabetic peripheral neuropathy, cardiovascular autonomic neuropathy,^230^ and the number of chronic diseases^61,281^ increased GrimAge significantly. Both GrimAge acceleration and DunedinPACE had a positive relationship with hypertension.^160^ GrimAge acceleration was significant between diabetes and the years since obesity occurred, when comparing normal glucose levels, though hyperglycemia was not significant.^33^ Duration of T1D, CVD, and retinopathy were insignificant to GrimAge.^230^ Similarly, another report found diabetes insignificant to GrimAge.^342^ DiffAge, resulting from the difference between calculated biological age and chronological age had a significant association with hypertension, diabetes, heart disease, stroke, and cancer.^333^ Another study showed increasing DiffAge with T2D, hypertension, and breast cancer.^112^ Page of aging was significant to diagnoses of cancer, heart attack, and diabetes.^188^ Subjects with COPD had a significantly higher ImmunolAge.^346^ Other reports supported that calculated biological age significantly correlated with hypertension,^103^ diabetes,^103,240^ stroke, cancer,^240^ and chronic disease diagnosis or family history of.^103^ Mild Traumatic Brain Injury sustained after 60 years of age increased MRI based Brain Age by 10 years (P=NR).^133^ Pyrkov et al.,^239^ found that those with T2D had older biological ages than their peers without (P=NR). Dyslipidemia was not correlated with calculated biological age.^103^ Morbidity correlated with calculated biological age (based on nine biomarkers)^240^ and the difference between proBNP age and chronological age (P<.01).^152^ Rapp and colleagues developed a measure of physiological age, and reported that those with accelerated biological aging had significantly higher scores on the comorbidity index (P = .00).^185^ Metabolic Age acceleration was significantly higher in those with diabetes,^166,235^ ypertriglyceridemia, and a history of myocardial infarction.^235^ Subjects with diabetes had older ages, calculated using the Klemera-Doubal Method (KDM), than those without diabetes,^347^ while another report found this relationship was only significant for age groups <45 and >65.^337^ Subjects with an increased accelerated KDM age had a higher risk for stroke and CVD.^137^ Participants with obstructive CAD had an older biological age, as measured using the 7-CpG clock; after sex adjustment this association was no longer significant.^234^ Researchers of one study assessed the impact of PTSD and MDD comorbidity on GrimAge, with results indicating that there was no difference in GrimAge for those with PTSD both with and without MDD.^219^ In a participant sample of women, Horvath, Hannum, GrimAge, and PhenoAge biological aging measures were all insignificant to coronary heart disease prevalence.^274^ No significant difference in GrimAge was found between patients with hypersexual disorder and those without.^248^

Several other biomarkers were analyzed to chronic health indicators. suPAR levels significantly increased in those with one or more current health conditions.^153^ Chronic obstructive pulmonary disease was associated with lower dehydroepiandrosterone and growth hormone levels (P<.05).^243^ Those with CVD had heightened low-density lipoprotein (LDL), triglycerides (TG), HbA1C, and lower eGFR.^341^ PAD patients had lower HDL and albumin, and heightened TG, C-reactive protein, along with glucose (P<.05).^339^ Diabetes had a significant association to DNAm PAI-1.^342^ The prevalence of T2D was significantly less in SERPINE1 carriers.^348^ Brain-PAD score had a positive relationship with number of chronic diseases (P<.05),^61^ but did not correlate with CVD, diabetes, or stroke.^256^ In an elderly population, comorbidity, diabetes, and history of myocardial infarction were associated with GDF-15 levels, however hypertension, hypercholesteromelia, previous percutaneous coronary intervention, previous coronary artery bypass graft, previous acute heart failure, previous stroke, and peripheral artery disease were not.^148^ In a study that assessed 51 chronic health conditions, it was found that chronic kidney disease significantly contributed to mortality prediction.^329^ Mortality also had a significant relationship to CVD, ischemic heart disease, and cerebrovascular disease, but not cancer.^168^ GDF-15 was also significantly associated with a scale used to assess physical comorbidity burden, albeit this relationship was weak.^211^ Hair whitening score and LDL were significant in predicting CAD, while triglycerides and high-density lipoprotein were not.^349^ Compared to HIV-negative subjects, those with HIV had higher CD4 T cells, CD8 T cells, PhenoAge, GrimAge, and Horvath’s age, while Hannum’s age was insignificant.^350^ Similarly, Schantell and colleagues^351^ reported that individuals with HIV had a significantly older biological ages (P = 0.037). Expression of CDKN2A positively correlated with HIV (P < .01).^89^ Bountziouka and colleagues^199^ compared multimorbidity to frailty; the risk of frailty significantly increased with each additional medical condition (P <0.0001). St. Sauver and researchers^352^ separated their participants into multimorbidity percentiles, and reported that those with higher multimorbidity percentiles had significantly increased levels of IL-6 and TNF-a compared to their peers, but had no differences in IL-10.

### Medications and Treatments

Various biological aging markers were compared to medication use and treatments. TL had a significant inverse relationship with antidepressants.^69^ Decreasing LTL was associated with cardiac medication, anti-inflammatories (P<.01),^38^ 2 and antihypertensives (P<.02)^47,85^.

Yeap and colleagues^238^ found a positive correlation between LTL and hypertension medication. Medication use had a significant association with LTL in African American men.^169^ No significant difference was found between TL and implantable cardioverter defibrillator (ICD) therapy; however the load-of-short telomeres was significantly higher in ICD therapy individuals.^340^ Cholesterol lowering medication had an inverse correlation with LTL, however the relationship was insignificant following model adjustments.^45^ After 60 days of relaxation practices, LTL was preserved in healthy subjects, and declined in patients from a cardiology ward (P=.05).^131^ Psychiatric medication use in men from ages 20-38 years old did not relate to TL.^164^ There was no association between TL and electroconvulsive therapy in severely depressed patients.^95^ Several other treatments were insignificant compared to TL including sleep medication,^353^ psychoactive medication,^38^ hypertension medication,^292^ lipids medication,^238,292^ blood pressure medication,^45^ duration of growth hormone (GH) treatment, and cumulative GH dose.^77^ The number of medications taken in an older Chinese population were insignificant to TL.^71^ Two reports also found that medication use and TL were not related.^50,210^ In HIV patients following two years of antiretroviral therapy (ART), PhenoAge and GrimAge acceleration significantly increased, along with CD4 T cells, naïve CD8 T cells, B cells, and granulocytes.

Total CD8 T cells, exhausted CD8 T cells, natural killer cells, and monocytes had a negative association with ART,^350^ while TL and CDKN2A expression were insignificant.^89^ TL had a significant inverse association with combination ART, white matter hypertension was insignificant.^227^ Antihypertensive use significantly increased Metabolic Age Score.^235^ Two studies compared the use of antihypertensive medication to biological clocks; authors of one reported that this was associated with significantly greater odds of having accelerated aging via the PhenoAge, GrimAge, and DunedinPACE clocks,^160^ whereas the second study found that this was significantly associated with the Horvath, PhenoAge, Hannum, and PAI-1 clocks, but not with GrimAge.^342^ Polypharmacy had a positive relationship with GrimAge, inverse association with Hannum’s biological age, was significant with PhenoAge, and insignificant with Horvath’s clock in one study,^172^ and was associated with frailty in another.^336^ Increasing medication use decreased KDM age.^31^ DNA methylation age was significantly younger following 60 days of relaxation practices in controls, but not cardiology patients. Hypomethylation of KLF14 gene was significant following relaxation practices, but not ELOVL2, C1orf132, TRIM59 or FHL2; telomerase activity did not differ following treatment.^131^ Oral contraceptive use and hormonal replacement therapy in women had no association with Hannum, Horvath, Levine, or genome-wide DNA methylation age.^232^ Antipsychotic medication use was significantly positively associated with pace of aging in one study,^188^ and in another, the use of antipsychotic medication at time of death in schizophrenic patients had no association with Horvath’s biological age.^354^ Antihypertensives and anti-hyperlipidemics in African Americans did not relate to Horvath or Hannum’s clocks.^116^ Medication use was not significant in terms of CDKN2A or p16INK4a expression.^49,50^ The use of blood pressure or lipid-lowering medications were insignificant to physiological dysregulation in male pilots.^236^ Anti-inflammatory medications did not differ suPAR levels at age 45.^153^ Plasma HIV viral load had no association with TL.^227^

### Other Chronic Health Indicators

Chronic HBV infection was related to increasing biological age, calculated using 10 biomarkers, while CMV and HCV infections were insignificant.^102^ The physical health index, consisting of nine clinical indicators of poor health had a significant inverse relationship with LTL in 38 year-old men.^164^ TL did not correlate with physical illness in one study.^53^ Using KDM as a biological age measure, the difference between KDM age and chronological age had a strong relationship with disability.^262^ After controlling for covariates, a pain-related disability was predictive of advanced GrimAge.^355^ Skin conditions heavily influenced the PhotoAge Clock, developed as a biological aging measure.^356^ The Skin-specific DNAm age predictor, based on 2266 CpG sites, found a significant difference in biological age when compared to those with and without psoriasis.^357^ Psoriasis was insignificant to the difference between KDM age and chronological age.^112^ One report found five biomarkers significantly changed in women 70 years and older with breast cancer undergoing chemotherapy including IL-6, MCP-1, IGF-1, IL-10, and TNF-α; LTL and RANTES were insignificant.^205^ Using 286 breast tissue specific CpGs to estimate DNAm age, breast tumour tissue was older than normal tissue (P=NR).^358^ Breast cancer increased KDM age, while leiomyoma and glaucoma were not significant.^112^ Visceral adipose tissue and subcutaneous adipose tissue had an inverse association with TL (P<.05).^82^ Colonic tumours had significantly higher white cell counts compared to rectal tumours, while tumour site of colorectal cancer patients did not relate to LTL.^86^ In patients 70+ years of age with breast cancer, biomarkers (TL, IL-6, MCP-1, and RANTES) did not relate to tumour size or subtype.^359^ Levels of sTIL, CD3, and CD8 cells had the highest accuracy for categorizing tumour infiltration in invasive breast carcinomas.^360^ Researchers of one study compared those with HPV-related tumours to those with HPV-unrelated tumours; results indicated that those with HPV-unrelated tumors had significantly accelerated EAA (Xiao 2021). Individuals with any urological cancers, localized diseases, and metastatic diseases were found to be biologically older than their peers in one study.^361^

### Molecular biology features

There were 281 articles which compared molecular biological indicators to chronological age. More detailed information of the findings reported on molecular biology indicators and biological clocks can be found in our sister manuscript. Dataset overlap occurred as follows: Costa Rican Study on Longevity and Healthy Aging,^66,362^ Dunedin Study,^188^ FACHS,^42,162^ Framingham Heart Study Offspring cohort,^342,363,364^ HRS,^35–37,126,158,186^ Health In Men Study,^238,292^ Helsinki Birth Cohort Study,^67,226^ Helsinki Businessmen Study,^7,193^ Korean Genome Epidemiology Study,^43,190,365,366^ Leiden Longevity Study,^367^ Lothian Birth Cohort 1921,^18,114,368^ Lothian Birth Cohort 1936,^61,256,319^ Multi-Ethnic Study of Atherosclerosis,^45,46^, NHANES10,23,30,32,33,258,288–291,301,315,330,369,370 Netherlands Study of Depression and Anxiety,38–40 NAS,^371,372^ Prevention of Renal and Vascular End stage Disease,^81,192^ Social Environment and Biomarkers of Aging Study,^237,257^ Study of Health in Pomerania,^235,373^ Sympathetic Activity and Ambulatory Blood Pressure in Africans (SABPA) cohort study, UK Biobank,^198,199,374^ VITAL-DEP,^146,181^ and the Women’s Health Initiative (WHI) Clinical Trials (CT) Cohort.^233,242^

### Genetic Markers

#### Telomere Length

Telomere length (TL) was compared to aging in 155 studies. 112 of these studies found significant findings, of which 104 found a significant negative relationship,^7,10,30,33,35–40,43,45–48,50,54–67,70,72–74,79–98,100,101,164,165,169,176,178,181,184,186,190,192,204,209,210,214,223,227,237,238,244,247,251,258,288–290,303,315,322,341,348,359,365,370,371,375–388^ two studies found a positive correlation,53,68 while 6 of these studies did not report the directionality between these variables.^32,69,131,248,298,339^ Overall, 27 studies found an insignificant association between aging and TL.^76,77,99,110,157,175,177,193,199,205,208,222,228,256,259,264,270,272,293,302,340,389–394^ Three studies found an inverse correlation between TL and aging; however, the significance was not reported (Baranyi, 2022; Marriott 2023; Topiwala, 2022).^198,319,374^ Finally, nine studies found mixed results.^71,78,224,226,276,353,395–397^ Barha and colleagues^224^ found with increasing age, TL significantly shortened in women who had experienced child mortality, but not in those who had not experienced child mortality. Two studies found shorter TL with increasing age in control groups (both P<.1) but not in runners (both P>.4).^395,396^ Fang and fellow researchers^78^ found an overall inverse relationship in their study population (P<.001) but following adjustments for other factors in their final models this association was insignificant. Telomeres significantly shortened with age in men but not women in two studies.^71,353^ Kajantie and colleagues^226^ found TL decreased with age in the Helsinki Birth Cohort Study sample, while this relationship was only significant in men for the Helsinki Study of Low-Birth-Weight sample, and insignificant in the FinnTwin16 Study cohort. TL significantly shortened in older subjects, though this result was not significant in Master rowers.^276^ Men without diabetes were found to have an inverse association between age and TL, though this finding was insignificant in women and men with type 2 diabetes.^397^

#### CpG Methylation

A total of 19 articles compared various CpG site methylation and age. Predicted chronological age by CpG methylation patterns correlated significantly with donor fibroblasts chronological age.^398^ The methylation of CpG islands of ELOVL2, FHL2, and PENK were strongly related to age.^399^ Hypermethylation of three PCDHGA3 CpG sites were associated with frailty, especially cg17588578.^400^ There were 223 of 448 genes analyzed that contained a CpG related with increased aging.^363^ An increase of approximately nine years was found between biological age (determined by 4 CpG sites within genes ASPA, EDARADD, ELOVL2 and PDE4C) and chronological age (P < .001).^401^ Hypermethylation of 9 CpG sites of ELOVL2 was significant to increasing age.^402^ Daunay^403^ found a positive significant relationship between the methylation of CpGs ELOVL2, KLF14, PDE4C and TRIM59 with age, and a significant inverse association between age and CpGs *ASPA and EDARADD.* Higher chronological age correlated with the hypermethylation of a CpG unit within RAB32 and hypomethylation of a CpG unit within RHOT2.^206^ The methylation of genes ELOVL2, C1orf132, KLF14, TRIM59 and FHL2 were considered DNAmAge in one study, which showed statistical significance in positively correlating with chronological age.^131^ Sturm and colleagues^404^ assayed 867,000 CpGs, all of which had a significant inverse relationship with chronological age. From 765,808 CpG positions utilized, 4930 were significant with age in an African American cohort, while 469 were significant in a Caucasian population, only 301 of these CpG positions were consistent among both groups.^141^ Two reports considered the same eight CpG units (cg09809672 cg02228185 cg19761273 cg16386080 cg17471102 cg24768561 cg25809905 cg10917602) for significance with age, all of which were significant in one study,^383^ while the other found cg24768561 insignificant.^110^ Further, Vetter and others^383^ found that units cg16386080 and cg24768561 increased methylation with age, while hypomethylation was found in the other six units. After assessing units CpG_5, CpG_18.19, CpG_23.24, and CpG_25.26, the only significance was found in CpG_25.26, decreasing methylation with age.^207^ Using the Holm stepdown Bonferroni procedure, 232 of 484,220 CpG units showed significant changes with chronological age, while the Benjamini– Hochberg procedure found 3064 of these sites to be significant.^233^ Another report found five insignificant CpG units to chronological age of the 71 assayed.^162^ One article reported the CpG methylation of the genes ASPA and PDE4C increased over 1-year, while ITGA2B methylation did not differ.^154^ A positive association was found between age and 53% of the 2266 CpG sites from skin tissue assessed.^357^ There were 32 CpGs of 307,745 investigated which correlated strongly with facial age, the strongest significance was found in unit cg00871706.^368^

#### Specific Genes

Seventeen studies assayed chronological age and gene expression. Six studies compared CDKN2A expression with chronological age. Two studies found gene expression increases with age (both P<.05),^90,145^ while three studies found that the relationship between age and CDKN2A expression was insignificant.^49,50,91^ One study found a positive relationship between CDKN2A expression and age in HIV-negative participants, but this correlation was insignificant in HIV-positive participants (P=.17).^89^ DNA methyltransferase three alpha carriers show significant evidence for accelerated aging.^405^ A significant decrease in ELOVL2 mRNA levels was found with increasing age.^404^ Chronological age was positively associated to histidine-rich glycoprotein expression (P<.001),^334^ tripeptidyl peptidase I (P=.04),^293^ and ApoB100/ApoA1 (P=NR).^255^ With increasing age, sjTREC content declined significantly.^402^ Age was found to have an inverse relation with p16INK4a expression (P=.004),^360^ however another study found this correlation insignificant.^50^ Out of five genes (LRRN3, GRAP, CCR6, VAMP5 and CD27), all five were significant to age in the one population, while only LRRN3, CCR6 and CD27 were significant when in a different population.^406^ There were 448 and 481 genes, respectively, to be significantly associated with age in two studies.^363,364^ RNA microarray datasets from vastus laterallis muscle, brain cortex, and kidney cortex showed significant correlation with aging at two distinct ages (43 and 75), however this was not shown in the kidney medulla.^407^ There was no relationship found between NrF2 and chronological age.^145^ Age was not associated with APOE e4 carrier status.^126^

#### DNA Methylation

Five studies researched DNA methylation in relation to chronological age. McClelland and colleagues^165^ found a significant inverse association between these two variables. One study found that DNA methylation and chronological age remained constant in their strong relationship when measured during pregnancy and post-partum.^408^ Madrigano, with fellow researchers,^409^ determined a positive correlation between age and the methylation of the following genes: IFNγ, F3, CRAT and OGG, and hypomethylation of GCR, iNOS, and TLR2 genes with age. One model determined by Lin and others^18^ used over 2100 DNA methylation profiles in determining a predicted age that was highly related to chronological age. The final article showed no association between chronological age and genome-wide DNA methylation.^232^

#### Specific Cells

Twelve articles compared specific cells to chronological age. Five of these studies were significant; three determined a negative relationship between CD8+/CD4+ cells and age,^114,138,221^ while two studies^219,360^ found a positive association between naïve CD8+ cells and age. McEwen and fellow researchers^362^ also found an inverse correlation between CD8+ cells and age, although the significance is not reported. Age advancement was associated with increased CD8+T cells in another study (P = NR).^102^ A positive correlation was found between CD28-T cells and age (P<.01).^219^ Additional results show a significant inverse relationship between age and Natural killer cells and increasing B cells with age.^221^ Prediction of age was younger by a significant 4.15 years on average when considering CD4+ T cells instead of CD14+ monocytes.^142^ Zhu^410^ found 9 cell subtypes of 29 studied to be significant to chronological age including: NK-GZMH, CD8-CTL, CD8-Naive, CD4-Tm, CD4-Naive, Naive-B, NK-FCER1G, CD14-MC, and Memory-B-CRIP1. One study found the correlation between age and CD4+/CD8+ counts to be insignificant.^411^ Another study found mixed results, as strong correlations were observed between change in DunedinPACE and change in proportion of CD8+ T cells, CD4+ T cells, natural killer cells, and B cells, but not monocytes.^167^ Berg and researchers^280^ (2022) reported mixed results due to finding significant negative correlations between CD8+ T cells, CD4+ T cells, B cells and Monocytes with accelerated GrimAge, but no significant correlation was observed for natural killer cells.

#### Other Genetic Markers

Several other genetic markers were compared to aging in 29 studies. Mitochondrial copy number was assayed to chronological age in seven studies; four of these studies found that mtDNA copy number levels decreased significantly with chronological age.^92,214,371,382^ Alternatively, three other studies found no significant relation between these variables.^146,181,318^ Five studies assayed terminal restriction fragment length (TRF) and age. Four of these studies resulted in an inverse association between the two variables, two of which were significant,^348,412^ and two did not report significance.^353,381^ Alternatively, one article found TRF increased significantly with chronological age.^265^ Two studies assessed telomerase activity and aging; both of which found inverse relationships between the variables (P = NR)^272^ (P=.0016).^377^ Four studies assayed miRNAs to chronological age. Berben and fellow researchers^360^ found significance for multiple miRNA’s including miR-326, although the directionality is not reported, along with a positive relationship for miR-18a and miR-19b, and negative association between age and miR-155. Another study reported 37 of 175 miRNA’s analysed were significantly different in expression for younger vs. older samples, however this difference was no longer significant following Bonferroni corrections.^413^ Waaijer and colleagues^414^ found that miR-633 levels were only age-dependent under stressed conditions, significantly increasing with age. Expression of miR-21-5p and miR-126-3p significantly increased in age, except for the ulracentennarians group, while miR-146a-5p and miR-181-5p were insignificant.^147^ A significant positive correlation was found between chronological age and Cdc42 mRNA level.^398^ Carriers of SERPINE1 allele showed increased lifespans compared to those without (P = .03).^348^ The healthy aging gene score, made up of 150 RNA markers, increased significantly with age across various brain regions.^415^ Both Rs4746720 and Rs3758391 had a positive relationship with age (P<.01).^271^ Contini and colleagues^273^ found 69 of 79 assayed salivary proteins and peptides significantly decreased in elderly subjects. Beta-2 microglobulin increased significantly with age.^326^ There were 19 biomarkers analyzed that were significant to age, and composed a biological aging model.^416^ A positive correlation between DNA damage in white blood cells and age was found (P = NR).^272^ Two single nucleotide polymorphisms were significantly associated with accelerated aging, including rs11668344, however rs16991615 was insignificant.^242^

Plasminogen activator inhibitor-1 was assessed to age in three studies; Lind and colleagues^113^ found a significant positive correlation between the variables, while other articles showed there was no significance to the relationship.^47,417^ Of the 22 Senescence-associated secretory phenotype (SASP) markers, only 17 were significantly associated with chronological age following model adjustments.^417^ Mitochondrial haplogroup clusters were not significant in terms of aging.^318^ Oxidative stress parameters malondialdehyde, paraoxanase, and Trolox equivalent antioxidant capacity did not correlate with age.^147^

### Blood Chemistry and Nutrients

#### Cholesterol

A total of 29 articles compared the levels of cholesterol to chronological age. Total cholesterol increased with age in 12 articles,^23,92,134,150,165,291,326,333,347,377,387,418^ had a negative relationship with age in 5 studies,^152,173,240,266,411^ while two studies were insignificant.^139,419^ One study found an inverse association between total cholesterol and LDL with age, this result was only significant in men, whereas total cholesterol and LDL increased with age in females.^107^ Low-density lipoprotein (LDL) levels significantly heightened with age in seven reports,^92,134,150,333,373,387,418^ decreased with increasing age in one study,^266^ and was insignificant in three.^377,381,419^ Reports of high-density lipoprotein (HDL) interestingly showed four positive^23,92,134,418^ and four inverse relationships to age,^188,333,381,420^ although 4 additional findings were insignificant.^150,301,377,419^ Zhong and fellow researchers^266^ found that HDL significantly increased with age in males, but not in females. Two studies also found that both total cholesterol and HDL increased with age, the significance was not reported.^255,291^ Another study showed HDL and total cholesterol increasing and then decreasing throughout the lifespan.^277^ Another study reported that cholesterol was among the seven most important biomarkers for aging in Canadian (HDL), South Korean (total cholesterol), and Eastern European (LDL) populations.^330^ There was no significant difference found in HDL/LDL ratio between fast and slow aging groups in Sternang and colleagues’^261^ research.

#### Blood Glucose, Insulin, Triglycerides, and Related Components

Thirty-two studies assessed blood glucose, insulin, triglycerides, and related components. Fasting blood glucose significantly increased with age in six articles, ^150,173,277,333,418,420^ and was reported insignificant in six other studies.^92,233,381,411,421,422^ Mamoshina and colleagues^139^ determined that fasting blood glucose had the strongest correlation to age compared to other biological indicators in their study. Glucose significantly increased with age.^107,326^ Triglycerides had a significant positive relationship with age in six articles,^92,150,235,277,333,420^ although four reports showed insignificant associations between the variables.^134,261,377,387^ Four articles found a negative correlation between triglycerides and age, although this was only true for men in three studies^107,266,373^ and another did not report the significance of this inverse relation.^255^ Al-Daghiri and others^397^ reported that triglycerides predicted 18% of the variance in TL (P < 0.001). Brunt and researchers^134^ assessed circulating interleukin-37 (IL37) serum levels and found that the IL37:IL6 ratio was inversely correlated with fasting blood glucose levels. Triglyceride-glucose index decreased significantly with increasing age.^233^ Blood glucose was found to be insignificant compared to age in three articles.^134,152,261^ Banszerus and colleagues^234^ reported that triglycerides were not associated with DNAm Age. Bahour and colleagues^347^ found that hemoglobin A1C percentage was strongly correlated with increased BA in participants with T2D (P < .0001) Triglycerides and blood glucose were rated in the seven most important biomarkers to determine biological age in one report.^330^ Insulin resistance was significantly related to cellular aging in one study.^370^ Fasting insulin and HOMA-IR had a significant positive association with age in one study,^233^ while three studies reported insignificance between age and insulin as well as HOMA-IR.^134,377,422^

#### Other Blood Chemistry

Various blood chemistry components were questioned as determinants of age in 36 papers. Glycsylated hemoglobin (HbA1C) was found to have a positive significant association with age in ten studies,^23,92,235,291,301,326,347,377,411,423^ and decreased with age in one study (P = NR).^255^ One study reported age to significantly increase with HbA1C, however this was only significant in men, not women.^373^ Another reported noted that HbA1C had the largest significance in their age-predicting model.^139^ Kristic and colleagues^424^ found that galactosylation levels had a strong correlation to age. Age was moderately associated with HbA1c in another study,^296^ while insignificant in another.^422^ It was observed in one article that urea nitrogen and age have an inverse relationship (P=NR),^255^ while ten papers reported a significant positive association^23,240,291,301,326,337,347,381,411,418^, and one study did not report directionality of this association.^308^ Similarly, urea also increased significantly with age,^377,421^ this positive correlation was also determined in a third study, though the significance was not reported.^330^ Holly and fellow researchers^406^ stated a significant difference in urea levels in older and younger cohorts. One study found urea to be insignificant to chronological age.^150^ One study showed an increase in folate as we age (P = NR),^372^ one found a significant association between the variables^308^ while two other reports found folate to be insignificant with aging.^92,421^ Fibrinogen levels significantly heightened with age in two studies,^47,377^ while another report found significance in men but not in women.^373^ Vitamin B12 was significantly higher in older subjects.^92^ Another report found the relationship to be insignificant between B12 and age.^261^ Vitamin B6 and B12 were reported to have the weakest correlation with age compared to other variables in another study.^372^ Calcium was found to decrease with age,^421^ although a separate finding showed this insignificant.^261^ Sodium was found to be one of the top seven most important biomarkers for aging in a South Korean population,^330^ although was found to be insignificant when compared to age in two other reports.^381,421^ Uric acid had a positive correlation with age (P<.001),^107,326,418^ however the relationship was insignificant in two studies.^150,381^ Another report found uric acid to decrease with age (P = NR).^173^ Homocysteine had a significant inverse association with age,^261^ while two other reports found this correlation positive.^92,372^ Apolipoprotein A1 (App-A1) had a significant increase with age, while apolipoprotein B (ApoB) decreased in aging males, and increased in older females.^107^ Langevin^188^ found aliopoprotein B100/A1 ratio to decrease with age. Plasma protein profile consisting of 77 plasma proteins was within five years of accuracy in predicting chronological age;^231^ another study found that total protein levels were inversely related to age (P<.0001).^421^ In one report the von Willebrand factor was found to have a positive association with age,^377^ while a contrasting finding showed insignificance between the variables.^47^ Ferritin was found to increase significantly with age,^32,337^ similar to C-peptides,^377^ serum markers of fibrosis,^425^ serum phosphate,^165^ and inorganic phosphate.^421^ Estimated glomerular filtration rate and vitamin D both significantly decreased with age in two independent studies.^165,333^ Circulating bone marrow-derived progenitor cell counts also had an inverse relationship with age (P = NR).^55^ Four of 22 blood metals were found to be significantly associated with age, arsenic showed a positive relationship while vanadium, cobalt and zinc were negatively correlated with age.^426^ Potassium was not significant when compared to aging.^377,381,421^ Transferrin also had an insignificant association with age.^418,419^ Five other biomarkers were found to be insignificant when compared to age in independent studies including: globulin,^381^ free T4 cells,^421^ prealbumin,^418^ cdc-42 protein levels,^398^ and aldosterone.^377^

### Endocrine

There were 12 articles that compared aging to endocrine related biomarkers. Three of these studies found IGF-1 to decrease significantly with age.^238,377,411^ The strongest biomarker associated with age was IGF-1.^359^ Additionally, IGFBP1 increased with age, while IGFBP3 declined (both P<.001).^238^ Karametos and colleagues^243^ found that dehydroepiandrosterone (DHEA) and growth hormone (GH) also had a significant inverse relationship with chronological age. Thyroid stimulating hormone was found to have a significant positive association with age in one study,^421^ however another article found this correlation insignificant.^261^ Waking cortisol levels increased with age, however the rate of diurnal cortisol decline was insignificant.^149^ Two studies found cortisol insignificant compared to age.^345,422^ GDF-15 was higher in older geriatric patients (P<.01), this was finding was supported in 2 other studies,^211,366^ however in a multivariate analysis, age was not an independent factor for GDF-15.^148^ Srour^296^ found GDF-15 to moderately associate with age. Similarly, IGFBP-2 had a positive significant relationship with age.^366^ Testosterone, calculated free testosterone, dihydrotestosterone, and oestradiol all decreased with age, while sex hormone binding globulin increased.^292^ Another article contradicted this, finding testosterone insignificant in terms of age, while estradiol increased significantly in women only.^422^ One study reported insignificance between IGF-1 and age.^205^

#### Immunology and Inflammation

Various immunological and inflammatory biomarkers were compared to chronological age in 46 studies. Hemoglobin was found to have a significant negative relationship with age in eight articles,^23,152,261,266,277,333,381,421^ while two other studies reported decreasing hemoglobin levels with aging (P=NR).^291,330^ Alternatively, a separate paper found a significant positive association between hemoglobin and age.^92^ Hemoglobin’s correlation with age was insignificant in two studies.^301,418^ C-reactive protein (CRP) was found to increase with age in eight articles,^23,107,134,240,301,326,363,381^ decrease in two reports,^64,411^ while the relationship was insignificant in 6 other studies.^147,152,277,406,418,422^ Additionally, Rahman and Adjeroh^291^ found an inverse correlation between age and CRP, while Schrempft^173^ found CRP increased positively with age (P = NR). One study found CRP and white blood cell counts to remain stable with age.^188^ Finally, Srour^296^ found CRP to moderately associate with aging. There were five findings of a negative association between white blood cell count and age,^23,255,291,301,398^ while three studies found this relationship insignificant.^381,411,421^ Zhong and colleagues^266^ found that white blood cell count increases significantly in females, but not in males. Red blood cell count was found to have a significant inverse correlation with age.^23,266,301,381^ Another report confirmed this trend; however, the significance was not reported.^291^ Florian and fellow researchers^398^ found that red blood cell count was insignificant to chronological age. Red cell distribution width increased significantly with age.^277,411^ In older adults, MMP-9 increased significantly.^427^ Lymphocytes were found to decrease with age,^291^ the significance of this relationship was reported in 4 studies.^23,237,266,301^ Lymphocyte percentage, hematocrit, hemoglobin, and monocyte count were significant in terms of aging in one report, directionality was not reported.^308^ Hematocrit had a significant negative association when compared to age.^23,266^ Another study confirmed this trend (P = NR),^291^ and three other reports found the correlation insignificant.^301,406,418^ Neutrophils increased significantly with age.^237,277^ Zhong and colleagues^266^ reported this was only significant in females. Monocytes had a significant positive relationship with age.^266^ Platelet counts decreased with age in two studies^23,291^ and was insignificant in two other reports.^301,381^ Interleukin 6 (IL-6) had a significant increase with age in eight studies.^134,165,277,352,359,363,366,411^ Holly and fellow researchers^406^ found a significant difference in IL-6 levels when comparing older and younger cohorts. One report showed the relationship between age and IL-6 to be insignificant.^64^ Interleukin 1A had a significant inverse correlation with age, while IL-17A was significantly positive when compared to age.^360^ Two studies found IL-10 to increase significantly with age,^352,411^ while two other findings showed this association insignificant.^205,366^ An inverse relationship was found between IL-37 and age.^134^ Lin and colleagues^363^ found that as age increases, p-selectin, ICAM1, osteoprotegrin, TNRF2, and MCP-1 significantly increases as well. Koh^427^ supported the positive relationship between age and MCP-1. Another report found that MCP-1 had significant age-related changes,^359^ while the sister study reported the relationship insignificant.^205^ Another study found MCP-1 also insignificant with aging.^366^ Age was associated with increased TNF-alpha levels,^64,205,352,366^ though one report did not support the significance of this association.^134^ Crimmins and fellow researchers^411^ found CMV and TNRF1 to have a significant positive relationship with age, while TGF-beta significantly decreased.

Reactive oxygen species and total antioxidant capacity were significant in increasing with age.^97^ Similarly, mean cell volume had a positive significant correlation with age.^152,411^ Erythrocyte sedimentation rate increased significantly with age,^152^ along with Plasma suPAR levels.^153,427^ Global white matter integrity and GAL-9 significantly declined with increasing age.^130,360^ Cytomegalovirus optical density increased with age (P=NR).^240^ Of the 22 immunoglobulin G (IgG) glycans analyzed, 19 of them were found to be significant in terms of aging, 10 IgG glycans were significant in both men and women.^150^ Five of 14 biomarkers were selected to compute a biological age due to significant correlation with chronological age: natural killer activity, lymphoproliferation, neutrophil chemotaxis, neutrophil phagocytosis, and lymphocyte chemotaxis.^217^ The following biomarkers did not significantly correlate with age: mean corpuscular volume,^421^ IL-1Ra,^366,411^ CM-CSF, IFN-gamma, IGFBP-7, IL-5, IL-6, IL-7, IL-8, IP-10, IL-15, IL-17A, MIP1Beta, TIMP1, TIMP2, VEGF-a,^366^ Regulated on Activation, Normal T cell Expressed and Secreted (RANTES),^205^ leukocytes,^152^ LP-PLA2 mass and LP-PLA2 activity.^363^

### Hepatic variables

There were 25 reports comparing hepatic biomarkers and age. Albumin was found to significantly decrease with age in nine studies.^23,107,152,266,301,326,381,411,421^ One study found a significant positive association.^347^ An inverse relationship between albumin and age was found in three studies (P = NR).^277,291,330^ Holly and fellow researchers^406^ found a significant difference in albumin levels between older and younger cohorts. Albumin and age had a significant correlation in one study.^308^ Albumin was not significantly correlated with age in two findings.^240,261^ Glycated albumin and fructosamine had a significant positive relationship with age.^326^ AP increased significantly with age.^23,240,301,421^ One report found AP to increase with age (P = NR).^173^ Rahman and Adjeroh^291^ found a weak positive correlation between AP and age.

Another report contradicted this, finding that AP and age had a significant inverse association.^411^ Aspartate transaminase (AST) increased with age significantly.^150,333,418^ Three studies found this correlation insignificant.^139,381,421^ Alanine transaminase (ALT) had a significant positive relationship with age in one study,^150^ and a significant negative association in another.^418^ Kang and colleagues^333^ found ALT increased significantly in women and decreased in males with increasing age. One finding showed ALT and age had an insignificant correlation.^381^ Gamma-glutamyl transpeptidase and age had a significant positive relationship in one report,^333^, another study found an inverse association,^107^ while other researchers found this association insignificant.^381^ Age and CDC42-GTP had a significant linear relationship.^398^ Glutamic oxaloacetic transaminase increased significantly with age.^337^ Hepatic collagen content was significantly correlated with age acceleration.^425^ ALT had a significant inverse association with age, while aspartate aminotransferase had a positive relation to age.^107^

### Renal variables

A total of 26 studies assayed renal markers and chronological age. Eight reports found a significant positive association between creatinine and age.^92,107,150,152,240,326,347,421^ Zhong and colleagues^266^ found creatinine to decrease significantly with age. One article declared a significant finding between the variables without stating directionality^308^ while another found the relationship of creatinine and age to be insignificant.^381^ Serum creatinine increased significantly in older subjects.^23,333,411^ Urine creatinine and age had a significant inverse correlation.^337^ With each decade, creatinine clearance decreased (P<.01). ^173,277^ Cystatin C and age had a significant positive association.^107,265,271,326,411,418^ One finding showed a moderate association between cystatin C and age.^296^ Estimated glomerular filtration rate decreased with age (all P<.05).^92,165,266,326^ One study found urine protein to increase significantly with age,^333^ while another group of researchers found urine protein to decrease with age (P<.01).^418^ Albumin−creatinine ratio (ACR) was found to be significantly associated with age in one report,^235^ and insignificant in another.^92^ Hertel and fellow researchers^235^ also found microalbuminuria to have a significant relationship with age. Urine red blood cells and age had a significant negative correlation, whereas bilirubin increased significantly, and urine white blood cells were not associated with age.^418^ Levels of urinary iPF2alpha-VI had an inverse correlation to age while 8-deoxyguanosine (8-oxoG) and 8-dihydroguanosine (8-OHdG) increased significantly in females only, and DTyr was insignificant.^229^ One study indicated that eGFR was not related to biological age.^234^ Neither urine malondialdehyde^376^ or urine cg^421^ had a significant relationship with age.

### Cardiovascular and Respiratory

Cardiovascular and respiratory biomarkers were assayed in terms of chronological age in 8 studies. With increased aging, N-terminal fragment B-type natriuretic peptide precursor significantly increased.^152,411^ Respiratory sinus arrhythmia declined significantly with age.^345^ Peak flow and age had a significant inverse association.^411^ D-dimer was found to increase with age (both P < .05). ^165,265^ Another report found this relationship insignificant.^47^ Higher age increased risk for developing atrial fibrillation.^342^ Carotid-femoral pulse wave velocity associated significantly with age.^218^ Of 109 cardiac indicators screened for association to chronological age, carotid vascular and cardiac ultrasound, pulmonary function, and atherosclerotic indices were the most frequently selected.^428^ Pulse pressure have a positive relationship with age, while pulse transit time decreased (P = NR).^429^

### Other Biological Indicators

Homeostatic Dysregulation, as measured using the Mahalanobis distance method, was found to have a significant positive relationship with age in 4 studies.^257,258,306,369^ Belsky and colleagues^283^ found that homeostatic dysregulation increased in individuals with a greater pace of aging at 38 years old (P < .001). With increasing age, Graf and colleagues^158^ found greater homeostatic dysregulation (P = NR). One study found a moderate association between age and homeostatic dysregulation (P = NR).^195^ AL was found to increase significantly with age.^369,430^ One study had a similar finding though significance was not reported.^195^ Physiological dysregulation also increased with age.^236,332,369,430^ Physiological dysregulation also increased with age.^236^ The chronological age at death and the tooth annulation method age estimation had a significant positive correlation.^431^ Additionally, there were several other biomarkers that showed a significant positive relationship with age including: adiponectin,^165^ white matter hypertension,^227^ plasma Fetuin-A concentration,^86^ follicule stimulating hormone,^432^ and ultra-weak photon emission of skin.^151^ Ocular biomarkers were measured in two sister studies, which found lens density to increase significantly with age.^90,91^ Both arteriolar and venular diameters of the retinal vessel calibre had an inverse association with age in one of the articles,^91^ while the other found only the arteriolar diameter to be significant in terms of aging, decreasing until the age of 50, then increasing with age.^90^ The retinal nerve fibre layer thickness also had a significant negative relationship with age,^90^ the other report found all measurements except for nasal to be consistent with this finding.^91^ For corneal endothelial cell parameters, endothelial cell density (ECD) and coefficient of variation (CV) decreased significantly with age, while the hexagonality index (Ex) increased.^90^ the other report found all measurements except for nasal to be consistent with this finding.^90^ For corneal endothelial cell parameters, endothelial cell density (ECD) and coefficient of variation (CV) decreased significantly with age, while the hexagonality index (Ex) increased.^90^ Alternatively, the sister study found ECD to increase with age, CV decreased, and Ex was not significant in terms of age.^91^ Wiweko and fellow researchers^432^ found a significant inverse correlation between both anti-Mullerian hormone (AMH) levels and antral follicle count (AFC) in comparison to age. Metabolic syndrome was significant when compared to the difference between biological and chronological age.^113^ Both auricular surface and pubic symphysis showed a strong positive association with chronological age.^431^ Active cell mass was found to decrease in women with accelerated aging.^433^ There were 30 of 34 biological indicators found to associate with age in one study.^108^ Of the 71 metabolites with a sex-specific regulation evaluated by Sol,^434^ 37 were significant in terms of aging, regardless of age. Levine^23^ found a positive relationship between age and cytomegalovirus optical density, an inverse association with forced expiratory volume, while granulocyte percent was not significant to age. The metabolic age model, as measured using nine metabolic markers correlated well with chronological age (P = NR).^166^ The presence of DNA damage foci and damaged telomeres were positively associated with chronological age of cultured fibroblast donors (P < .01); however, the presence of micronuclei was not related to age.^367^ Brain-PAD scores did not correlate significantly with chronological age.^61,256^ Similarly, basal fat oxidation was insignificant when compared to age.^435^

### Physical health indicators

A total of 225 studies examined physical health variables. Of these, 77 had dataset overlap, including: Dunedin,153,189,252,255,283,436 NHANES,^10,23,30,31,144,155,258,291,369,370,437^ Berlin Aging study,^75,269,314,438^ Sister Study,^160,297,439^ PREVEND,^81,192^ pSoBid,^96,165^ CARDIA study,^338,382^ SLAS-2,^180,266^ SER,^224,389^ a subpopulation of the China Kadoorie Biobank,^107,197^ Sympathetic Activity and Ambulatory Blood Pressure in Africans study population,^47,48^ HRS population,^35,36,104,120,411^ UK Biobank,^199,374^ Lothian Birth Cohort,^61,191,256,368^ VITamin D and OmegA-3 TriaL-Depression Endpoint Prevention study,^146,181^ MESA,^45,46^ MAPT,^278,440^ National Health and Resilience in Veterans Study population,^176,202^ Netherlands Study of Depression and Anxiety population,^38,39^ Korean Genome Epidemiology Study population,^43,190,366^ Korean KNHANES study,^103,161,337^ Health in Men Study population,^238,292^ European Prospective Investigation into Cancer (EPIC) study,^178,296^ two that utilized data from 139 Han Chinese participants,^265,412^ and two that used data with the same heterosexual adults.^49,50^

### Anthropometric variables

#### Body proportions

Twenty-three studies examined the relationship between body proportions and biological age. Of those 23 studies, researchers of 14 studies reported that higher body fat,^302,373^ lower muscle mass,^30,75^ greater hip circumference,^231^ greater waist circumference,^113,197,233,296,370,374,424^ greater A Body Shape Index (ASBI) and greater waist-to-height ratio,^416^ and greater waist-to-hip ratio^113,297^ were all associated with greater biological age. Three studies showed no significant relationship between body proportions and biological age,^68,262,266^ and six reported mixed results.^77,96,150,190,382,394^ Of the mixed results, two studies found a significant positive relationship between biological age and lean muscle mass, but not body fat percentage.^77,394^ Revesz and colleagues^382^ found that greater waist circumference was associated with greater DNA methylation and telomere attrition over a 10-year period, but not TL. Results from Shiels and colleagues^96^ showed that waist and hip circumference was associated with TL for a low SES sample but not significant for high SES. Yu and colleagues^150^ reported that waist circumference was significant associated with greater biological age for the full sample, but not for males or females individually, whereas Shin and researchers^190^ found a negative relationship between waist circumference and waist-to-hip ratio and biological age for men but not women.

Twelve studies compared body proportions to chronological age. Seven of these papers reported that greater chronological age was significantly associated with higher waist circumference,^337,373,377,420,441^ higher thigh circumference,^266^ higher waist-to-hip ratio,^377^ higher body fat percentage,^337,442^ and higher soft lean mass.^337,442^ Researchers from one study reported no association between body proportions and chronological age,^92^ and researchers from three studies reported mixed findings.^277,433,443^ Of the mixed results, Negasheva and colleagues ^433^ found that women over the age of 90 had higher hip-to-waist ratios, and lower fat and muscle mass, but no difference in waist circumference as compared to women aged 60 to 90. Results from Kuo and authors^277^ showed that waist circumference increased with chronological age and lean body mass increased to mid-life and then steadily decreased; mid-thigh muscle decreased after age 50 for men at a steeper rate than women, and total fat mass increased until age 60 for women and age 80 for men, subsequently decreasing. Another study indicated that, for women, lean body mass was negatively correlated with chronological age, and body fat, waist circumference, hip circumference, and waist-to-hip ratio were all positively correlated with chronological age; for men in the same study, lean body mass and hip circumference were negatively correlated with chronological age, and waist circumference, waist-to-hip ratio, and body fat were positively correlated with chronological age.^443^

#### Body mass index

Ninety-nine studies compared BMI to biological age; of those studies, authors of 42 studies reported that a higher BMI was significantly associated with greater biological age or indicators of biological aging,^10,39,47,48,53,72,81,82,103,104,111,113,128,134,153,155,160,161,192,197,202,219,229,232,233,235,238,240,272,292,293,296,297,342,369,370,373,374,397,424,437,444 37^ found no relationship between BMI and indicators of biological aging,^35,43,46,49,50,63,70,73,77,93,95,97,98,146,147,152,157,171,172,176,178,181,190,193,210,211,226,227,234,237,243,262,309,313,339, 349,366,390,395^ and researchers of 21 studies reported mixed results.^31,35,36,54,55,83,85,111,166,179,182,203,211,230,244,284,345,347,353,422,443^

Of the mixed results, seven studies showed gender differences.^31,35,36,111,353,422,443^ Four studies reported that BMI significantly predicted biological age (*p* < .01), but that this effect was stronger for males than other genders.^35,36,111,353^ In contrast, two studies found the inverse: a significant association between higher BMI and greater biological age for women (p < .001), but not for men.^85,443^ One study found that BMI was correlated with both perceived aging and perceived age among women, but only with perceived aging for men.^422^ Five research groups^166,179,182,230,244^ reported that higher BMI was related to some indicators of biological aging but not others. Other findings include that TL was associated with weight for only select weight categories,^54^ the relationship between BMI and LTL was not significant after controlling for other variables,^55^ and that this relationship existed at only select time points in one study.^83^ Levine and Crimmins^31^ evaluated the relationship between changes in BMI and changes in biological aging over time and found that increases in BMI over time were significantly associated with increases in biological age and that this effect was greater for adults with greater chronological age; this effect disappeared, however, after controlling for gender differences.

Furthermore, if BMI did not change over time, individuals showed a decrease in biological age. Results of one study indicated that increased BMI was associated with older biological ages for participants without diabetes, but this relationship was not found among participants with diabetes.^347^ Kankaanpaa and colleagues^203^ reported that the relationship between BMI and biological clocks was significant for most analyses, but this varied by the clock examined and which covariates were included. The correlation between GDF-15 and BMI was significant only among participants in a late-life depression group in one study.^211^ Mathewson and colleagues^345^ also examined BMI, and reported that the relationship between BMI and biological aging varied by group and sex. Results of one study indicated that BMI and biological aging had a nonlinear relationship.^284^

Ten studies compared BMI to chronological age. Results of two studies showed that lower BMI was significantly associated with higher chronological age,^82,433^ and three studies reported that higher BMI was significantly associated with higher chronological age.^173,377,442^ Three studies did not report a significant relationship^92,349,387^ and two reported mixed results.^277,337^ Of the mixed results, Jee and colleagues^337^ showed that increases in BMI were associated with chronological age for women but not men, and Kuo and researchers^277^ revealed that BMI increased from early to late mid-life, then subsequently decreased as chronological age increased.

### Height

Seven studies compared biological age to height, two of which found a significant negative relationship between height and biological age,^231,443^ and five of which found no relationship.^68,77,111,266,293^ Four studies also compared height to chronological age; two of these studies showed that height was significantly negatively associated with chronological age,^266,433^ one reported a positive relationship,^373^ and one reported no association.^349^

### Obesity and weight change

Researchers in 18 studies explored the relationship obesity and weight changes had with biological aging. Of the 15 studies, nine reported that obesity was associated with higher biological age96,120,168,189,231,293,338,429,437 and six found no association.66,68,181,224,266,397 A study by Cao and researchers^437^ found significant associations between biological aging and absolute weight changes. Another study also reported that weight loss increased per standard deviation of TL loss.^199^ Nwanaji-Enwerem and colleagues^385^ found that weight loss had no significant effect on any epigenetic aging measures. Five studies also compared obesity to chronological aging, where three found that obesity was positively associated with higher chronological age^92,266,377^ and two did not report a significant relationship.^349,433^ Two studies also compared weight loss or gain to biological age; Ng and colleagues^180^ reported that unintended weight loss significantly predicted greater biological age, and Belsky and researchers^306^ found no association.

### Frailty and physical functioning

#### Frailty

Of the 34 studies that examined the frailty as an indicator of biological age, 20 found that higher frailty was significantly associated with greater biological age,^34,52,90,148,185,191,197,199,213,225,262,313,331,336,360,361,400,417,421,445^ seven were not significant,^71,90,91,204,269,393,413^ and seven reported mixed results.^60,172,204,205,251,359,446^ Four of the mixed results compared frailty with TL. Mehta and colleagues^251^ found that frailty was significantly associated with TL (P = .02) when unadjusted, but this association was no longer significant when covariates were controlled for. Arts and researchers^204^ reported that TL was significantly associated with frailty, but was not predictive of frailty in a 2-year follow up; additionally, all associations became non-significant after adjusting for chronological age.

Results from one study indicated that TL was associated with frailty but no other biomarkers.^205^ The fourth study reported no association between TL and frailty, but found a significant positive correlation between IL-6 and frailty (p = .01).^359^ Li and colleagues^60^ found that frailty was correlated with DNA methylation (r = .24), but not TL. Three research groups that reported mixed results compared frailty with DNA methylation. McCrory and colleagues^172^ found that DNA methylation with GrimAge was associated with higher frailty (P = NR), but DNA methylation with PhenoAge was not associated with higher frailty once adjusted for other variables. Both Kim and colleagues^446^ and Gale and colleagues^191^ reported that DNA methylation was positively associated with frailty (P < .01 for both), but chronological age was not.

The authors of five studies looked at chronological age in relation to frailty, where one reported a significant positive association,^204^ two were not significant,^172,191^ and two reported mixed results.^446,447^ Of the mixed results, Arosio and colleagues^447^ reported that the association between frailty and chronological age varied across their centenarian, control, and offspring groups, with only the offspring group producing a significant relationship (P = .020). The other mixed result showed a significant relationship between frailty and DNA methylation which was dependent on chronological age, and became insignificant after adjusted for this.^446^

### Physical Functioning

Thirty-five studies compared physical functioning (such as mobility, balance, fatigue, gait, locomotion, or handgrip strength) to biological age; authors of 20 found a significant relationship between lower physical functioning and greater biological age,^60,73,102,120,153,185,189,199,215,251,255–257,262,266,278,373,440,448,449^ five found no significant association,^204,207,269,276,431^ and ten reported mixed results.^52,61,172,252–254,258,355,394,441^ Of the mixed results, Beam and associates^253^ reported that physical functioning was correlated with biological age but functional ability was not. Greater biological age was associated with decreased physical functioning in all domains except for grip strength.^254^ Belsky^252^ found that all physical functioning was associated with KDM biological age and pace of aging, but not with TL or DNA methylation. Finkel and colleagues^52^ reported sex differences in the association between physical functioning and biological age, where females experienced a greater increase in biological age with lower physical functioning than males. Golab and colleagues^441^ tested biological age against a variety of physical functioning measures for groups of 20 and 30 year olds; significant differences in balance and sit-and-reach task for 30 year olds by biological age but not 20 year olds, no difference in the tapping test and biological age between 20 and 30 year olds, and significant differences in broad jumping and grip strength as compared to biological age for both the 20 and 30 year old groups. McCrory and researchers^172^ found that DNA methylation was associated with slower walking speed and lower grip strength (P < .01, P < .001; respectively), but these significant relationships disappeared after multivariable adjustment. McLachlan and colleagues^61^ compared walking speed to DNA methylation (P = .016), not significantly associated with TL or brain age. Peterson and colleagues^355^ found that DNAmGrimAge was associated with balance, but not sit to stand, walking, or total SPPB scores. Lower grip strength, but not walking speed or knee extension strength, was associated with higher age acceleration, and TL was not associated with physical functioning in one study.^394^

Researchers of 12 studies compared physical functioning to chronological age; eight of these groups of researchers reported a significant relationship between high chronological age and lower physical functioning,^52,172,257,258,266,431,433,441^ one was non-significant,^207^ and three reported mixed results.^277,337,442^ Of those mixed results, one study reported that grip strength decreased with chronological age at a steeper rate in men than women (P < .001).^277^ Jee and colleagues^337^ reported that vertical jump and grip strength were negatively correlated with chronological age (P < .01), and whole body reaction time was positively correlated with chronological age (P < .01). In their 2012 study, Jee and colleagues^442^ also found that all measures of physical functioning were associated with chronological age (P < .01), except for the sit-and-reach test in females.

### Pain

The authors of four studies compared pain to biological age, in which one reported that higher pain levels were significantly associated with shorter telomeres (P = .039)^209^ and two reported mixed results.^129,258^ Cruz-Almeida and colleagues^129^ found that greater pain completing daily activities (P < .010) and at anatomical pain sites (P < .001) were associated with older biological age, but pain frequency and duration were not. The second study indicated that chronic and joint pain were both related to greater biological age (P < .001), but not to TL.^258^ Peterson and others^355^ measured high- and low-impact knee pain and found that participants with high-impact pain had a biologically older epigenetic age, they had an older AgeAccelGrim (P < 0.001) relative to their chronological age. All groups of researchers also compared pain to chronological age, but all results were not significant.

### Cardiorespiratory

A total of 78 studies examined cardiorespiratory variables, including blood pressure, respiratory health, cholesterol levels, and other cardiovascular measurements.

### Blood Pressure

There were 48 studies that assessed blood pressure (BP) in relation to biological age, of which 14 reported significant results,^48,66,85,113,128,134,144,178,240,241,293,411,444,450^ 15 found insignificant results,^45,68,116,148,165,183,193,230,238,262,292,382,395,406,412^ and six found mixed results.^58,82,339,342,347,387^ In comparison to chronological age, five studies reported a significant relationship between BP and age,^23,92,266,377,420^ whereas two found insignificant results^349,433^ and seven reported mixed results.^150,277,291,337,412,421,442^

Overall higher BP was significantly associated with biological aging in two studies,^113,411^ increased hazard ratio for all-cause mortality in one study,^144^ and a perceived aging measure.^450^ Besides these significant results, 14 studies concluded insignificant results regarding the association between BP and biological aging, including nine studies which did not find significant relationships between BP and LTL,^45,68,165,193,238,292,382,395,412^ and one study which found no relationship between BP and mortality risk. Another study examined overall BP and LTL and reported a significant inverse relationship between LTL and systolic and diastolic BP, although this was insignificant when age was controlled for.^82^ Of the significant results, higher systolic BP was predictive of faster biological aging according to calculated biological age or biomarkers.^128,134,240,241,347,411,444^ Seven studies observed significant associations between higher systolic BP and shorter TL,^48,58,85,178,293,339,387^ and one study indicated higher systolic BP was associated with longer LTL.^66^ Systolic BP was reported to be unrelated to the rate of biological aging in eight studies.^68,116,148,230,238,292,382,412^ Higher diastolic BP was significantly associated with shorter LTL in one study,^66^ but not in another.^339^ Another study that found higher aortic systolic BP to be significantly associated with short telomere proportion (P = .01) did not find the same significance for aortic diastolic and radial BPs.^58^ One study identified that systolic BP was significantly associated with PhenoAge, GrimAge, Hannum and DNAm PAI-1, but not Horvath, whereas diastolic BP was significantly associated with GrimAge, but not Horvath, PhenoAge, Hannum or DNAm PAI-1.^342^

Fourteen studies compared BP to chronological age, of which the authors of four studies reported that their measure of BP was lower among younger participants,^23,92,377,420^ one reported that sitting diastolic BP was negatively correlated with chronological age (P < .01),^266^ and two found an insignificant relationship between BP and chronological age.^349,433^ Another seven studies that reported mixed results regarding the relationship between BP and chronological age, in which the results differed by the BP measure used^291,412,421^ and gender.^150,277,337,442^

### Respiratory

There were 14 studies examining respiratory health variables. In comparison to biological age, seven found significant results,^134,243,256,258,266,303,411^ six found insignificant results,^36,135,189,262,276,293^ and one found mixed results.^115^ Biological aging was significantly related to maximal oxygen consumption (VO2 max),^134,258,303^ peak flow,^411^ forced expiratory volume at 1 second (FEV1)^243,266^ and forced vital capacity (FVC)^243^. However, a different study reported that FEV1, FVC, and the FEV1/FVC ratio were not strongly correlated with biological age.^262^ Drewelies and colleagues^135^ found that lung capacity, measured by the FEV1, was not associated with biological age. Seki and researchers^276^ also concluded that VO2 max was not associated with biological age as measured by TL. One study reported mixed results regarding biological aging, in which FEV1 was significantly associated with biological age measures for women only, but FVC was significant for both men and women.^115^ Regarding LTL, one study found that VO2 max was not an independent predictor of TL,^293^ and Kemp and Ferraro^36^ did not find any significant associations between TL and respiratory diseases during childhood. A study that assessed lung function^256^ concluded that a higher brain-PAD score was significantly associated with poorer lung function (P = .02).

In comparison to chronological age, five studies found significant results,^23,266,337,433,442^ one found insignificant results,^258^ and one was mixed.^277^ Chronological age was significantly associated with VO2 max,^337^ FEV1,^23,266,337,442^ FVC,^266,337,433,442^ expiratory breath-holding (seconds).^433^ Otherwise, Hasting and colleagues^258^ did not find any significant associations between VO2 max and chronological age, and Kuo and others^277^ found that FEV1, FVC, and the FEV1/FVC ratio were negatively associated with age, but the rates of decline varied between men and women.

### Cardiovascular

A total of 24 studies assessed cardiovascular variables other than BP and respiratory health. Out of these studies comparing cardiovascular health to biological age, 10 reported significant results,^38,58,88,107,116,218,265,293,342,427^ while ten consisted of insignificant results,94,111,230,237,339,347,387,388,412,438 and the remaining four were mixed.85,113,379,449 The ten studies reporting significant results related to biological age examined a variety of cardiovascular measurements. LTL was significantly associated with resting heart rate,^293,427^ coronary artery calcium score,^88^ pre-ejection period and respiratory sinus arrythmia,^38^ peak strain rates,^427^ mitral annulus peak E anterior wall (MVEA) and intima-media thickness (IMT),^265^ systemic vascular resistance index (SVRI) and pulse wave velocity.^58^ Roetker and others^116^ reported that cross-sectional difference in carotid IMT was a significant predictor of older biological age (P < .001). Koh and colleagues^427^ examined a variety of atrial measurements, heart rate, and cardiovascular risk factors, and found that they significantly predicted biological age. Contrastingly, ten studies did not find significant results regarding the association between biological age or biomarkers and cardiovascular variables including resting heart rate,^111,387^ pulse pressure,^237,412^ incident atrial fibrillation,^94^ ejection fraction,^339^ pulse rate,^230^ risk of heart attack,^438^ and several other electrocardiogram (ECG) and cardiovascular ultrasound measurements.^388,412^ Additionally, two studies concluded that the associations between the DiffAge measure of biological aging and markers of subclinical cardiovascular disease were only significant when adjustments for sex were made.^113,379^ Martens and colleagues^85^ found that associations between LTL quartiles and central retinal arteriolar equivalent (CRAE) and venular equivalent (CRVE), as well as their ratio (AVR) differed based on sex. Toyoshima and others^449^ found that differences between biological and chronological age were correlated with the ankle-brachial index, but not carotid intima-media thickness and pulse wave velocity.

Ten studies compared cardiovascular health to chronological age. Of these, six had significant results, in which chronological age was positively correlated to pulse pressure,^373,412^ incident atrial fibrillation,^94^ pulse rate,^433^ heart rate,^433^ arterial index for males,^377^ carotid IMT,^116^ and several other ECG and cardiovascular ultrasound measurements.^412^ One study reported that chronological age was not related to pulse rate.^291^ Three studies reported mixed results between cardiovascular variables and age; Martens and colleagues^85^ reported that AVR was not associated with chronological age in women and men separately, but each year increase in age for the total population showed a negative trend in the AVR. The two other studies found that chronological age was only associated with some cardiovascular ultrasound and ECG measurements, but not with others.^271,418^

### Reproductive and mammogram variables

Twelve studies examined the relationship between reproductive-related variables and biological aging, three of which reported significant results^232,386,432^ and four insignificant.^115,160,391,408^ Biological aging was negatively associated with number of live births,^232^ AFC,^432^ AMH levels,^432^ and age at first live birth,^232^ and a positively associated with follicle stimulating hormone levels^432^ and age at menarche.^232^ A study by Pruszkowska-Przybylska and colleagues^386^ investigated differences in epigenetic age acceleration between preeclampsic women and normotensive women, and found significant results on three aging measures (P < 0.001). Results indicated no relationship between biological age and menopausal status,^115^ AMH levels,^391^ hormone use, parity, and menopausal status^160^ and in the rate of DNA age acceleration between 8 weeks pregnant to one year post-birth.^408^ Another five studies reported mixed results.^224,366,369,389,439^ Bahra and colleagues^224^ found the number of children born to a woman to be a significant predictor of TL (P = .033), but the relationship between TL and biological age was only significant for women who had experienced child mortality, showing a negative association (P = .015), compared to those who had not (P = .716). Another study found the change in number of surviving offspring to be positively associated with TL (P = .029), however, maternal age at first birth and average inter-birth interval were not significantly associated with TL (P > .20).^389^ Kresovich and colleagues^439^ found no associations between epigenetic age acceleration and abnormal bleeding, gestational hypertension, or eclampsia.

However, a history of abnormal glucose tolerance during pregnancy was associated with increased epigenetic age by the Hannum (P = .03) and Levine (P < .01) clocks, and age at first birth was inversely associated with aging by the Hannum clock (P = 0.03), but not the Horvath or Levine clocks.^439^ A study by Shin and others^366^ found that only some plasma proteomics were correlated with menopausal age and years since menopause, while others were not. Lastly, Shirazi and colleagues^369^ reported the number of live births to be associated with biological age in post-menopausal women, but not in premenopausal women (P = NR).

Two studies examined mammogram-related variables in comparison to biological aging, one of which reported a significant relationship between TL and urinary malondialdehyde (MDA) and their association with mammographic density; mammographic density and TL were inversely associated at low levels of MDA, while there was a J-shaped association at high levels of MDA (P = .001). ^376^ The other study did not find significant results.^232^

### Hair, skin and teeth

Of the seven studies that examined facial aging, three reported significant results,^254,255^ with results indicating that perceived facial aging was significantly correlated with biological age and pace of aging.^254,255^ Two studies reported mixed results;^252,368^ one study found that facial aging was significantly related to pace of aging (P = 7.56E-10), shorter TL (P = .033), 71-CpG clock (P = .001), KDM Biological age (P = 3.81E-11), and age-related homeostatic dysregulation (P = 3.90E-12), but was not related to the 353-CpG clock or 99-CpG clock.^252^ Another study found significant associations between face-age acceleration and DNAm age (P = 5.1 x 10-6), and the epigenetic predictor of facial age was positively correlated with measured facial age (r = .66) and predicted mortality risk (P = 8.2 x 10-4), but no significant association was found between epigenetic age acceleration and face-age acceleration.^368^ Two studies found insignificant results.^422,436^ The other reported insignificant results, as there were no associations between facial aging and biological aging observed.^422^

One study measured skin-related variables and found insignificant results towards biological age, in which chronological age was associated with flatness and a decreased thickening of the epidermis (P < .001) as well as an increased amount of elastic fibers in the reticular dermis (P = .03).^451^ A single study assessed the extent of gray and white hairs with a hair whitening score in relation to aging; hair whitening was significantly associated with chronological age (P = .001).^349^ Meisel and colleagues^373^ examined teeth related variables and found significant associations with biological and chronological age. The distance from the cementoenamel junction to the bottom of the peridontal pocket was a significant positive predictor of chronological age (P < .001). Tooth loss was significantly associated with increased biological age (P < .001).^373^

### Family history

Six studies examined family history; three reported significant results,^161,283,349^ two were insignificant,^97,232^ and one was mixed.^228^ One study by Belsky and colleagues^283^ found a significant association between familial longevity and pace of aging, in which study members with more personal-history risk characteristics had a faster Pace of Aging (P < .001). Family histories of coronary artery disease (CAD) and premature coronary artery disease (PCAD) were assessed in three studies; one indicated that a family history of CAD was significantly associated with hair whitening (P = .004),^349^ another found that the relationship between family history of CAD and LTL varied between their groups of participants,^228^ and the analysis from the third study did not indicate a significant association between LTL and family history of premature coronary artery disease.^97^ One study assessed the association between family history of breast cancer, defined as having at least one first-degree relative diagnosed with it, and biological aging, and found no relationship between these variables.^232^ Han and colleagues^161^ assessed the relationship between biological age and a personal or family history of chronic disease; family history of chronic disease was a significant predictor of biological age (P < .001).

### Sleep

Five studies compared sleep to biological aging. Four of these had significant results^128,202,241,320^ and one reported insignificant results.^138^ Sleep behaviour,^320^ sleep quality^202^ and sleep duration^128,241^ were significantly associated with biological age. Peixoto and colleagues^138^ assessed whether the presence or absence of normal sleep was associated with biological age, but did not find any significant results.

### Other physical health variables

Organ functioning was assessed in four studies.^122,241,265,271^ Two of these both examined kidney functioning through Cystatin C levels (CYSC), and one also examined total lung capacity. One study found CYSC to be negatively correlated to telomere restriction fragment (P = .023, r = −.193), as well as a positive correlation between CYSC and chronological age (P = .0001, r = .405).^265^ The other study found CYSC to be an effective marker of chronological aging (P = NR), but total lung capacity was not.^271^ A study by Galkin and researchers^122^ found that people with heart, liver, and lung conditions were shown to have increased age acceleration. Wang and others^241^ also suggested that lung function was significantly associated with biological aging.

Three studies assessed own-birth related variables. One found significant results,^68^ while the other two were insignificant.^77,226^ Higher gestational age was significantly associated with longer LTL (P = .02), and subjects born preterm had shorter LTL in comparison to those born at term (P = .003).^68^ Another study found no significant association between LTL and gestational age, birth weight, or birth height (P > .28).^77^ Similarly, Kajantie and colleagues^226^ found that LTL was not significantly associated with any measures of body size at birth; they suggest that LTL in adults could be more sensitive to psychosocial factors in early life, rather than to other early-life adversities (such as varying body size at birth).

Two studies assessed vision. Viljanen and colleagues^445^ reported mixed results due to finding that eyeball assessment had a predictive value for participants whose biological age was lower than their chronological age, but not for those who had accelerated biological aging. One other study assessed vision and hearing.^258^ Worse vision and hearing were each significantly associated with older KDM biological age (P = 7.77E-08; P = 8.51E-08, respectively), LM biological age (P = 2.00E-09; P = 5.68E-12, respectively), and worse homeostatic dysregulation (P = 5.7E-05; P = 1.34E-10, respectively). Poorer hearing was significantly associated with shorter TL (P = .037), but vision was unrelated to TL. Vision was not correlated with chronological age (r = .24, P = NR), whereas hearing was moderately correlated with increased chronological age (r = .58). One study that assessed hearing through audiometric measurements found results suggesting that auditory functions were significantly associated with biological aging.^241^

Two studies compared metabolism and metabolic variables to biological aging. A study by Hawn and others^220^ concluded that metabolic pathology, measured by a MetS score, was significantly associated with GrimAge residuals. Sol and colleagues^434^ identified that 37 metabolites were significantly associated with biological age.

A study by Vetter and others^314^ examined Vitamin D levels and found that participants who were deficient and untreated had higher biological age. Their results showed that Vitamin D–deficient participants who chose to start supplementation showed a 2.6-year lower 7-CpG DNAmAA (p = 0.011) and a 1.3-year lower Horvath DNAmAA (p = 0.042) compared to the untreated and vitamin D–deficient participants.

A study by Zhang and colleagues^452^ assessed breast cancer risk in relation to biological aging and found mixed results. They examined 22 phthalate and phenol analytes, and 11 associations of these with breast cancer risk were modified by leukocyte TL. For the majority of the analytes, concentrations were inversely associated with breast cancer risk at a shorter LTL and then became positively associated with breast cancer risk at a longer LTL.^452^

### Biological aging algorithms and clocks

In total, 331 studies assessed calculated either biological age or acceleration and compared this to chronological aging. The most common clock used was Horvath’s 353-CpG Pan-Tissue clock, followed by Levine’s PhenoAge, and then the Hannum 71-CpG clock. There was dataset overlap in the studies included in this section, including with the Dunedin Cohort,^153,163,189,252,254,255,283^ the Lothian Birth Cohort of 1936,^61,114,256,319^ the FACHS population,3,6,41,42,119,125,162,167,171,174,280 the NHANES study population,23,31,34,240,258,291,301,369,437 the Framingham Heart Study,114,182,295,363,364 the NAS,114,117,254,318,371,372,453 the ARIC COHORT,^116,326,342^ the Australian Mammographic Density Twins and Sisters Study (AMDTSS),^232,454^ the Berlin Aging Study,^75,135,269,314,383,438^ the CALERIE trial,^254,306,307^ the Canadian Longitudinal Study on Aging (CLSA),^124,262^ the CARDIA study population,^137,212,338^ the Finnish Twin Cohort (FTC),109,203 the HRS,104,105,120,126,127,136,156,158,187,281,282,411 the HANDLS study,^140,141,170^ the Irish Longitudinal Study on Aging (TILDA),^172,430^ the Korean KNHANES study,^103,161,337^ the Moli-Sani study,^168,310,312^ the Sister Study population,^160,249,297,305^ the Vietnam Era Twin Registry,^200,201^ and the Women’s Health Initiative study.^233,274^ Two studies used the same dataset of a sample of Han Chinese people,^444,450^ two additional studies used the same dataset of geriatric care home employees,^98,184^ and another two studies used the same dataset of participants with experience of knee pain for at least 3 months.^355,455^

### Horvath 353-CpG Pan-Tissue Clock (2013)

A total of 90 studies utilized the Horvath’s Pan-Tissue clock to calculate biological age or intrinsic epigenetic age acceleration (IEAA).^60,65,105,106,109,113–116,120,121,123,127,128,130,132,141,158,159,166,172,181,200,201,219,221,222,230,232,233,244,246,248–250,252,254,260,267–269,274,281,285,287,295,297,298,305,307,309,313,314,317–319,321,323,342,344,345,350,351,354,357,362,368,371,372,386,391,404,425,430,439,446,448,454,456–463^ Sixty-one of these studies reported a significant relationship between this clock and chronological age,^65,105,106,113,114,118,127,128,130,132,141,158,166,181,182,201,221,222,230,246,248–250,254,260,267,269,281,284,285,295,297,298,305,307,309,313,314,317–319,321,323,342,344,345,350,351,357,371,372,386,391,404,425,430,446,448,456,461^ and 15 reported insignificant findings.^115,116,120,121,200,274,354,368,439,454,459,462,463^ Eight studies had mixed results, with results as follows. Biological age was higher than chronological age for obese patients both before and after bariatric surgery.^458^ Kankaanpää and colleagues^109^ found sex differences in biological age acceleration where males exhibited greater acceleration (P < .001), and this difference increased with age. Higgins-Chen and colleagues^244^ noted a significant DNAm deceleration for males using either Clozapine alone (P = .0024) or with other antipsychotics (p = .0053). Two studies by McEwen and researchers (2018; 2017)^362,460^ found that the preprocessing methods of samples strongly affected age correlation rates within the same clock and that the correlation between biological and chronological age was dependent on the types of CD8 T cells. Another study found that biological age increased by an equivalent 12 years across a 12-year period (SD = 3)^252^

Last, Christiansen and colleagues^457^ noted that a strong correlation between biological and chronological age increased concurrently at younger ages, but biological age decelerated at older chronological ages. Five studies reported high correlations between Horvath’s calculated age and chronological age, but did not report a significance value.^123,172,233,268,287^ Additionally, Li and colleagues^60^ found varying results for correlations between biological age and chronological age (r = .24 to r = .87).

### Hannum 71-CpG clock

The 71-CpG Hannum Clock was used to calculate biological age or extrinsic epigenetic age acceleration (EEAA) in 72 studies;^3,6,60,105,106,109,111,113–116,120,121,123,127,132,141,158,159,166,172,181,200,201,221,222,232,244,246,248–250,252,254,260,263,267,269,274,281,285,287,295,297,298,305,307,313,314,319,321,323,325,341,342,344,350,351,362,368,385,386,404,426,430,439,448,454,456,457,460,464^ authors of 53 studies reported a significant and positive relationship between Hannum’s 71-CpG clock and chronological age.^3,6,105,106,111,114,120,127,132,141,158,162,166,181,201,221,222,232,246,248–250,254,263,267,269,274,281,285,295,297,298,305,307,313,314,319,321,323,342,344,350,351,385,386,404,426,430,448,456,457,464^ Two studies reported a strong correlation between the two variables (P = NR).^172,287^ Nine studies were insignificant.^113,115,116,121,200,325,368,439,454^ Eight studies identified mixed results; Higgins-Chen and researchers^244^ indicated a sex effect favouring men, showing an age acceleration for men taking clozapine by itself (P = .0074) or alongside other antipsychotics (P = .0216), but not for women. A study by McEwen and colleagues^460^ found a significant change in results dependant on the preprocessing procedures of the same DNAm dataset, potentially skewing the clocks legitimacy. The findings of McEwen^362^ and fellow researches showed that a positive relationship to age (r = 0.86) exists, but third variables affected this correlation significantly. Li and colleagues^60^ found varying results for correlations between biological age and chronological age (r = .24 to r = .87). Starnawska and colleagues^260^ reported that this clock was overestimating DNAm age and its positive correlation to chronological age. Study members’ epigenetic clocks ticked forward by 12-14 years (for the 353 CpG clock, M=12y, SD=3; for the 99 CpG clock, M=13y, SD=4; for the 71 CpG clock, M=14y, SD=5). Across a 12-year period in one study, BA increased by 14 years (SD = 5).^252^ Sex differences in biological age acceleration were reported by Kankaanpää and colleagues,^109^ where males exhibited greater acceleration (P < .001), and this difference increased with age. Lastly, results from George and researchers^123^ indicated that the difference between biological and chronological age was less than one year (P = NR).

### Levine PhenoAge

Levine’s PhenoAge was applied in 73 studies.^60,105,106,109,120,121,123–125,127,135,136,142,158–160,166,171,172,182,183,187,200,201,215,222,230,232,244,249,250,254,258,267,269,274–276,281,282,284–287,297,298,305,307,308,314,319–321,323,335,338,342,347,350,369,371,385,386,391,404,408,411,430,437,439,448,456,461,462^ Fifty-nine studies reported PhenoAge was significantly and positively related to chronological age.^105,106,124,125,127,135,142,158,160,166,171,172,182,183,187,201,215,222,230,232,244,249,250,254,258,267,269,275,276,281,282,284–286,297,298,305,307,308,314,319–321,335,342,347,350,369,371,385,386,391,404,411,437,448,456,461,462^ Martin and colleagues^287^ found a strong relationship between PhenoAge and chronological age (P = NR). There were nine insignificant findings,^120,121,159,200,274,338,408,430,439^ and four mixed findings. A study by Li and researchers^60^ showed a significant variation between the correlations between biological age and chronological age (r =0.24 to r=0.87). Farina and colleagues^136^ found that Black and Hispanic participants had accelerated biological aging compared to White participants (p < .001), and that this was true for both male and female samples. Kankaanpää and colleagues^109^ found sex differences in biological age acceleration where males exhibited greater acceleration (P < .001), and this difference increased with age. Lastly, results from George and researchers^123^ indicated that the difference between biological and chronological age was less than one year (P = NR).

### Klemera-Doubal method

The Klemera-Doubal method (KDM) was used in 27 studies.^18,23,107,137,158,180,195,212,229,254,257,258,262,300,306,311,320,324,326,332,335,337,347,364,369,428,465^ The KDM algorithm was statistically related to chronological age in 21 studies, ^23,107,158,195,212,229,257,258,300,306,311,320,324,326,332,335,337,347,369,428,465^ whereas the KDM analysis produced four insignificant results.^18,137,262,364^ One study reported mixed results, with sex differences detected in which the effect size for women was larger than for men, likely due to women having a larger sample size of applied biomarkers.^180^ Another study indicated that per every one calendar year, KDM BA increased by only 0.71 years.^254^

### GrimAge

The mortality based GrimAge clock was used 67 times.^42,60,61,104,105,109,115,119–121,123–127,158–160,171,172,174,200–203,215,219,220,222,230,244,248–250,267,269,274–276,280–282,285,286,295,297,298,305,307,314,319,323,338,342,350,355,371,386,408,448,453,455,456,466,467^ Fifty-eight studies reported that GrimAge was significantly related to chronological age.^104,105,119,124,125,127,160,171,172,201–203,215,219,222,248–250,267,269,275,276,280–282,285,286,295,297,298,305,307,314,319,338,342,350,371,372,385,386,408,448,453,455,456,466,467^ Studies using GrimAge had 13 insignificant findings^42,60,61,115,120,121,159,174,200,230,244,274,323^ and four mixed results. O’Shea and colleagues^126^ found that chronological age was not correlated with GrimAge, but was strongly positively correlated with GrimAge rate (r = 0.78). A study by Peterson and colleagues^355^ showed that GrimAge was higher for those with high impact pain, but the overall sample had a minimal difference between biological and chronological age. Kankaanpää and researchers^109^ found sex differences in biological age acceleration where males exhibited greater acceleration (P < .001), and this difference increased with age. Lastly, results from George and colleagues^123^ indicated that the difference between biological and chronological age was less than one year (P=NR).

### Pace of Aging

Pace Of Aging was compared to chronological age in 18 studies,105,106,120,125,142,153,158,170,171,174,203,215,254,267,275,285,307,323 of which some calculated pace of aging with an algorithm titled DunedinPoAm. Authors of eleven studies reported a positive and significant correlation between pace of aging and chronological age,^105,106,120,125,142,170,171,174,203,215,254,267,307^ and four studies found insignificant biological-chronological age correlations.^153,275,285,323^

### Weidner’s 99 CpG clock

The 99-CpG clock by Weidner and researchers was used five times.^18,105,252,362,368^ There was one significant finding^105^ and one insignificant finding.^368^ Another study reported that over a 12-year period, BA according to this clock increased by an average 12 years (SD = 3).^252^

#### Horvath skin and blood clock (2018)

Six studies used the Horvath’s Skin and Blood clock.^105,230,244,357,404,461^ Five studies reported significant results with a positive linear correlation between chronological age and predicted biological age.^105,230,357,404,461^ One reported mixed finding, in which Higgins-Chen and colleagues^244^ found a significant DNAm deceleration only for males using Clozapine (P = .0024).

#### 7-CpG Vidal-Bralo

The 7-CpG Vidal-Bralo clock was used in six studies,^105,110,135,234,383,438^ with all reporting that this clock was significantly correlated with chronological age. Of these, one also reported a sex effect, in which this clock was significantly more predictive of chronological age for men compared to women, and reported minimal aging acceleration across the sample (−0.01 ± 7.81 years).^110^

### Novel clocks

Different methodologies of calculating a biological age or rate of aging were categorized as a novel clock if they were developed specifically for the study they were reported in, or if they were infrequently examined. Novel Clocks were mentioned as new methods of biological age estimators 63 times.^23,34,51,60,61,103,105,112,133,138,139,153,161,166–168,189,211,216,217,231,241,256,270,291,299,301,306,310,311,327–330,337,346,351,356–358,363,369,385,386,393,398,402,403,408,411,418,420,424,429,442–444,446,450,461–463,468^ Fifty studies reported a significant relationship between biological and chronological age.^23,34,61,103,105,133,138,139,161,166–168,189,211,216,217,231,241,299,301,306,310,311,327–330,337,351,356–358,369,385,386,393,398,402,403,411,418,424,429,442,444,446,450,461–463^ A strong correlation was found between chronological age and biological age calculated by a combination of the KDM and PCA methods.^112^ Nine studies returned insignificant results.^60,153,270,291,363,408,468^ There were four studies that reported mixed findings.^256,346,420,443^ For example, Cole and colleagues’ study^256^ showed a significant correlation between brain-predicted age and chronological age (r = 0.94) but not between brain-predicted age difference (brain-PAD) and chronological age, and detected a sex effect, with females averaging a younger brain-predicted age compared to males. Kang and others^420^ found all biomarkers used to calculate biological age in their metabolic syndrome (MS) model to be significant predictors of age (P < .001), except for pulse pressure. Another group of researchers created a biological age based on anthropometric measures, in which 76.3% and 70.5% of the variation in the body shape biological age was accounted for by chronological age for women and men, respectively.^443^ Mate and colleagues^346^ reported that biological and chronological age were correlated for the whole sample (P < .05), but biological age was elevated for participants with COPD compared to those without.

## Discussion

This scoping review included 447 studies which examined the relationship between interdisciplinary variables and the rate of biological aging. Results of this review included a comparison of biomarker and biological clocks with both chronological age and interdisciplinary variables. Most interdisciplinary variables did not demonstrate a consistent relationship with aging biomarkers, although some variables were more consistent.

Multiple indicators of biological age were examined throughout this review, including biomarkers such as TL, DNA methylation, and specific aging clocks. Of these indicators, TL was most frequently examined and yielded mostly significant results compared to chronological age. Three studies were most commonly utilized, including the Horvath’s 353-CpG Pan-Tissue clock, followed by Levine’s PhenoAge, and then the Hannum 71-CpG clock; for all three clocks, more than half of the studies that examined them reported significant results in relation to chronological age.

Compared to the interdisciplinary variables, shorter telomeres were linked to strongly influential variables such as smoking, lower SES, and chronic diseases. Aging clocks, particularly Horvath, Hannum, and GrimAge, were widely used and often showed strong associations with various aging factors. GrimAge, in particular, often showed significant associations with health risks and lifestyle factors such as smoking and disease presence, supporting its predictive utility for mortality and morbidity. Studies employing these clocks often yielded significant findings, suggesting that DNA methylation is a reliable marker for capturing nuanced aging differences. The use of different aging clocks also reveals that certain clocks may be more sensitive to specific health and lifestyle variables. For instance, GrimAge often showed stronger associations with smoking and chronic disease compared to other clocks, emphasizing that some aging markers may be more predictive of morbidity and mortality.

Throughout our review, demographic and SES factors provided the strongest evidence for influencing biological aging. Demographic factors, including sex/gender and race/ethnicity in particular, were consistently and significantly associated with biological age or rates of biological aging, often drawing conclusions from large-scale, robust datasets (e.g., NHANES, HRS, Dunedin cohort). Reviewed research consistently showed that women had lower biological age compared to men, evidenced by longer telomeres and lower rates of age-related biomarkers. This relationship was also evident across multiple aging clocks (e.g., Horvath, Hannum, PhenoAge) and diverse populations, making sex/gender one of the most strongly supported variables. Findings also consistently indicated that racial disparities, often moderated by socioeconomic factors, impacted rates of biological aging; for example, when compared to White individuals, Black individuals typically showed significantly increased biological age, although these differences diminished with SES (e.g., higher income). Higher SES in general was consistently and strongly associated with slower rates of epigenetic aging and longer telomeres, and effects were observed across multiple indicators of SES including income, education, and financial stress.

Additional physical health-related factors were strongly evidenced to influence biological age, including physical activity and smoking. Numerous studies showed that physically active individuals had longer telomeres and lower biological ages on various clocks (e.g., DNAm age, PhenoAge) compared to sedentary individuals, and that both the frequency and intensity of activity matter, with moderate to high levels of activity showing the most substantial benefits for slowing biological aging. Smoking was consistently linked to accelerated aging across biological markers where smokers were consistently biologically older than non-smokers. Findings are particularly strong for heavy and long-term smokers, but even low levels of smoking were associated with accelerated biological aging, showing that smoking’s impact on aging is dose-dependent and well-documented. Chronic conditions like cardiometabolic diseases (e.g., hypertension, diabetes) and mental health disorders (e.g., severe depression) were also strongly associated with accelerated biological aging. Research across large datasets indicated that individuals with these conditions often show increased biological age relative to chronological age, and specific biomarkers frequently showed accelerated aging in individuals with chronic diseases.

The weakest evidence for influences on biological age were evident in the environmental and built environment section of this review. Although there is interest in understanding how environmental factors might influence biological aging, findings are inconsistent, and studies are fewer in number compared to other variables. Research examining population density, type of housing, and rural versus urban settings showed mixed results, with some indicating associations with aging markers (e.g., TL) and others indicating no significant impact. The existing studies for this section were often smaller in scope and lacked the large, diverse datasets that supported findings in other areas. The impact of environmental variables also varied widely depending on study design and geographic region, making it difficult to draw firm conclusions from these results. Environmental impacts on aging are also likely influenced by numerous other interacting factors, such as SES and access to resources, which complicates the interpretation of any direct relationship with biological aging.

Additional variables outside of this section showed weaker evidence for influencing biological age in this review. Only one study reviewed the impact of sexual orientation (e.g., men who have sex with men versus men who do not) on biological aging, and it found no significant association with aging markers. The evidence is minimal, as few studies have examined this variable, and findings are insufficient to draw broader conclusions about its impact on aging. Additionally, while stress and anxiety are often associated with health outcomes, their specific impact on biological aging remained inconsistent. For example, studies on perceived or chronic stress largely showed insignificant effects on TL and other aging biomarkers, suggesting that general stress alone may not directly accelerate biological aging. The impact of anxiety disorders on aging indicators like TL also produced mixed results, as some studies showed no association, while others found small but inconsistent correlations depending on anxiety type or sample population. Social relationships as a predictor of biological aging also produced mixed findings overall, but this may be due this variable being a less concrete variable to define and difficult to measure precisely.

In the remaining sections, several overarching themes emerged around the influence of diet, early life experiences, and specific biomarkers on biological aging. Diets rich in antioxidants, fiber, and healthy fats, such as the Mediterranean diet, contributed to longer telomeres and healthier DNA methylation profiles. This points to the potential of diet as a modifiable factor for aging, particularly when it emphasizes plant-based and whole-food elements. Childhood experiences, especially trauma and SES adversity, were consistently associated with accelerated biological aging. Studies in this review indicated that individuals who experienced adverse childhood events (e.g., trauma, low parental income) often displayed telomere shortening and accelerated DNA methylation age in adulthood. This suggests that early life stress has lasting biological effects, supporting a pattern where life-course factors substantially influence aging rates.

Unlike smoking, which consistently accelerated aging, the relationship between alcohol and biological aging appears more complex. Some studies suggested that moderate alcohol consumption may have minimal or even positive effects on aging markers like TL, while heavy alcohol consumption accelerated aging. These mixed results were surprising and highlight the possibility that alcohol’s impact may depend on factors like drinking patterns, lifestyle context, and individual metabolic differences. Although physical activity generally slowed biological aging, the optimal exercise amount and intensity were unclear. Some studies found that moderate physical activity provided the most benefit, while excessive or very intense exercise did not offer additional aging benefits and, in some cases, was correlated with biomarkers of stress. This finding was also surprising, as it suggests that there may be a threshold where exercise benefits diminish, challenging the idea that “more is always better”. These patterns underscore a nuanced view of biological aging, where modifiable factors such as diet and early life experiences can significantly shape long-term health trajectories. They also highlight areas needing further research, such as the specific effects of alcohol and optimal levels of physical activity for aging.

Several variables were very commonly examined across the subsections of this review, often with significant results. TL was one of the most frequently examined indicators of biological aging, and many studies linked shorter telomeres with factors like smoking, lower SES, and chronic disease. TL frequently yielded significant results, particularly in relation to lifestyle variables (e.g., smoking, physical activity) and demographic indicators (e.g., age, sex).

Similarly, aging clocks, particularly Horvath, Hannum, and GrimAge, were widely used and often showed strong associations with various aging factors. Smoking was also commonly examined and almost always showed a significant relationship with accelerated aging, affecting both TL and methylation clocks. Indicators of SES, such as income, education, and financial stress, were widely studied and often significantly associated with biological aging. Higher SES typically correlated with slower aging, while lower SES was associated with telomere shortening and accelerated DNA methylation age. Sex and gender differences were also commonly reported, with women generally displaying slower biological aging than men across various markers. Significant findings for sex differences appear in both TL studies and epigenetic aging clocks, with women often having longer telomeres and lower biological ages than men. These commonly examined variables not only had a high frequency of study but also tended to yield significant findings. This likely reflects the robust, measurable effects of these factors on biological aging. Their strong and consistent associations suggest that they may be some of the most reliable indicators in aging research, providing clear links between lifestyle, demographic, and social factors and biological aging rates.

Focusing heavily on commonly examined variables has both benefits and potential drawbacks. Examining these variables more frequently may help to refine our understanding of reliable aging markers and clarify how these indicators correlate with health risks. The consistency of these reported relationships can support the development of standardized measures in aging research, making it easier to compare findings across studies. Additionally, repeatedly finding that factors like smoking and low SES accelerate aging underscores the importance of modifiable lifestyle factors in aging and social determinants of health. The evidence provided in this review may strengthen public health recommendations and inform policy efforts to address preventable aging-related health disparities. However, focusing primarily on well-studied variables may lead to under-examining other factors that could also play important roles in aging. This may limit our understanding of the full spectrum of aging influences and potentially overlook modifiable factors. We are also more likely to see relationships between variables if they are examined more frequently, and the frequency of significant findings may be artificially elevated (or diminished) due to the sheer number of times something was examined. Additionally, some of the most commonly examined variables may not be of clinical importance. For example, telomeres were the most common biological indicator examined in the research included but are not utilized commonly by clinicians. Thus, other variables that are more accessible for clinicians to consider in their practice may be more useful to examine in future research.

This review also has intersectional and interdisciplinary implications for biological aging research. Results showed that sex/gender, race/ethnicity, and SES clearly influence biological aging, and often interact with each other in compounding ways. Across sections in this review, these variables are noted to produce mixed findings for many variables influencing biological age. Additionally, the repeated studying of the same variables may have intersectional implications, as this can create research gaps in understudied populations or less-explored aging dimensions. For example, factors like sexual orientation or exposure to specific psychosocial stresses may yield meaningful insights if given more attention, especially for marginalized groups who experience unique aging influences due to intersectionality. It is clear that the field of social determinants of health was well represented in this review and has strong evidence for influencing biological age across diverse populations.

### Broader Implications

As our results highlight the potential utility of a multitude of variables for examining the rate of aging, future researchers should seek to identify reliable biomarkers of aging that are practical for clinical use. For example, considerations should be made to the cost, accuracy, time, and ease of sample collection,^469^ which may limit uptake in healthcare practices. Researchers should prioritize feasibility and reliability of these measures, with a focus on investigating conflicting findings related to biological age (e.g. alcohol consumption and physical activity).

When examining aging, researchers should consider a wide array of biomarkers, as our results indicated variability in the relationship between each biomarker and aging. This suggests that relying on multiple biomarkers may provide a more accurate and nuanced understanding of the aging process and its diverse biological influences. As many of the studies reviewed in the current paper were cross-sectional, there is a need for longitudinal research assessing how biological aging variables change over time, to provide insight that may otherwise be overlooked. Researchers should also consider prioritizing the study of modifiable factors that healthcare providers can realistically incorporate into practice, such as diet, exercise, and social support, and what variables could be incorporated further.

For clinicians and healthcare providers, our findings outline the importance of targeting modifiable risk factors, allowing for earlier identification of risks increasing biological aging. Further, results for both modifiable and non-modifiable risk factors may be useful for clinicians when creating treatment plans, determining potential prognoses, and tailoring preventive strategies to support healthier aging outcomes. By incorporating biological age assessments into patient evaluations, healthcare providers could improve treatment outcomes and create individualized care plans more effectively. Additionally, our results can inform gender-specific care; recognizing that women may be biologically younger on average (e.g., longer telomeres), but may face more health challenges in older adulthood (e.g., frailty).

Policymakers could utilize insights to shape public health initiatives focusing on identified modifiable risk factors that are consistently associated with biological age. Our results indicate that addressing health disparities is crucial to reducing advanced biological aging; policy makers should consider ways to create equitable health policies to address the relationships between socioeconomic factors and biological aging outlined in this paper.

These findings have implications for aging interventions and strategies, highlighting the importance of integrating lifestyle interventions—such as diet and exercise—into programs aimed at promoting longevity and quality of life. A collaborative approach across interdisciplinary researchers, healthcare providers, and policymakers is essential to effectively address risk factors of increased biological aging.

### Comparison to Previous Reviews

Earlier reviews have examined the advantages and limitations of various methods for predicting biological age and have examined specific and limited biomarkers and variables.^470,471^ These analyses have been limited in scope and have often not utilized an interdisciplinary perspective. Researchers have consistently highlighted the need for greater emphasis on this approach in future studies.^472,473^ Our review intended to fill this gap. Findings from the current review increase evidence of the need for standardization of biological aging measures.^474,475^ Similar to previous findings, we found strong evidence of clocks (particularly Horvath’s, Levine, and the 7-CpG clock) along with TL as predictive measures of biological aging.^475,476^ Strong variables that impacted biological age were supported in past reviews such as gender differences,^472,474,477^ adverse childhood relationships,^476^ socioeconomic status,^472,474^ and physical activity.^476,477^ We found an inconsistency among papers when looking at alcohol use and aging, which was similar to findings in previous reviews.^476,478^ We found environment and population to be some of the variables with weakest evidence, which contradicts past reviews that show they are strong predictors of biological aging.^473,479^

### Certainty and Limitations of Evidence

The validity of our results is strengthened by the number of included studies. By utilizing a broad selection criteria, included studies came from a wide range of disciplines, such as psychology, biological, and nursing. The number of included studies in this review is noteworthy, as the substantial literature breadth allows for the examination and comparison of a much larger range of variables, providing a more comprehensive and holistic view of the factors influencing aging. The large sample of studies also enhances the reliability of our findings by reducing potential biases that could arise from a smaller dataset.

One limitation of this review is the lack of risk of bias assessment. As studies varied in their methodology, outcomes, and populations, assessing risk of bias was not feasible and would not produce comparable results. Dataset overlap was another limitation; as many of the studies used multiple datasets, merging studies together by their dataset was not possible.

## Conclusion

Given the multitude of research examining aging from different research fields, a comprehensive examination of biomarkers and interdisciplinary variables in relation to aging is important in order to draw well-rounded and informed conclusions. The various biomarkers did not have a completely consistent relationship with chronological age, but some biomarkers and clocks demonstrated a stronger and more reliable association, such as telomere length. Of the interdisciplinary variables, we found that demographic variables (i.e., sex/gender, race/ethnicity), socioeconomic status, smoking, chronic health conditions, and childhood experiences demonstrated some of the strongest and most consistent relationships with the rate of biological aging.

## Supporting information

Table S2 - Data Extraction

Table S1 - Inclusion and Exclusion Criteria

Appendix A - Search Strategy

## Data Availability

All secondary data produced in the present study are available upon reasonable request to the authors.

## Acknowledgments

The authors would like to thank Reza Sayar and Victoria Pickles for their support in the database searches, Yaël Stevens for general research support and managing article references, Lena Fellenius for support in the analysis, Lea Friedemann for data extraction support, and Jessica Mussell for support in developing our search strategy.

