## Supplementary material for "The Relationship Between Biological Aging with Interdisciplinary Health Indicators: A Scoping Review": Table S1 - Inclusion and Exclusion Criteria

**Table 1: Outcome Categories of Included Studies.**

| Outcome category | Definition | Example variables |
|---|---|---|
| Molecular biological indicators | Variables at the molecular scale within the biology field. | Telomere length, mitochondrial copy number, telomerase activity, DNA methylation, basal oxidation, terminal restriction fragment length (TRF). |
| Physical health indicators | Physiological indicators which could be assessed by a visit to a physician. | Frailty, cardiorespiratory, reproductive outcomes, anthropometric measurements, family history of disease. |
| Mental health and substance use indicators | Outcomes related to mental health, stress and trauma, or the consumption of substances, excluding those prescribed by a physician. | Diagnosis of a mental disorder, stress, PTSD, emotions, abuse, adverse life events. |
| Cognition and neurology indicators | Variables which are related physiologically to the brain or to cognition. | Cognitive impairment, neuropsychiatric symptoms, memory, intelligence, mind wandering, rumination. |
| Social Relationships and Interpersonal Systems indicators | The social sphere; variables relating to social interactions. | Romantic relationship quality or duration, contact with children, neighbourhood-related variables, discrimination. |
| Demographic indicators | Who an individual is; variables which are not chosen. | Sex/gender, ethnicity/race, sexuality, education, income. |
| Lifestyle and environment indicators | What an individual chooses to do; variables which are impacted by choice, and which can be changed. | Athletics, sedentary activity, smoking, alcohol consumption, engagement in meditation. |
| Health outcomes indicators | Variables which may be the targeted outcome of an intervention by a physician or care team. | Quality of life, mortality risk, emergency room visits, self-reported aging, life satisfaction. |
| Chronic illness indicators | Variables related to diagnosed physical disease, chronic illnesses, and related treatments. | Diagnosed infection, prescribed medication, disabilities, tumours, chemotherapy, HIV intervention. |
| Algorithms, models, and clocks | A calculated biological age or rate of aging via an algorithms. | Horvath’s Skin & Blood Clock, GrimAge, Klemera Doubal method. |
