## Appendix A - Search Strategy for "The Relationship Between Biological Aging with Interdisciplinary Health Indicators: A Scoping Review"

### PubMed Search

| Search number | Query |
|---|---|
| 1 | "Aging" [Title/Abstract] OR "Epigenetics" [Title/Abstract] OR "Telomeres"[Title/Abstract] |
| 2 | "biological age" [Title/Abstract] OR "biological ageing" [Title/Abstract] OR "biological aging" [Title/Abstract] |
| 3 | telomer* [Title Abstract] OR methyl* [Title/Abstract] OR DNAm[Title/Abstract] OR "epigenetic age" [Title/Abstract] OR "epigenetic ageing " [Title/Abstract] OR "epigenetic aging" [Title Abstract] OR "age estimator" [Title/Abstract] OR "epigenetic clock" [Title/Abstract] OR "biological clock" [Title/Abstract] OR "methylation clock" [Title/Abstract] OR "age predictor" [Title/Abstract] OR "age estimation" [Title/Abstract] OR "age acceleration" [Title/Abstract] OR "accelerated age" [Title/Abstract] OR "estimate age" [Title/Abstract] OR "Dnam Age" [Title/Abstract] OR "methylation age" [Title/Abstract] OR methylation profile [Title/Abstract] OR "aging rate" [Title/Abstract] OR "ageing rate" [Title/Abstract] OR "biological markers "[Title/Abstract] OR biomarkers [Title/Abstract] OR "chronological age" [Title/Abstract] OR "chronological aging" [Title/Abstract] OR "chronological ageing" [Title/Abstract] |
| 4 | (((((DNA Methylation [MeSH Terms]) OR (Epigenesis, Genetic [MeSH Terms])) OR (Telomere Shortening [MeSH Terms])) OR (Biological Clocks/Physiology [MeSH Terms])) OR (Biological Clocks/Genetics [MeSH Terms])) OR (Aging/Genetics [MeSH Terms]) |
| 5 | #1 OR #3 |
| 6 | #2 AND #4 Filters: from 2011 – |
| 7 | #2 AND #4 Filters: Humans, from 2011 - |
| 8 | #2 AND #4 Filters: Humans, Adult: 19+ years, from 2011 |
| 9 | #2 AND #4 Filters: Humans, Adult: 19+ years, English, from 2011 - |

### CINAHL Search: Abstract search

| Search number | Query | Limiters |
|---|---|---|
| 1 | AB "Aging" OR "Epigenetics" OR "Telomeres" | Expanders - Apply equivalent subjects Search modes - Boolean/Phrase |
| 2 | AB "biological age" OR "biological ageing" OR "biological aging" | Search modes - Boolean/Phrase |
| 3 | AB telomer* OR methyl* OR DNAm OR "epigenetic age" OR "epigenetic ageing" OR "epigenetic aging" OR "age estimator" OR "epigenetic clock" OR "biological clock" OR "methylation clock" OR "age predictor" OR "age estimation" OR "age acceleration" OR “predict age” OR "accelerated age" OR “estimate age” OR | Search modes - Boolean/Phrase |

| | | |
|---|---|---|
| | “Dnam Age” OR “methylation age" OR "methylation profile" OR "model of aging" OR "model of ageing" OR "aging rate" OR "ageing rate" OR "biological markers" OR biomarkers OR "chronological age" OR "chronological aging" OR "chronological ageing" | |
| 4 | S1 OR S3 | Search modes - Boolean/Phrase |
| 5 | S2 AND S4 | Search modes - Boolean/Phrase |
| 6 | S2 AND S4 | Limiters - Published Date: 20210901-; English Language; Human; Age Groups: All Adult |

**CINAHL Search: Title Search**

| Search number | Query | Limiters |
|---|---|---|
| 1 | TI "Aging" OR "Epigenetics" OR "Telomeres" | Expanders - Apply equivalent subjects Search modes - Boolean/Phrase |
| 2 | TI "biological age" OR "biological ageing" OR "biological aging" | Search modes - Boolean/Phrase |
| 3 | TI telomer* OR methyl* OR DNAm OR "epigenetic age" OR "epigenetic ageing" OR "epigenetic aging" OR "age estimator" OR "epigenetic clock" OR "biological clock" OR "methylation clock" OR "age predictor" OR "age estimation" OR "age acceleration" OR “predict age” OR "accelerated age" OR “estimate age” OR “Dnam Age” OR “methylation age" OR "methylation profile" OR "model of aging" OR "model of ageing" OR "aging rate" OR "ageing rate" OR "biological markers" OR biomarkers OR "chronological age" OR "chronological aging" OR "chronological ageing" | Search modes - Boolean/Phrase |
| 4 | S1 OR S3 | Search modes - Boolean/Phrase |
| 5 | S2 AND S4 | Search modes - Boolean/Phrase |
| 6 | S2 AND S4 | Limiters - Published Date: 20210901-; English Language; Human; Age Groups: All Adult |

**PsycINFO Search: Abstract search**

| Search number | Query | Limiters |
|---|---|---|
| 1 | MA "Aging" OR MA "Epigenetics" OR MA "Telomeres" | Search modes - Boolean/Phrase |
| 2 | AB "biological age" OR "biological ageing" OR "biological aging" | Search modes - Boolean/Phrase |
| 3 | AB telomer* OR methyl* OR DNAm OR "epigenetic age" OR "epigenetic ageing" OR "epigenetic aging" OR "age estimator" OR "epigenetic clock" OR "biological clock" OR "methylation clock" OR "age predictor" OR "age estimation" OR "age acceleration" OR “predict age” OR "accelerated age" OR “estimate age” OR “Dnam Age” OR “methylation age" OR "methylation profile" OR "model of aging" OR "model of ageing" OR "aging rate" OR "ageing rate" OR "biological markers" OR biomarkers OR "chronological age" OR "chronological aging" OR "chronological ageing" | Search modes - Boolean/Phrase |
| 4 | S1 OR S3 | Search modes - Boolean/Phrase |
| 5 | S2 AND S4 | Search modes - Boolean/Phrase |
| 6 | S2 AND S4 | Limiters - Publication Year: 2011-; Age Groups: Adulthood (18 yrs & older); Population Group: Human |

**PsycINFO Search 2: Title search**

| Search number | Query | Limiters |
|---|---|---|
| 1 | MA "Aging" OR MA "Epigenetics" OR MA "Telomeres" | Search modes - Boolean/Phrase |
| 2 | TI "biological age" OR "biological ageing" OR "biological aging" | Search modes - Boolean/Phrase |
| 3 | TI telomer* OR methyl* OR DNAm OR "epigenetic age" OR "epigenetic ageing" OR "epigenetic aging" OR "age estimator" OR "epigenetic clock" OR "biological clock" OR "methylation clock" OR "age predictor" OR "age estimation" OR "age acceleration" OR “predict age” OR "accelerated age" OR “estimate age” OR “Dnam Age” OR “methylation age" OR "methylation profile" OR "model of aging" OR "model of ageing" OR "aging rate" OR "ageing rate" OR "biological markers" OR biomarkers OR "chronological age" OR | Search modes - Boolean/Phrase |

| | | |
|---|---|---|
| | "chronological aging" OR "chronological ageing" | |
| 4 | S1 OR S3 | Search modes - Boolean/Phrase |
| 5 | S2 AND S4 | Search modes - Boolean/Phrase |
| 6 | S2 AND S4 | Limiters - Publication Year: 2011-; Age Groups: Adulthood (18 yrs & older); Population Group: Human |

**SPORTDiscus Search: Abstract search**

| Search number | Query | Limiters |
|---|---|---|
| 1 | AB "Aging" OR "Epigenetics" OR "Telomeres" | Expanders - Apply equivalent subjects Search modes - Boolean/Phrase |
| 2 | AB "biological age" OR "biological ageing" OR "biological aging" | Search modes - Boolean/Phrase |
| 3 | AB telomer* OR methyl* OR DNAm OR "epigenetic age" OR "epigenetic ageing" OR "epigenetic aging" OR "age estimator" OR "epigenetic clock" OR "biological clock" OR "methylation clock" OR "age predictor" OR "age estimation" OR "age acceleration" OR “predict age” OR "accelerated age" OR “estimate age” OR “Dnam Age” OR “methylation age" OR "methylation profile" OR "model of aging" OR "model of ageing" OR "aging rate" OR "ageing rate" OR "biological markers" OR biomarkers OR "chronological age" OR "chronological aging" OR "chronological ageing" | Search modes - Boolean/Phrase |
| 4 | S1 OR S3 | Search modes - Boolean/Phrase |
| 5 | S2 AND S4 | Search modes - Boolean/Phrase |
| 6 | S2 AND S4 | Limiters - Published Date: 20210901-; Language: English |

**SPORTDiscus Search: Title search**

| Search number | Query | Limiters |
|---|---|---|
| 1 | TI "Aging" OR "Epigenetics" OR "Telomeres" | Search modes - Boolean/Phrase |
| 2 | TI "biological age" OR "biological ageing" OR "biological aging" | Search modes - Boolean/Phrase |

| | | |
|---|---|---|
| 3 | TI telomer* OR methyl* OR DNAm OR "epigenetic age" OR "epigenetic ageing" OR "epigenetic aging" OR "age estimator" OR "epigenetic clock" OR "biological clock" OR "methylation clock" OR "age predictor" OR "age estimation" OR "age acceleration" OR “predict age” OR "accelerated age" OR “estimate age” OR “Dnam Age” OR “methylation age" OR "methylation profile" OR "model of aging" OR "model of ageing" OR "aging rate" OR "ageing rate" OR "biological markers" OR biomarkers OR "chronological age" OR "chronological aging" OR "chronological ageing" | Search modes - Boolean/Phrase |
| 4 | S1 OR S3 | Search modes - Boolean/Phrase |
| 5 | S2 AND S4 | Search modes - Boolean/Phrase |
| 6 | S2 AND S4 | Limiters - Published Date: 20210901-; Language: English |
